# Public Support for Tobacco Endgame Policies: A Systematic Review and Meta-analysis

**DOI:** 10.1101/2024.01.07.24300815

**Authors:** Hana Kim, Coral Gartner, Richard Edwards, Cheneal Puljević, Kylie Morphett, Dong Ha Kim, Hae-Ryoung Chun, Martin Ekdahl, Heewon Kang

## Abstract

**Introductions:** An increasing number of countries are adopting the tobacco endgame goal. High levels of public support can accelerate momentum towards implementing tobacco endgame policies. We aimed to conduct a systematic review on the level of public support for tobacco endgame policies and to examine the geographical distribution of studies, support among key populations (adolescents and young adults, people who smoke), and the association between survey design and support.

**Methods:** We searched Embase, PubMed, Scopus, Web of Science, and Google Scholar for studies published from 2013 onwards. Google was used to search the grey literature. The reference lists of included articles were hand-searched. Studies were included if they reported the proportions of people supporting one or more endgame policies. Risk of bias was assessed using the JBI checklist for prevalence studies.

**Results:** Forty-seven articles were included. New Zealand and the United States were the countries with the most studies (n=11, respectively). Three-level meta-analyses showed the highest support for mandating a very low nicotine content in tobacco products (76%, 95% CI 61–87%). Meta-regressions were performed to assess the associations of population subgroup and survey design with support levels. The level of support was lower among people who smoke compared to the general population (β range: −1.59 to −0.51). Support for some policies was lower when *neutral* or *don’t know* response options were included.

**Conclusions:** Public support for most tobacco endgame policies was high.

**Implications:** Assessing public support can assist with progressing tobacco endgame policies. Policies that are widely supported by the public may be more politically feasible to implement. Qualitative studies and trial studies can further inform communication and implementation strategies for tobacco endgame policies.

## INTRODUCTION

A growing number of tobacco control research activities are directed towards tobacco endgame policies.[1–3] The concept of the tobacco endgame refers to achieving near-zero smoking prevalence within a defined (proximate) timeframe This may require innovative policies [1] that complement conventional demand reduction measures, such as those included in the MPOWER package. Rather than the typical incremental intensification of existing measures, endgame policies address the fundamental factors that sustain the commercial tobacco market, such as the addictiveness of tobacco products (e.g., by mandating a very low nicotine content (VLNC) standard for smoked tobacco), the availability of tobacco products (e.g., by substantially reducing the number or types of tobacco retailers), or the tobacco industry’s commercial activities (e.g., by implementing a regulated market model). Because the tobacco endgame is a relatively new paradigm and endgame interventions go beyond more familiar ‘business-as-usual’ measures, public support of tobacco endgame policies is vital to facilitate policymaker considering their implementation.[4] Some countries have established tobacco endgame goals including Aotearoa/New Zealand (A/NZ) (≤ 5% by 2025),[5] Australia (< 5% by 2030),[6] Canada (5% by 2035),[7] and Ireland (≤ 5% by 2025).[8] The public health benefits of endgame policies and of achieving the tobacco endgame are clear.[8 10] However, there are still concerns about the feasibility of some of the proposed policies because their political acceptability is uncertain, and many have not been implemented or have only been implemented on a local level.[1] Widespread understanding and support among the public for the endgame concept and associated endgame policies would enhance their feasibility.[11] But there has been little systematic examination of public support for these types of bold policies or the general concept of phasing out tobacco sales.

Assessing support levels among population subgroups allows the identification of those that could effectively advocate for policy implementation and those that may require targeted education on tobacco endgame policies or more consultation.[12] In particular, identifying the level of support among people who smoke tobacco is required because such individuals would be most affected by endgame policies. Implementation and communication strategies aligned with the perceptions of people who smoke will maximise effectiveness and compliance.

Adolescents and young adults (AYAs) also represent key population subgroups for assessing the level of support for tobacco endgame policies. These subgroups would be the first to live in a society without widespread tobacco use and some endgame policies such as the tobacco free generation proposal are specifically directed at them and may result in constraints on their choices as well provide them with specific protection from harm.

Another important consideration when assessing support for tobacco endgame policies is the design of survey questionnaires, including the response options. Responses may be influenced by survey design features,[13] such as data collection methods, the wording of the questions, response options and question preambles, types of response options provided (*e*.*g*. Likert-type response, response options with timeframes for implementation), the order in which response options are presented, inclusion of a *neutral* or *don’t know* response option, and the extent of the description provided with the question. Because the public may have a limited understanding of endgame policies and their consequences, support levels may be particularly susceptible to the way the questions are asked and the response options provided. To our knowledge, there has not been a systematic review of the evidence on public support for tobacco endgame policies. Therefore, our primary objective was to systematically review the literature to identify, appraise, and synthesise existing evidence on the level of public support for the tobacco endgame goal and related policies. The secondary objectives were to identify geographical regions that have and have not assessed public support for endgame policies, assess support levels among key population subgroups (people who smoke and AYAs), and evaluate the methodologies (questionnaire designs and data collection modalities) used to measure the level of public support.

## METHODS

### Search strategy

We conducted a systematic review and meta-analysis in accordance with the registered review protocol.[14] Based on a report on the optimal combination of databases to guarantee adequate coverage,[15] Embase, PubMed, Web of Science, Scopus, and Google Scholar were searched for relevant articles and reports published from 1 January 2013 onwards. Studies carried out before 2013 were excluded as the findings may be less relevant. The database search was conducted on 28 December 2022, and was updated on 3 April and 1 June 2023. The search terms (Table S1) include two parts: terms to identify assessment of support, and terms for tobacco endgame goals or related policies based primarily on Puljević *et al*.[1] Google Scholar was searched with simplified terms, and only the first 10 pages were examined. Additionally, we searched Google using simplified terms to identify grey literature. The reference lists of the included articles were hand searched for relevant articles. After removing duplicates, two reviewers (HnK and HwK) independently screened the titles and abstracts of the articles. Subsequently, the same reviewers independently reviewed the full texts of articles identified for possible or probable inclusion through screening to assess eligibility. Any conflicts were resolved by discussion.

### Eligibility criteria

Studies were considered eligible for inclusion if they reported the proportion of the general population (including population sub-groups - people who smoke and AYAs) supporting one or more tobacco endgame policies. Studies that did not present the level of support but reported the denominator and numerator that allowed calculation of the proportion of the public supporting endgame policies were also included. Policies previously defined as inherent endgame policies in a previous review[1] were considered eligible. Conventional approaches—such as setting product standards, increasing tobacco taxes, and restricting retailer availability—were eligible only if they were worded to an extent considered sufficient to phase out smoking (Table S2). For example, “*the number of places that can sell tobacco products should be reduced by 95%*” was considered an endgame policy, whereas “*the number of places where cigarettes and tobacco could be purchased should be restricted*” was not. Therefore, some publications or estimates were excluded even though the policies with which they were concerned were explicitly framed as endgame policies (Table S2). No language restriction was applied to the searches, but articles had to have at least an English abstract to be considered eligible, as determined during screening. We included only studies with a sample size of ≥ 400 to guarantee a 5% or less margin of error. Studies were excluded if: only estimates for measures other than policy support were reported (*e*.*g*., awareness, potential behavioural responses); support was reported among groups not considered part of the general public (*e*.*g*., policymakers, tobacco control experts and advocates, tobacco retailers); only qualitative methods were used; the full text was unavailable; or they were funded by the tobacco industry. Editorials, commentaries, perspectives, and letters were also excluded unless original findings were reported.

### Data extraction

Data extraction was performed by HnK and HwK using a preprepared extraction form. After the initial extraction, each reviewer cross-checked the data extracted by the other reviewer. Ten percent of the extracted data, selected at random, were checked by a third reviewer (HC). The extraction form encompassed the year of publication and data collection, study design, geographic location and setting, description of the sample, sample size, name of the data source, mode of data collection, representativeness of the data (considered representative of the target population if a probability-based random sampling method, survey weights, or matching on demographic characteristics was applied), endgame policy assessed, and percentage estimates with 95% confidence intervals (CIs) of those showing support. Responses stronger than a neutral response (e.g., agree and strongly agree) were summed to calculate the proportion of people supporting a given policy.

Data were recorded for the whole population, people who smoke, AYA (16–24 years of age) and AYA who smoke if the requisite data were available. For studies that provided support estimates according to the smoking frequency (*e.g*., daily, occasional) of people who smoke, estimates of support among people who smoke daily were extracted because they comprised the majority of people who smoked in most studies. If estimates were provided for other population subgroups (*e*.*g*., people who smoke who have and have not attempted to quit smoking) rather than an overall estimate, both estimates were extracted. Weighted estimates were prioritised if both weighted and unweighted estimates were provided.

The survey questions and response options used to assess support levels, and the corresponding estimates for each response option in the total sample, were also extracted. Data were extracted in separate tables according to response type: Likert-type, forced-type, and options with timeframes. The corresponding author of the study was contacted if the full questionnaire was not reported in the publication.

### Quality assessments

Quality assessments of the included studies were conducted using the JBI checklist for prevalence studies.[16] The purpose of a quality appraisal was to identify how well the assessment was conducted, rather than how they were reported.[17] Therefore, we assessed the quality of relevant methodological papers, data resources, and information acquired from the corresponding author, as well as the information provided in the included publications.

### Data synthesis and analysis

Estimates of public support for each population group (general public, people who smoke, AYA, and AYA who smoke) were obtained for each endgame policy category. A box plot was generated to show estimates for each policy. Meta-analyses were conducted if at least three representative estimates of support for a policy among the general population were available. Because proportions typically do not follow a normal distribution, logit transformation was applied to the support level data before conducting the meta-analyses. We also calculated standard errors (SEs) of the proportion estimates for studies that did not provide SEs or CIs as follows:

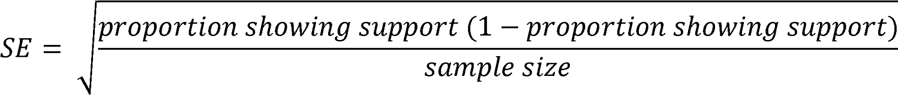

Levels of support for tobacco endgame policies may vary according to the tobacco control landscape of a country. Therefore, we conducted three-level meta-analyses which considers within-study, between-study, and between-country-heterogeneity.[18 19] Meta-analysis per policy was conducted when there were three or more estimates available among the whole population. Forest plots were used to visualise the back-transformed results of the meta-analyses. Author (publication year), sample size, and data collection year were provided in the forest plots. If needed, subcategories were indicated in the forest plots as some studies provided multiple estimates. Heterogeneity was assessed using level-specific I^2^ statistics. We used funnel plots depicting sample size against percentage supporting the policy to assess publication bias.

To evaluate the associations among population group, survey design features, and levels of support, three-level meta-regression analyses were conducted including the following variables: population/group (general population, people who smoke, AYA, and AYA who smoke), response option types (Likert, forced, timeframe), inclusion of *neutral* or *don’t know* response options (neither, one, or both), and data collection modality (to identify possible social desirability bias; face-to-face, telephone interviews, or other). The outcomes of the meta-regression analyses were logit-transformed proportion showing support. Meta-regression analyses were only conducted where there were three or more estimates of support levels per policy. β coefficients, representing the average difference in logit-transformed proportion for the reference group and the comparison group, along with their 95% CIs were calculated. Meta-analyses and meta-regressions were conducted using the rma.mv function of the *metafor* R package.

### Sensitivity analyses

As a sensitivity analysis, we conducted additional meta-analyses by excluding studies with at least one *no*, *unclear*, or *not applicable* response to the quality assessment criteria. Consistent with the approach used in the main meta-analyses, sensitivity analyses were performed on policies with three or more estimates available.

## RESULTS

### Descriptions of the included studies

Figure S3 shows the process of searching, screening, and selecting relevant publications. A total of 10,309 records were identified by searching the databases. In total, 47 records met the inclusion criteria.

The characteristics of the included publications are listed in Table 1. A total of 406,645 subjects were included (median, 2,594; range: 450–113,459). All included studies were cross-sectional in design. Of the 47 studies included in the review, 11 (23.4%) were undertaken in NZ and the United States of America (USA), and 5 (10.6%) in Australia and the United Kingdom (UK). Six studies included participants from more than one country. The geographical distribution of the included studies is shown in Figure S4. Participants in some of the studies were limited to people who smoke[20–30] or AYAs,[31–35] so 17 of the 47 included studies did not report percentage support estimates for the whole general population. For population subgroups, 36 studies provided estimates for people who smoke, 11 for AYA, and 7 for AYA who smoke. Sixteen studies collected data using a web tool, 13 by telephone, and 9 using face-to-face interviews. Paper-based surveys or those using multiple modes of administration were less common.

**Table 1.**
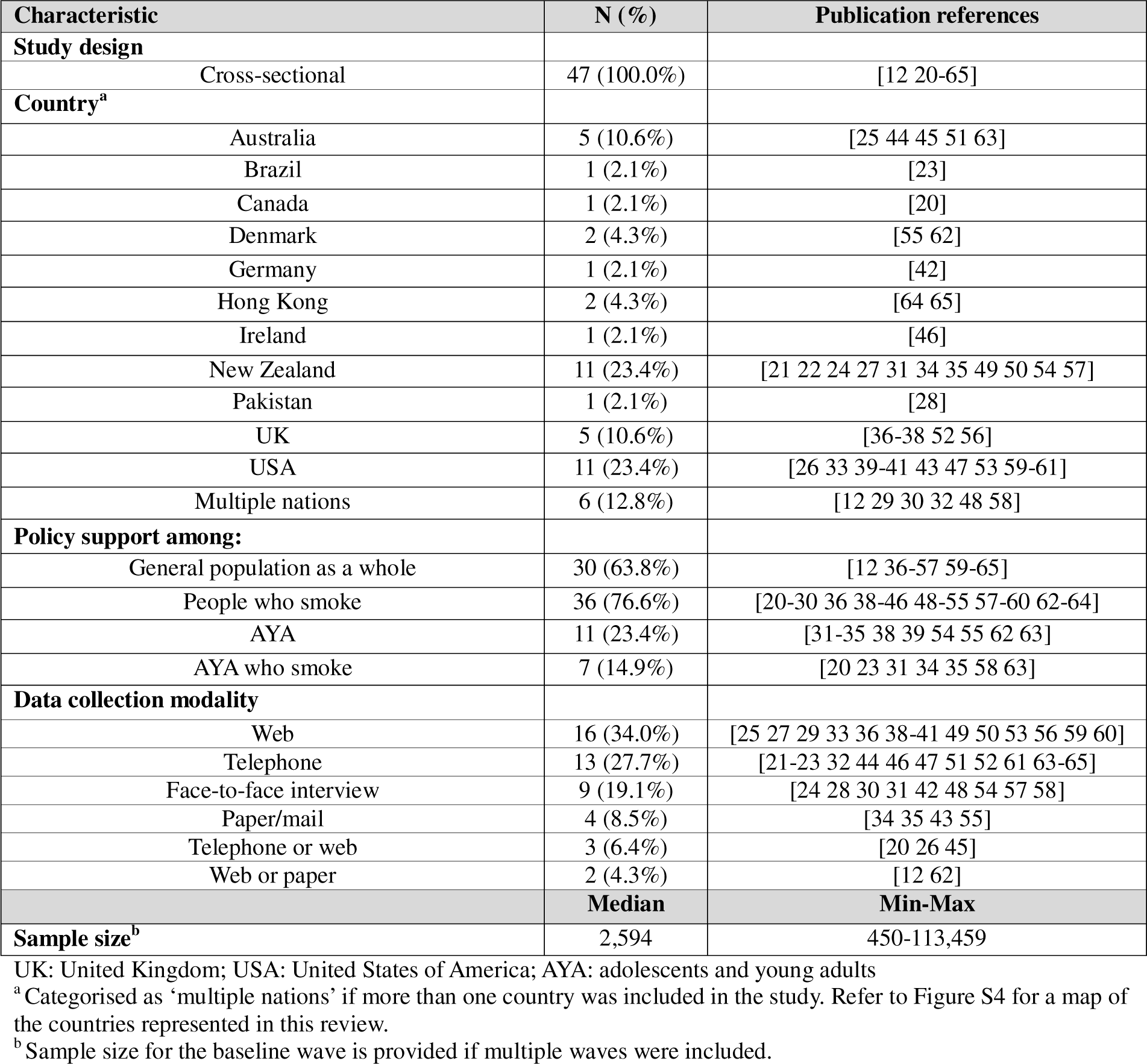
Characteristics of the included studies (N = 47)

| Characteristic | N (%) | Publication references |
| --- | --- | --- |
| **Study design** |  |  |
| Cross-sectional | 47 (100.0%) | [12 20-65] |
| **Country<sup>a</sup>** |  |  |
| Australia | 5 (10.6%) | [25 44 45 51 63] |
| Brazil | 1 (2.1%) | [23] |
| Canada | 1 (2.1%) | [20] |
| Denmark | 2 (4.3%) | [55 62] |
| Germany | 1 (2.1%) | [42] |
| Hong Kong | 2 (4.3%) | [64 65] |
| Ireland | 1 (2.1%) | [46] |
| New Zealand | 11 (23.4%) | [21 22 24 27 31 34 35 49 50 54 57] |
| Pakistan | 1 (2.1%) | [28] |
| UK | 5 (10.6%) | [36-38 52 56] |
| USA | 11 (23.4%) | [26 33 39-41 43 47 53 59-61] |
| Multiple nations | 6 (12.8%) | [12 29 30 32 48 58] |
| **Policy support among:** |  |  |
| General population as a whole | 30 (63.8%) | [12 36-57 59-65] |
| People who smoke | 36 (76.6%) | [20-30 36 38-46 48-55 57-60 62-64] |
| AYA | 11 (23.4%) | [31-35 38 39 54 55 62 63] |
| AYA who smoke | 7 (14.9%) | [20 23 31 34 35 58 63] |
| **Data collection modality** |  |  |
| Web | 16 (34.0%) | [25 27 29 33 36 38-41 49 50 53 56 59 60] |
| Telephone | 13 (27.7%) | [21-23 32 44 46 47 51 52 61 63-65] |
| Face-to-face interview | 9 (19.1%) | [24 28 30 31 42 48 54 57 58] |
| Paper/mail | 4 (8.5%) | [34 35 43 55] |
| Telephone or web | 3 (6.4%) | [20 26 45] |
| Web or paper | 2 (4.3%) | [12 62] |
|  | **Median** | **Min-Max** |
| **Sample size<sup>b</sup>** | 2,594 | 450-113,459 |

UK: United Kingdom; USA: United States of America; AYA: adolescents and young adults
<sup>a</sup> Categorised as 'multiple nations' if more than one country was included in the study. Refer to Figure S4 for a map of the countries represented in this review.
<sup>b</sup> Sample size for the baseline wave is provided if multiple waves were included.

Table 2 shows the numbers of included publications and estimates of support for each endgame policy. Most studies provided stratified estimates by policy and population group, and a few provided estimates by period (n=3) [31 32 51] and survey design (n=5).[21 44 49 51 53] A total of 235 estimates were identified; some were excluded because they included the same population[62] as a prior study.[55] Among the 47 included studies, 7 reported support estimates for an endgame goal.[21 27 31 38 46 49 50]

**Table 2.** Policy support estimates for each endgame policy and population/group (n = 47 articles).

| Policy category | Policy | Publication references | Number of publications (number of estimates) | Number of estimates per group |  |  |  |
| --- | --- | --- | --- | --- | --- | --- | --- |
|  |  |  |  | G | S | A | AS |
| Tobacco endgame goal (n = 17) |  | [21 27 31 38 46 49 50] | 7 (17) | 5 | 7 | 3 | 2 |
| Product focused (n = 21) | Mandate very low nicotine content for smoked tobacco products | [20 21 24 26 29 40 43 46 47 53 54 59-61] | 14 (30) | 10 | 17 | 2 | 1 |
|  | Set product standards for smoked tobacco products to reduce appeal and addictiveness | [20 21 23 29 30 46] | 6 (14) | 2 | 10 | 0 | 2 |
|  | Move consumers from combustible tobacco products to reduced-risk nicotine products (*e.g.*, e-cigarettes) | [25] | 1 (1) | 0 | 1 | 0 | 0 |
| User focused (n = 6) | Require consumers to obtain a license or prescription to purchase tobacco | [46 65] | 2 (3) | 2 | 1 | 0 | 0 |
|  | Restrict tobacco supplies and sales by birth year (*i.e.*, tobacco-free generation) | [46 52 63 65] | 4 (9) | 4 | 3 | 1 | 1 |
| Market/supply focused (n = 40) | Ban commercial sale of combustible tobacco | [12 21 22 28 31-35 39 41 42 44-46 48 50 51 55 57 58 62 64 65] | 24 (134) | 19 | 18 | 91 | 6 |
|  | Set a regularly reducing quota on the volume of tobacco products manufactured or imported | [65] | 1 (1) | 1 | 0 | 0 | 0 |
|  | Reduce commercial viability of tobacco companies | [28 36-38 42 46 56 58] | 8 (13) | 6 | 5 | 1 | 1 |
|  | Increase tobacco taxes to an extent where tobacco products are unaffordable | [21 38 46] | 3 (6) | 2 | 3 | 1 | 0 |
|  | Restrict tobacco retailer density/location/type/licensing to substantially limit availability | [21 45 46 50] | 4 (7) | 3 | 4 | 0 | 0 |
| Institutional structure focused (n = 0) | Transfer management of tobacco supply with a mandate to phase out tobacco sales | - | 0 | 0 | 0 | 0 | 0 |
|  | Performance-based regulation on tobacco industries | - | 0 | 0 | 0 | 0 | 0 |

G, general public; S, people who smoke; A, adolescents and young adults, AS, adolescents and young adults who smoke

Of the 21 studies on product-focused policies, 14 related to mandating a VLNC standard for smoked tobacco products.[20 21 24 26 29 40 43 46 47 53 54 59-61] Among the six studies related to setting product standards to make products less appealing, five pertained to banning all additives,[20 21 23 29 30 46] and one to banning filters.[46] Only one study focused on a policy intended to promote the use of ‘clean’ nicotine products (e.g., e-cigarettes, nicotine gum, etc.) as a substitute for cigarettes.[25] Six studies evaluated user-focused policies; two pertained to the requirement for a licence or prescription to purchase tobacco[46 65] and four to restricting tobacco sales and supplies by birth year (i.e., tobacco-free generation, TFG).[46 52 63 65]

Market/supply-focused policies were the focus of the largest number of studies (n=40). Twenty-four studies measured support for banning cigarette sales.[12 21 22 28 31-35 39 41 42 44-46 48 50 51 55 57 58 62 64 65] One study provided estimates for AYA in each of the 27 European Union member states for 2008, 2011, and 2014 (81 estimates),[32] resulting in 134 support estimates for banning cigarette sales. Eight studies focused on reducing the commercial viability of tobacco companies,[28 36-38 42 46 56 58] and three on increasing tobacco tax to an unaffordable level.[21 38 46] Among the four studies on restricting retailers, three were about restricting their density[21 45 46] and one pertained to restricting their type.[50] Table S5 reports the endgame policies addressed in each study and Tables S6.1 to S6.11 list the characteristics of those studies.

### Quality assessment

Among the 47 studies, 28 studies were at risk of bias with at least one *no, unclear,* or *not applicable* response (Figure S7). The low score for the item ‘appropriate statistical analysis and reporting’ was due to not reporting CIs for the percentages. The detailed results of the quality assessments are provided in Table S7.

### Support estimates for endgame policies

In Figure 1, box plots and median percentage support estimates are provided for each policy, in the general population (top panel) and among people who smoke, AYA, and AYA who smoke (bottom panels). Numbers of studies, estimates, and descriptive statistics per policy/group are listed in Table S8. Although there was only one relevant article/estimate, the highest support in the general population was for regularly reducing the quota for manufactured or imported tobacco products (n=1, 80.0%). Among the policies with multiple estimates, mandating a VLNC standard had the highest level of support (n=10, median=75.9%), followed by the tobacco endgame goal (n=5, median=74.0%) and reducing tobacco company viability (n=6, median=73.5%).

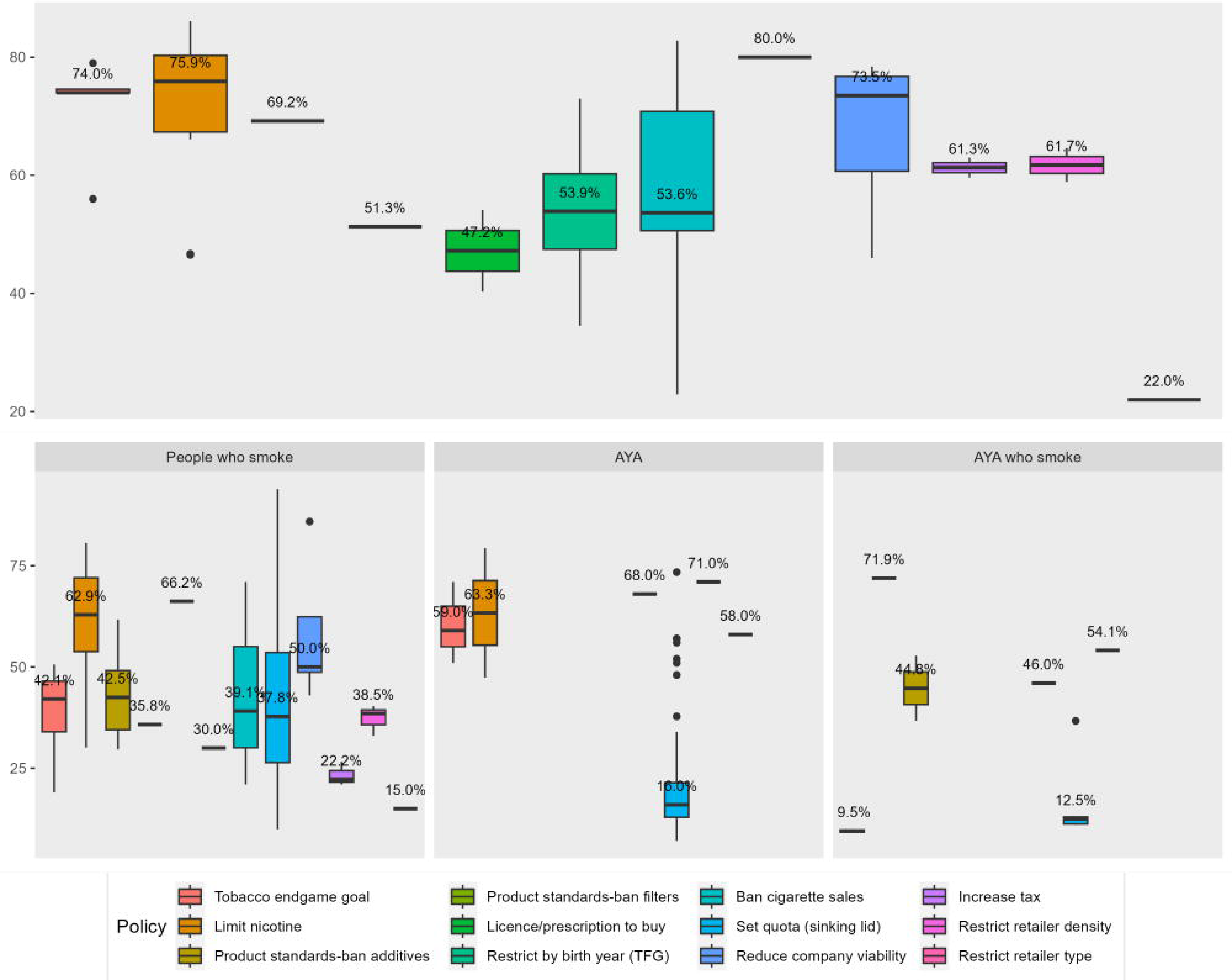

Support levels among people who smoke, AYA, and AYA who smoke were lower than those of the general public (Figure 1, lower panels). Among people who smoke (n=17, median=62.9%), AYA (n=2, median=63.3%), and AYA who smoke (n=1, 71.9%), support estimates were highest for mandating VLNC.

### Meta-analyses

We were able to conduct meta-analyses of pooled estimates of support among the general population for a tobacco endgame goal,[38 46 49 50] limiting nicotine content,[40 43 46 47 54 59] restricting sales and supply by birth year (TFG),[46 52 63 65] banning cigarette sales,[39 42 44-46 48 50 51 55 57 62 64 65] and reducing tobacco company viability.[36-38 42 56]

Forest plots for these policies are shown in Figure 2. Among the policies included in the meta-analyses, the highest support was for VLNC (panel (b); 76%, 95% CI 61–87%), followed by the tobacco endgame goal (panel (a); 72%, 95% CI 61–81%), and reducing the commercial viability of tobacco companies (panel (e); 69%, 95% CI 54–80%). Despite being estimated in the largest number of studies, support was lowest for tobacco-free generation (panel (c); 54%, 95% CI 29–77%) and banning cigarette sales (panel (d); 55%, 95% CI 38–70%).

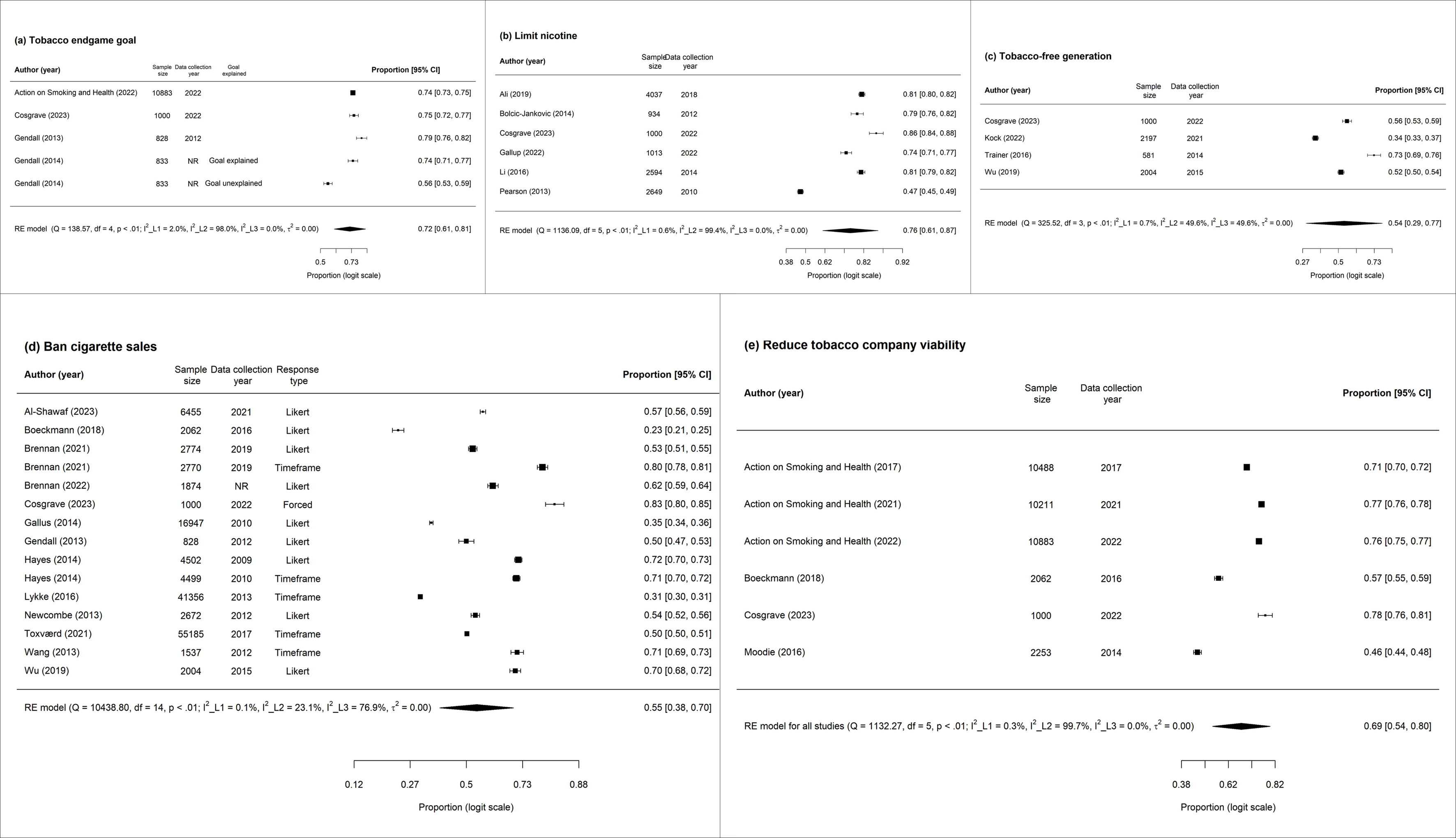

Substantial heterogeneity was identified. Within-study heterogeneity (level-1 I^2^) was small for all analyses (0.1–2.0%). Substantial between-study heterogeneity (level-2 I^2^) was identified for the tobacco endgame goal (I^2^=98.0%), VLNC (I^2^=99.4%), and reducing tobacco company viability (99.7%). The between-country heterogeneity (level-3 I^2^) ranged from 0% to 76.9%. The results of the sensitivity analyses were not materially different from the main analyses (Figure S9). Funnel plots depicting sample size against percentage support showed no evidence of publication bias (Figure S10).

### Meta-regression

Meta-regressions were performed to identify variation in levels of support by population group and survey design. Unlike the meta-analyses, which were limited to representative estimates of support for the general population, we used estimates for all population subgroups in the meta-regression analyses. Therefore, additional assessments could be performed for product standards (banning all additives), increasing tax to an unaffordable level, and restricting retailer density to reduce availability. However, four estimates from three studies were excluded from the meta-regression analyses because they did not report the size of the subgroups,[54 57 58] and two estimates were excluded from two studies[45 65] because they reported estimates for only some of the response options (ban within 5 years[45] and ban within 10 years,[65] see Table S11.3) required to calculate the proportion supporting the endgame policy.

The results of the meta-regression analyses indicated that the level of support among people who smoke was lower than in the general population for all policies analysed (Table 3). However, the difference was not statistically significant for reducing tobacco company viability (*β* −0.76, 95% CI −1.65, 0.12), increasing tax (*β* −1.67, 95% CI - 4.50, 1.16), or restricting retailer density (*β* −1.07, 95% CI −2.45, 0.31). Compared to the reference group, AYA who smoke showed the lowest level of support for the tobacco endgame goal *β*-2.86, 95% CI −3.71, −2.00), banning cigarette sales *β*-2.48, 95% CI −2.97, −1.99), restricting sales by birth year (*β*-1.24, 95% CI −2.01, −0.48), and banning all additives (*β*-0.76, 95% CI −1.20, −0.32).

**Table 3.** Meta-regression results for the association of population group and design features with level of support.

| Variables | Policy |  |  |  |  |  |  |  |
| --- | --- | --- | --- | --- | --- | --- | --- | --- |
|  | Tobacco endgame goal | Limit nicotine | Ban all additives | Restrict by birth year | Ban cigarette sales | Reduce company viability | Increase tax | Restrict retailer density |
| | $\beta$ (95% CI)* | $\beta$ (95% CI)* | $\beta$ (95% CI)* | $\beta$ (95% CI)* | $\beta$ (95% CI)* | $\beta$ (95% CI)* | $\beta$ (95% CI)* | $\beta$ (95% CI)* |
| **Population group** |  |  |  |  |  |  |  |  |
| General population | Reference | Reference | Reference | Reference | Reference | Reference | Reference | Reference |
| People who smoke | **-1.59**<br>(-1.99, -1.19) | **-0.56**<br>(-1.04, -0.08) | **-0.51**<br>(-0.91, -0.11) | **-0.52**<br>(-0.97, -0.06) | **-0.66**<br>(-0.95, -0.38) | -0.76<br>(-1.65, 0.12) | -1.67<br>(-4.50, 1.16) | -1.07<br>(-2.45, 0.31) |
| AYA | -0.37<br>(-1.03, 0.30) | -0.17<br>(-1.17, 1.50) | - | -0.33<br>(-0.94, 0.28) | **-0.39**<br>(-0.71, -0.08) | 0.32<br>(-1.17, 1.80) | -0.12<br>(-3.58, 3.34) | - |
| AYA who smoke | **-2.86**<br>(-3.71, -2.00) | -0.24<br>(-1.52, 1.05) | **-0.76**<br>(-1.20, -0.32) | **-1.24**<br>(-2.01, -0.48) | **-2.48**<br>(-2.97, -1.99) | - | - | - |
| **Response option type** |  |  |  |  |  |  |  |  |
| Likert | - | Reference | - | - | Reference | - | - | - |
| Forced | - | -0.52<br>(-1.26, 0.21) | - | - | **-0.80**<br>(-1.22, -0.38) | - | - | - |
| Timeframe | - | - | - | - | **0.74**<br>(0.32, 1.16) | - | - | - |
| **Inclusion of *Neutral* or *Don't know* among the response options** |  |  |  |  |  |  |  |  |
| Neither | Reference | Reference | Reference | - | Reference | Reference | Reference | Reference |
| One of two | -0.84<br>(-1.71, 0.02) | -0.25<br>(-0.78, 0.27) | -0.09<br>(-0.22, 0.03) | Reference | **1.01**<br>(0.59, 1.43) | 1.19<br>(-0.67, 3.05) | 0.07<br>(-4.10, 4.24) | -0.32<br>(-2.15, 1.52) |
| Both | **-1.73**<br>(-2.72, -0.74) | **-0.86**<br>(-1.64, -0.09) | - | -1.16<br>(-3.01, 0.68) | 0.39<br>(-0.13, 0.91) | -0.30<br>(-1.27, 0.68) | 0.03<br>(-3.87, 3.92) | -0.07<br>(-1.81, 1.66) |
| **Data collection modality** |  |  |  |  |  |  |  |  |
| Human (telephone or face-to-face) | Reference | Reference | Reference | - | Reference | Reference | - | - |
| Non-human (web, paper, mixed) | 0.53<br>(-0.23, 1.30) | -0.34<br>(-0.85, 0.16) | **-0.78**<br>(-1.26, -0.29) | - | -0.29<br>(-0.59, 0.01) | 0.22<br>(-1.72, 2.17) | - | - |

\* Estimates are based on logit-transformed proportions.
AYA, adolescents and young adults; CI, confidence interval
Estimates with p-values < 0.05 are shown in bold typeface.
Cells with - indicate that the level of support was not identified for the variable/category, and thus were not included in the model.

Regarding survey design, response option types, inclusion of *neutral*/*don’t know* options, and data collection modalities were examined. Most studies used Likert-type response options (Table S11.1) to assess public support. Banning cigarette sales was the only policy for which all three response options were examined. Compared to Likert-type responses, forced-type responses were associated with less support for banning cigarette sales ( *β*-0.80, 95% CI −1.22, −0.38), whereas the timeframe response option was associated with greater support (*β-*0.74, 95% CI 0.32, 1.16). Inclusion of both *neutral* and *don’t know* options was associated with decreased support for the tobacco endgame goal *β*-1.73, 95% CI −2.72, −0.74) and limiting nicotine in smoked tobacco products *β* CI −1.64, −0.09).

## DISCUSSION

Our systematic review and meta-analysis of 47 studies summarised the evidence for public support for a tobacco endgame goal and proposed policies to achieve it. The included research focused most on support for banning cigarette sales and mandating a VLNC standard, whereas support for moving consumers from smoked tobacco to reduced-risk products or requiring them to obtain a licence or prescription to purchase tobacco has received little attention. No evidence was identified for public support for institutional and market structure-focused measures.

Support among the public was high for most tobacco endgame policies. Our descriptive analyses indicated that the majority of the general population supports all endgame policies, except for requiring a licence or prescription to purchase tobacco and restricting the type of retailer that may sell tobacco products. The pooled support estimate for a mandated VLNC policy (76%) was the highest among all policies. Moreover, support for this policy was high across all population groups, indicating that support for this policy is supported even among people who smoke.[53] The support estimates for mandating VLNC policy also remained consistent even when the question was posed without any additional explanation,[26 61] suggesting that the findings are robust even when reasons for implementing are not provided. High support for the VLNC policy and its potential effect on reducing smoking prevalence[1] and improving equity,[3] suggests that it is an option that should be given strong consideration by countries seeking implementable endgame policies. Nevertheless, clear communication strategies must be developed as some studies show understanding of the policy is limited. For example, contrary to the belief of people who smoke, limiting nicotine reduces tobacco products’ addictiveness, not their harmfulness.[26]

Evidence of support for endgame policies came mainly from the USA, Europe, Australia, and NZ. Almost half of the studies included in the review were conducted in NZ and the USA. Among the 47 studies included, 44 were conducted on the continents of North and South America, Europe, and Oceania. Only three studies were conducted in Asia (two in Hong Kong and one in Pakistan), and none in Africa. Most countries that have measured public support are in the later stages of the tobacco epidemic, which are characterised by a low, and declining smoking prevalence.[66] Those in the earlier stages of the epidemic are progressing to implementation of incremental conventional measures. Together with efforts to implement conventional measures, countries in the earlier stages of the tobacco epidemic may benefit from adopting endgame policies to facilitate faster progress to the end of the tobacco epidemic. A national survey on support for tobacco endgame policies might facilitate successful adoption of endgame policies.[11] As presented in this review, such surveys will reveal strong support for endgame measures and hence may increase the priority and political will for implementation.

Establishing a tobacco endgame goal or announcing or implementing an endgame policy can facilitate examining and increasing support for these objectives. Since the declaration of the NZ endgame goal, public support for it and various endgame policies have been continuously measured among several population groups.[21 22 24 27 31 34 35 49 50 54 57] Likewise, 8 of 14 studies that examined support for VLNC policies were conducted in the USA prior to and following the announcement by the FDA of a plan to implement a policy limiting nicotine in cigarettes.[26 29 40 43 47 53 59-61] Evidence in various public health domains, including tobacco control,[67 68] have demonstrated a substantial increase in acceptability post-implementation. Measuring support repeatedly for endgame policies will be vital as increased support after public debate or policy implementation is associated with perceived effectiveness, alignment with social norms, and targeting equity.[69–72]

Studies that provided support estimates for multiple population subgroups enabled us to examine differences between subgroups using meta-regression models. Unsurprisingly, people who smoke showed lower levels of support than the general population. Hence, some governments may not be willing to start considering endgame policies until smoking prevalence has reached a sufficiently low level to ensure widespread public support. Among people who smoke,[20 29 64] intentions to quit or making quit attempts were also associated with higher support for policies including, mandating a VLNC standard, banning additives, and banning cigarette sales. This indicates that endgame policies which include strategies to support quitting will gain greater support. That said, the opinions of people who smoke should not be disregarded. In countries with endgame goals, smoking prevalence is low,[73] and people who smoke make up a minority with a weak public voice.[74 75] However, because people who smoke would be most affected by the implementation of endgame policies, priority policies and implementation plans should be accompanied by investigation of their views to facilitate and enhance implementation and effectiveness.

AYAs showed similar or slightly lower support levels for all policies compared to the general population. However, with only 23.4% of included studies measuring AYA support, a lack of data from AYAs who smoke precluded evaluation of their support levels for some policies. However, AYAs who smoke had the lowest level of support for the tobacco endgame goal, banning all additives, TFG, and banning cigarette sales. Although our data did not explain the low level of support among AYAs who smoke, a prior study suggested that adolescents who smoke may be less likely to support measures that may interfere with their continued smoking.[31]. Quantitative studies aiming to obtain estimates of support among AYAs are required, particularly for policies that have been under-represented within this population. Further, qualitative studies to identify reasons for opposition will inform communication and implementation strategies.

Examination of the impact of different survey design features was hampered by most studies using Likert-type response options to identify support. However, support for banning cigarette sales showed that the use of forced-type response options, which provided different policy options, was associated with lower levels of support. By contrast, provision of a timeframe was associated with a higher level of support. The inclusion of *neutral* and *don’t know* options was associated with lower support for the tobacco endgame goal and limiting nicotine in cigarettes, whereas it did not markedly affect extension of conventional approaches such as increasing tax and restricting retailers. This implies that the levels of support for innovative endgame policies may have been overestimated when such options were not provided or that some people who lean toward supporting the policy (rather than opposing it) feel some uncertainty about the policy. Very few people have experience with policies such as VLNC cigarettes, and some respondents may have provided socially desirable answers to a hypothetical policy. A study included in our review[53] suggested that survey designs, including the sequence of response options and the response types, have an impact on the levels of support for the VLNC policy. We suggest that the effects of survey features should be examined for a wider range of endgame policies. Methods of evaluating survey questions, such as cognitive interviewing, could be employed to examine the respondents’ understanding of survey questions. Further, support for endgame policies should be measured in trials[76] and/or purchasing experiments,[77 78] to identify the levels of support among people with some experience of the policy. Further, it is vital to measure support after the policy has been implemented to monitor public views.

Our study had several limitations. First, there is not yet a consensus on the definition of endgame policies. However, we applied policy categories that were used in previous studies.[1 2] Second, we may not have captured all studies that have examined public support for endgame policies. Although we searched multiple databases, articles that were not indexed in the selected database would not have been identified. Moreover, the sample size restriction resulted in excluding three studies that have assessed support. Third, estimates included in this review were predominantly measured in countries that are in the later stages of the tobacco epidemic. These estimates may not be generalisable to countries in the earlier stages of the epidemic. Fourth, our examination of survey design features was limited to response options and data collection modality. Some of the studies included in the review suggested that providing a more-detailed explanation of the endgame goal increased the level of support,[21 49] and providing negative response options before positive responses decreased the level of support.[53] Fifth, because of the lack of implementation of most endgame policies, the support estimates may have been biased as the respondents are unlikely to fully understand how the policies would be implemented and their impacts on both people who smoke and society in general.

Public support does not always guarantee policy effectiveness. However, it can create momentum to establish a political will to consider adopting endgame policies and to create evidence to develop implementation and communication strategies to guarantee their effectiveness. Based on our findings, we recommend the following. First, countries lacking estimates for endgame policies should gather evidence on the level of public support. There is a need for large, nationally representative estimates of the population views on which policies should be prioritised, and to identify which population group(s) may require additional consultation to understand their concerns. The survey sampling frame should be developed to provide valid estimates for the key population subgroups: people who smoke, AYAs, and AYAs who smoke. In countries with support estimates, qualitative studies among the public and key population can help identifying potential reasons for supporting/opposing. Second, the level of support should be monitored where endgame policies are being implemented to gather evidence based on experience. Repetitive measures will be required to monitor changes in support over time. Third, the effect of survey design on the levels of support for endgame policies should be evaluated in experimental studies. Design features such as response options, data collection modality, and detailed explanations of policies can be tested. In addition, qualitative studies inviting perspectives from political leaders, tobacco control advocates, experts, and stakeholders (e.g., tobacco-retailer owners) would enable refinement of the considerations involved in the implementation of endgame policies.

## CONCLUSION

This systematic review and meta-analysis showed a high level of support for tobacco endgame policies. We identified the highest support for VLNC and reducing the commercial viability of tobacco companies. Public support for some endgame policies has not been widely measured. Further research is required for regions that have yet to examine public support for endgame policies. The effects of survey design features, and reasons for support and opposition, need to be explored using a variety of study designs.

## Supporting information

The search strategy, data collection forms, data extracted from the included studies are provided in the main text or the supplementary materials

## AUTHOR STATEMENTS

A) Funding statement: This work was supported by the National Research Foundation of Korea (NRF) grant funded by the Korea government (MSIT) (No. 2021R1C1C2094375). CG is supported by an ARC Future Fellowship (FT220100186)
B) Competing Interests Statement: None declared
C) Data availability statement: The search strategy, data collection forms, data extracted from the included studies are provided in the main text or the supplementary materials. Analytic codes are available upon request.

## Data Availability

The search strategy, data collection forms, data extracted from the included studies are provided in the main text or the supplementary materials. Analytic codes are available upon request.

## ACKNOWLEDGEMENTS

We would like to acknowledge Mr Marcos Riba for providing librarian advice and support for conducting this systematic review.

