## Supplementary material for "Public Support for Tobacco Endgame Policies: A Systematic Review and Meta-analysis": The search strategy, data collection forms, data extracted from the included studies are provided in the main text or the supplementary materials

### 1. Search terms for each database

### Table S1.1. Search terms for Embase

| # | Category | | Search terms |
|---|---|---|---|
| 1 | Public support | | ('public support':ti,ab OR 'public opinion':ti,ab OR 'public view*':ti,ab OR 'public backing':ti,ab OR 'public accept*':ti,ab OR 'public attitude*':ti,ab OR 'public response*':ti,ab OR 'public perspective*':ti,ab OR 'public opposition*':ti,ab OR 'public perception*':ti,ab OR 'public emotion*':ti,ab OR 'public tolerance*':ti,ab OR support:ti,ab OR opinion:ti,ab OR view*:ti,ab OR backing:ti,ab OR accept*:ti,ab OR attitude*:ti,ab OR response*:ti,ab OR perspective*:ti,ab OR opposition* OR emotion*:ti,ab OR tolerance*:ti,ab) |
| 2 | Endgame policies | Endgame goal/ concept | (endgame*:ti,ab OR eliminat*:ti,ab OR 'phasing out':ti,ab OR 'phase out':ti,ab OR abolish:ti,ab OR abolition:ti,ab OR prohibit*:ti,ab OR ban:ti,ab)<br>AND<br>(smoking:ti OR smoke*:ti OR tobacco:ti OR cigarette*:ti OR nicotine:ti) |
| 3 | | Mandate very low nicotine levels in smoked tobacco products | ('nicotine content':ti,ab OR 'very low nicotine':ti,ab OR 'vlnc' OR 'nicotine reduc*':ti,ab OR 'reduced nicotine':ti,ab OR 'nicotine level':ti,ab OR 'maximum nicotine':ti,ab)<br>AND<br>(smoking:ti OR smoke*:ti OR tobacco:ti OR cigarette*:ti OR nicotine:ti) |
| 4 | | Set product standards for smoked tobacco products to reduce appeal and addictiveness | ('product standard*':ti,ab OR content:ti,ab OR ingredient*:ti,ab OR constituent*:ti,ab OR flavour*:ti,ab OR flavor*:ti,ab OR additive*:ti,ab OR menthol:ti,ab OR toxicant*:ti,ab OR emission*:ti,ab OR 'ph':ti,ab OR filter:ti,ab)<br>AND<br>(endgame*:ti,ab OR eliminat*:ti,ab OR 'phasing out':ti,ab OR 'phase out':ti,ab OR abolish:ti,ab OR abolition:ti,ab OR prohibit*:ti,ab OR ban:ti,ab)<br>AND<br>(smoking:ti OR smoke*:ti OR tobacco:ti OR cigarette*:ti OR nicotine:ti) |
| 5 | | Move consumers from combustible tobacco products to reduced risk nicotine products | ('alternative nicotine product*':ti,ab OR 'less harmful nicotine product*':ti,ab OR 'e-cigarette':ti,ab OR 'electronic cigarette':ti,ab OR vaping:ti,ab OR vape:ti,ab OR 'electronic nicotine delivery system*':ti,ab OR 'vaporiser*':ti,ab OR 'vaporizer':ti,ab OR snus:ti,ab OR 'vaping device':ti,ab OR 'oral snuff':ti,ab OR 'non-smoked':ti,ab OR 'non-smoked tobacco':ti,ab OR 'low nitrosamine smokeless':ti,ab OR 'reduced exposure product':ti,ab OR 'nicotine substitute':ti,ab OR 'tobacco substitute':ti,ab)<br>AND<br>(endgame*:ti,ab OR eliminat*:ti,ab OR 'phasing out':ti,ab OR 'phase out':ti,ab OR abolish:ti,ab OR abolition:ti,ab OR prohibit*:ti,ab OR ban:ti,ab)<br>AND<br>(smoking:ti OR smoke*:ti OR tobacco:ti OR cigarette*:ti OR nicotine:ti) |
| 6 | | Require consumers to obtain a license or prescription to purchase tobacco | ('smoker/s license':ti,ab OR licens*:ti,ab OR licenc*:ti,ab) NOT retail<br>AND<br>(smoking:ti OR smoke*:ti OR tobacco:ti OR cigarette*:ti OR nicotine:ti) |
| 7 | | Restrict tobacco sales and supplies by birth year | ('tobacco free generation*':ti,ab OR 'tobacco-free generation*':ti,ab OR 'age-of-sale':ti,ab OR 'age of sale':ti,ab OR 'smoke-free generation*':ti,ab OR 'smoke free generation*':ti,ab)<br>AND<br>(smoking:ti OR smoke*:ti OR tobacco:ti OR cigarette*:ti OR nicotine:ti) |
| 8 | | Ban commercial sales of combustible tobacco | (sale*:ti,ab)<br>AND<br>(endgame*:ti,ab OR eliminat*:ti,ab OR 'phasing out':ti,ab OR 'phase out':ti,ab OR abolish:ti,ab OR abolition:ti,ab OR prohibit*:ti,ab OR ban:ti,ab)<br>AND<br>(smoking:ti OR smoke*:ti OR tobacco:ti OR cigarette*:ti OR nicotine:ti) |

| # | Category | | Search terms |
|---|---|---|---|
| 9 | | Set a regularly reducing quota on the volume of tobacco products manufactured or imported | ('sinking lid':ti,ab OR 'sinking-lid':ti,ab OR quota*:ti,ab) AND (smoking:ti OR smoke*:ti OR tobacco:ti OR cigarette*:ti OR nicotine:ti)<br>AND<br>(smoking:ti OR smoke*:ti OR tobacco:ti OR cigarette*:ti OR nicotine:ti) |
| 10 | | Reduce commercial viability of tobacco companies | ('corporate manslaughter':ti,ab OR compensation:ti,ab OR litigation:ti,ab OR profitability:ti,ab)<br>AND<br>(smoking:ti OR smoke*:ti OR tobacco:ti OR cigarette*:ti OR nicotine:ti) |
| 11 | | Increase tobacco taxes to an extent where tobacco products are unaffordable | (tax*:ti,ab OR taxa*:ti,ab)<br>AND<br>(endgame*:ti,ab OR eliminat*:ti,ab OR 'phasing out':ti,ab OR 'phase out':ti,ab OR abolish:ti,ab OR abolition:ti,ab OR prohibit*:ti,ab OR ban:ti,ab)<br>AND<br>(smoking:ti OR smoke*:ti OR tobacco:ti OR cigarette*:ti OR nicotine:ti) |
| 12 | | Restrict tobacco retailer density/location/ type/licensing to substantially limit availability | (retail*:ti,ab)<br>AND<br>(endgame*:ti,ab OR eliminat*:ti,ab OR 'phasing out':ti,ab OR 'phase out':ti,ab OR abolish:ti,ab OR abolition:ti,ab OR prohibit*:ti,ab OR ban:ti,ab)<br>AND<br>(smoking:ti OR smoke*:ti OR tobacco:ti OR cigarette*:ti OR nicotine:ti) |
| 13 | | Transfer management of tobacco supply to an agency with a mandate to phase out tobacco sales | ('regulated market model':ti,ab OR monopson*:ti,ab OR 'non-profit agency':ti,ab OR 'tobacco control agency':ti,ab OR 'tobacco use management system':ti,ab)<br>AND<br>(smoking:ti OR smoke*:ti OR tobacco:ti OR cigarette*:ti OR nicotine:ti) |
| 14 | | Performance-based regulation on tobacco industries | ('polluter pays':ti,ab OR 'performance-based regulation':ti,ab OR 'performance based regulation':ti,ab OR 'performance-based agency':ti,ab OR 'performance based agency':ti,ab)<br>AND<br>(smoking:ti OR smoke*:ti OR tobacco:ti OR cigarette*:ti OR nicotine:ti) |

### Table S1.2. Search terms for PubMed

| # | Category | | Search terms |
|---|---|---|---|
| 1 | Public support | | "support"[Title/Abstract] OR "opinion"[Title/Abstract] OR "view*"[Title/Abstract] OR "backing"[Title/Abstract] OR "accept*"[Title/Abstract] OR "attitude*"[Title/Abstract] OR "response*"[Title/Abstract] OR "perspective*"[Title/Abstract] OR "opposition*"[Title/Abstract] OR "emotion*"[Title/Abstract] OR "tolerance*"[Title/Abstract] OR "public support"[Title/Abstract] OR "public opinion"[Title/Abstract] OR "public view*"[Title/Abstract] OR "public backing"[Title/Abstract] OR "public accept*"[Title/Abstract] OR "public attitude*"[Title/Abstract] OR "public response*"[Title/Abstract] OR "public perspective*"[Title/Abstract] OR "public opposition*"[Title/Abstract] OR "public perception*"[Title/Abstract] OR "public emotion*"[Title/Abstract] OR "public tolerance*"[Title/Abstract] |
| 2 | Endgame policies | Endgame goal/ concept | ("endgame*"[Title/Abstract] OR "eliminat*"[Title/Abstract] OR "phasing out"[Title/Abstract] OR "phase out"[Title/Abstract] OR "abolish"[Title/Abstract] OR "abolition"[Title/Abstract] OR "prohibit*"[Title/Abstract] OR "ban"[Title/Abstract])<br>AND<br>("smoking"[Title] OR "smoke*"[Title] OR "tobacco"[Title] OR "cigarette*"[Title] OR "nicotine"[Title]) |
| 3 | | Mandate very low nicotine levels in smoked tobacco products | ("nicotine content"[Title/Abstract] OR "very low nicotine"[Title/Abstract] OR "VLNC"[Title/Abstract] OR "nicotine reduc*"[Title/Abstract] OR "reduced nicotine"[Title/Abstract] OR "nicotine level"[Title/Abstract] OR "maximum nicotine"[Title/Abstract])<br>AND<br>("smoking"[Title] OR "smoke*"[Title] OR "tobacco"[Title] OR "cigarette*"[Title] OR "nicotine"[Title]) |
| 4 | | Set product standards for smoked tobacco products to reduce appeal and addictiveness | ("product standard*"[Title/Abstract] OR "content"[Title/Abstract] OR "ingredient*"[Title/Abstract] OR "constituent*"[Title/Abstract] OR "flavour*"[Title/Abstract] OR "flavor*"[Title/Abstract] OR "additive*"[Title/Abstract] OR "menthol"[Title/Abstract] OR "toxicant*"[Title/Abstract] OR "emission*"[Title/Abstract] OR "pH"[Title/Abstract] OR "filter"[Title/Abstract])<br>AND<br>("endgame*"[Title/Abstract] OR "eliminat*"[Title/Abstract] OR "phasing out"[Title/Abstract] OR "phase out"[Title/Abstract] OR "abolish"[Title/Abstract] OR "abolition"[Title/Abstract] OR "prohibit*"[Title/Abstract] OR "ban"[Title/Abstract])<br>AND<br>("smoking"[Title] OR "smoke*"[Title] OR "tobacco"[Title] OR "cigarette*"[Title] OR "nicotine"[Title]) |
| 5 | | Move consumers from combustible tobacco products to reduced risk nicotine products | ("alternative nicotine product*"[Title/Abstract] OR "less harmful nicotine product*"[Title/Abstract] OR "e-cigarette"[Title/Abstract] OR "electronic cigarette"[Title/Abstract] OR "vaping"[Title/Abstract] OR "vape"[Title/Abstract] OR "electronic nicotine delivery system*"[Title/Abstract] OR "vaporiser*"[Title/Abstract] OR "vaporizer"[Title/Abstract] OR "snus"[Title/Abstract] OR "vaping device"[Title/Abstract] OR "oral snuff"[Title/Abstract] OR "non-smoked"[Title/Abstract] OR "non-smoked tobacco"[Title/Abstract] OR "low nitrosamine smokeless"[Title/Abstract] OR "reduced exposure product"[Title/Abstract] OR "nicotine substitute"[Title/Abstract] OR "tobacco substitute"[Title/Abstract])<br>AND<br>("endgame*"[Title/Abstract] OR "eliminat*"[Title/Abstract] OR "phasing out"[Title/Abstract] OR "phase out"[Title/Abstract] OR "abolish"[Title/Abstract] OR "abolition"[Title/Abstract] OR "prohibit*"[Title/Abstract] OR "ban"[Title/Abstract])<br>AND<br>("smoking"[Title] OR "smoke*"[Title] OR "tobacco"[Title] OR "cigarette*"[Title] OR "nicotine"[Title]) |
| 6 | | Require a smoker's license or prescription to purchase tobacco | ("smoker's license"[Title/Abstract] OR "licens*"[Title/Abstract] OR "licenc*"[Title/Abstract]) NOT ("retail"[All Fields] OR "retailed"[All Fields] OR "retailer"[All Fields] OR "retailer's"[All Fields] OR "retailers"[All Fields] OR "retailing"[All Fields] OR "retails"[All Fields])<br>AND<br>("smoking"[Title] OR "smoke*"[Title] OR "tobacco"[Title] OR "cigarette*"[Title] OR "nicotine"[Title]) |
| 7 | | Restrict tobacco sales and supplies by birth year | ("tobacco free generation*"[Title/Abstract] OR "tobacco free generation*"[Title/Abstract] OR "age-of-sale"[Title/Abstract] OR "age-of-sale"[Title/Abstract] OR "smoke free generation*"[Title/Abstract] OR "smoke free generation*"[Title/Abstract])<br>AND<br>("smoking"[Title] OR "smoke*"[Title] OR "tobacco"[Title] OR "cigarette*"[Title] OR "nicotine"[Title]) |

| # | Category | | Search terms |
|---|---|---|---|
| 8 | | Ban commercial sales of cigarettes | ("sale*"[Title/Abstract])<br>AND<br>("endgame*"[Title/Abstract] OR "eliminat*"[Title/Abstract] OR "phasing out"[Title/Abstract] OR "phase out"[Title/Abstract] OR "abolish"[Title/Abstract] OR "abolition"[Title/Abstract] OR "prohibit*"[Title/Abstract] OR "ban"[Title/Abstract])<br>AND<br>("smoking"[Title] OR "smoke*"[Title] OR "tobacco"[Title] OR "cigarette*"[Title] OR "nicotine"[Title]) |
| 9 | | Set a regularly reducing quota on the volume of tobacco products manufactured or imported | ("sinking-lid"[Title/Abstract] OR "sinking-lid"[Title/Abstract] OR "quota*"[Title/Abstract])<br>AND<br>("smoking"[Title] OR "smoke*"[Title] OR "tobacco"[Title] OR "cigarette*"[Title] OR "nicotine"[Title]) |
| 10 | | Reduce commercial viability of tobacco companies | ("corporate manslaughter"[Title/Abstract] OR "compensation"[Title/Abstract] OR "litigation"[Title/Abstract] OR "profitability"[Title/Abstract])<br>AND<br>("smoking"[Title] OR "smoke*"[Title] OR "tobacco"[Title] OR "cigarette*"[Title] OR "nicotine"[Title]) |
| 11 | | Increase tobacco taxes to an extent where tobacco products are unaffordable | ("tax"[Title/Abstract] OR "taxa*"[Title/Abstract])<br>AND<br>("endgame*"[Title/Abstract] OR "eliminat*"[Title/Abstract] OR "phasing out"[Title/Abstract] OR "phase out"[Title/Abstract] OR "abolish"[Title/Abstract] OR "abolition"[Title/Abstract] OR "prohibit*"[Title/Abstract] OR "ban"[Title/Abstract])<br>AND<br>("smoking"[Title] OR "smoke*"[Title] OR "tobacco"[Title] OR "cigarette*"[Title] OR "nicotine"[Title]) |
| 12 | | Restrict tobacco retailer density/location/ Type/licensing to substantially limit availability | ("retail*"[Title/Abstract])<br>AND<br>("endgame*"[Title/Abstract] OR "eliminat*"[Title/Abstract] OR "phasing out"[Title/Abstract] OR "phase out"[Title/Abstract] OR "abolish"[Title/Abstract] OR "abolition"[Title/Abstract] OR "prohibit*"[Title/Abstract] OR "ban"[Title/Abstract])<br>AND<br>("smoking"[Title] OR "smoke*"[Title] OR "tobacco"[Title] OR "cigarette*"[Title] OR "nicotine"[Title]) |
| 13 | | Transfer management of tobacco supply | ("regulated market model"[Title/Abstract] OR "monopson*"[Title/Abstract] OR "non-profit agency"[Title/Abstract])<br>AND<br>("smoking"[Title] OR "smoke*"[Title] OR "tobacco"[Title] OR "cigarette*"[Title] OR "nicotine"[Title]) |
| 14 | | Performance-based regulation on tobacco industries | ("polluter pays"[Title/Abstract] OR "performance-based regulation"[Title/Abstract] OR "performance-based regulation"[Title/Abstract])<br>AND<br>("smoking"[Title] OR "smoke*"[Title] OR "tobacco"[Title] OR "cigarette*"[Title] OR "nicotine"[Title]) |

### Table S1.3. Search terms for Scopus

| # | Category | | Search terms |
|---|---|---|---|
| 1 | Public support | | TITLE-ABS ("public support" OR "public opinion" OR "public view*" OR "public backing" OR "public accept*" OR "public attitude*" OR "public response*" OR "public perspective*" OR "public opposition*" OR "public perception*" OR "public emotion*" OR "public tolerance*" OR support OR opinion OR view* OR backing OR accept* OR attitude* OR response* OR perspective* OR opposition* OR emotion* OR tolerance) |
| 2 | Endgame policies | Endgame goal/ concept | TITLE-ABS (endgame* OR eliminat* OR "phasing out" OR "phase out" OR abolish OR abolition OR prohibit* OR ban)<br>AND<br>TITLE (smoking OR smoke* OR tobacco OR cigarette* OR nicotine) |
| 3 | | Mandate very low nicotine levels in smoked tobacco products | TITLE-ABS ("nicotine content" OR "very low nicotine" OR "vlnc" OR "nicotine reduc*" OR "reduced nicotine" OR "nicotine level" OR "maximum nicotine")<br>AND<br>TITLE (smoking OR smoke* OR tobacco OR cigarette* OR nicotine) |
| 4 | | Set product standards for smoked tobacco products to reduce appeal and addictiveness | TITLE-ABS ("product standard*" OR content OR ingredient* OR constituent* OR flavour* OR flavor* OR additive* OR menthol OR toxicant* OR emission* OR "ph" OR filter)<br>AND<br>TITLE-ABS (endgame* OR eliminat* OR "phasing out" OR "phase out" OR abolish OR abolition OR prohibit* OR ban)<br>AND<br>TITLE (smoking OR smoke* OR tobacco OR cigarette* OR nicotine) |
| 5 | | Move consumers from combustible tobacco products to reduced risk nicotine products | TITLE-ABS ("alternative nicotine product*" OR "less harmful nicotine product*" OR "e-cigarette" OR "electronic cigarette" OR vaping OR vape OR "electronic nicotine delivery system*" OR "vaporiser*" OR "vaporizer" OR snus OR "vaping device" OR "oral snuff" OR "non-smoked" OR "non-smoked tobacco" OR "low nitrosamine smokeless" OR "reduced exposure product" OR "nicotine substitute" OR "tobacco substitute")<br>AND<br>TITLE-ABS (endgame* OR eliminat* OR "phasing out" OR "phase out" OR abolish OR abolition OR prohibit* OR ban)<br>AND<br>TITLE (smoking OR smoke* OR tobacco OR cigarette* OR nicotine) |
| 6 | | Require a smoker's license or prescription to purchase tobacco | TITLE-ABS ("smoke's license" OR licens* OR licenc*) AND NOT TITLE-ABS (retail)<br>AND<br>TITLE (smoking OR smoke* OR tobacco OR cigarette* OR nicotine) |
| 7 | | Restrict tobacco sales and supplies by birth year | TITLE-ABS ("tobacco free generation*" OR "tobacco-free generation*" OR "age-of-sale" OR "age of sale" OR "smoke-free generation*" OR "smoke free generation*")<br>AND<br>TITLE (smoking OR smoke* OR tobacco OR cigarette* OR nicotine) |
| 8 | | Ban commercial sales of cigarettes | TITLE-ABS (sale*)<br>AND<br>TITLE-ABS (endgame* OR eliminat* OR "phasing out" OR "phase out" OR abolish OR abolition OR prohibit* OR ban)<br>AND<br>TITLE (smoking OR smoke* OR tobacco OR cigarette* OR nicotine) |
| 9 | | Set a regularly reducing quota on the volume of | TITLE-ABS ("sinking lid" OR "sinking-lid" OR quota*)<br>AND<br>TITLE (smoking OR smoke* OR tobacco OR cigarette* OR nicotine) |

| # | Category | | Search terms |
|---|---|---|---|
| | | tobacco products manufactured or imported | |
| 10 | | Reduce commercial viability of tobacco companies | TITLE-ABS ("corporate manslaughter" OR compensation OR litigation OR profitability)<br>AND<br>TITLE (smoking OR smoke* OR tobacco OR cigarette* OR nicotine) |
| 11 | | Increase tobacco taxes to an extent where tobacco products are unaffordable | TITLE-ABS (tax* OR taxa*)<br>AND<br>TITLE-ABS (endgame* OR eliminat* OR "phasing out" OR "phase out" OR abolish OR abolition OR prohibit* OR ban)<br>AND<br>TITLE (smoking OR smoke* OR tobacco OR cigarette* OR nicotine) |
| 12 | | Restrict tobacco retailer density/location/ Type/licensing to substantially limit availability | TITLE-ABS (retail*)<br>AND<br>TITLE-ABS (endgame* OR eliminat* OR "phasing out" OR "phase out" OR abolish OR abolition OR prohibit* OR ban)<br>AND<br>TITLE (smoking OR smoke* OR tobacco OR cigarette* OR nicotine) |
| 13 | | Transfer management of tobacco supply | TITLE-ABS ("regulated market model" OR monopson* OR "non-profit agency" OR "tobacco control agency" OR "tobacco use management system")<br>AND<br>TITLE (smoking OR smoke* OR tobacco OR cigarette* OR nicotine) |
| 14 | | Performance-based regulation on tobacco industries | TITLE-ABS ("polluter pays" OR "performance-based regulation" OR "performance based regulation" OR "performance-based agency" OR "performance based agency")<br>AND<br>TITLE (smoking OR smoke* OR tobacco OR cigarette* OR nicotine) |

### Table S1.4. Search terms for Web of Science

| # | Category | | Search terms |
|---|---|---|---|
| 1 | Public support | | TS=("public support" OR "public opinion" OR "public view*" OR "public backing" OR "public accept*" OR "public attitude*" OR "public response*" OR "public perspective*" OR "public opposition*" OR "public perception*" OR "public emotion*" OR "public tolerance*" OR support OR opinion OR view* OR backing OR accept* OR attitude* OR response* OR perspective* OR opposition* OR emotion* OR tolerance*) |
| 2 | Endgame policies | Endgame goal/concept | TS=(endgame* OR eliminat* OR "phasing out" OR "phase out" OR abolish OR abolition OR prohibit* OR ban) |
| 3 | | Mandate very low nicotine levels in smoked tobacco products | TS=("nicotine content" OR "very low nicotine" OR "VLNC" OR "nicotine reduc*" OR "reduced nicotine" OR "nicotine level" OR "maximum nicotine")<br>AND<br>TI=(smoking OR smoke* OR tobacco OR cigarette* OR nicotine) |
| 4 | | Set product standards for smoked tobacco products to reduce appeal and addictiveness | TS=("product standard*" OR content OR ingredient* OR constituent* OR flavour* OR flavor* OR additive* OR menthol OR toxicant* OR emission* OR "pH" OR filter)<br>AND<br>TS=(endgame* OR eliminat* OR "phasing out" OR "phase out" OR abolish OR abolition OR prohibit* OR ban)<br>AND<br>TI=(smoking OR smoke* OR tobacco OR cigarette* OR nicotine) |
| 5 | | Move consumers from combustible tobacco products to reduced risk nicotine products | TS=("alternative nicotine product*" OR "less harmful nicotine product*" OR "e-cigarette" OR "electronic cigarette" OR vaping OR vape OR "electronic nicotine delivery system*" OR "vaporiser*" OR "vaporizer" OR snus OR "vaping device" OR "oral snuff" OR "non-smoked" OR "non-smoked tobacco" OR "low nitrosamine smokeless" OR "reduced exposure product" OR "nicotine substitute" OR "tobacco substitute")<br>AND<br>TS=(endgame* OR eliminat* OR "phasing out" OR "phase out" OR abolish OR abolition OR prohibit* OR ban)<br>AND<br>TI=(smoking OR smoke* OR tobacco OR cigarette* OR nicotine) |
| 6 | | Require a smoker's license or prescription to purchase tobacco | TS=("smoker's license" OR licens* OR licenc* NOT retail)<br>AND<br>TI=(smoking OR smoke* OR tobacco OR cigarette* OR nicotine) |
| 7 | | Restrict tobacco sales and supplies by birth year | TS=("tobacco free generation*" OR "tobacco-free generation*" OR "age-of-sale" OR "age of sale" OR "smoke-free generation*" OR "smoke free generation*")<br>AND<br>TI=(smoking OR smoke* OR tobacco OR cigarette* OR nicotine) |
| 8 | | Ban commercial sales of cigarettes | TS=(Sale*)<br>AND<br>TS=(endgame* OR eliminat* OR "phasing out" OR "phase out" OR abolish OR abolition OR prohibit* OR ban)<br>AND<br>TI=(smoking OR smoke* OR tobacco OR cigarette* OR nicotine) |
| 9 | | Set a regularly reducing quota on the volume of tobacco products manufactured or imported | TS=("Sinking lid" OR "sinking-lid" OR quota*)<br>AND<br>TI=(smoking OR smoke* OR tobacco OR cigarette* OR nicotine) |
| 10 | | Reduce commercial viability of tobacco | TS=("corporate manslaughter" OR compensation OR litigation OR profitability)<br>AND |

| # | Category | | Search terms |
|---|---|---|---|
| | | companies | TI=(smoking OR smoke* OR tobacco OR cigarette* OR nicotine) |
| 11 | | Increase tobacco taxes to an extent where tobacco products are unaffordable | TS=(Tax* OR Taxa*)<br>AND<br>TS=(endgame* OR eliminat* OR “phasing out” OR “phase out” OR abolish OR abolition OR prohibit* OR ban)<br>AND<br>TI=(smoking OR smoke* OR tobacco OR cigarette* OR nicotine) |
| 12 | | Restrict tobacco retailer density/location/ Type/licensing to substantially limit availability | TS=(Retail*)<br>AND<br>TS=(endgame* OR eliminat* OR “phasing out” OR “phase out” OR abolish OR abolition OR prohibit* OR ban)<br>AND<br>TI=(smoking OR smoke* OR tobacco OR cigarette* OR nicotine) |
| 13 | | Transfer management of tobacco supply | TS=(“regulated market model” OR monopson* OR “non-profit agency” OR “tobacco control agency” OR “tobacco use management system”)<br>AND<br>TI=(smoking OR smoke* OR tobacco OR cigarette* OR nicotine) |
| 14 | | Performance-based regulation on tobacco industries | TS=(“polluter pays” OR “performance-based regulation” OR “performance based regulation” OR “performance-based agency” OR “performance based agency”)<br>AND<br>TI=(smoking OR smoke* OR tobacco OR cigarette* OR nicotine) |

### 2. Scope of tobacco endgame policies

### Table S2. Tobacco endgame policies and examples of included/excluded policies

| Policy category | Policy | Considered an endgame policy? | Examples of included policies (as worded in the survey) | Examples of excluded policies (as worded in the survey) |
|---|---|---|---|---|
| Product focused | Mandate very low nicotine content for smoked tobacco products | Yes | • Requiring cigarette makers to lower the nicotine levels in cigarettes so that they are less addictive[1]<br>• A law that reduced the amount of nicotine in cigarettes and tobacco, to make them less addictive[2 3]<br>• The amount of nicotine in tobacco products should be reduced through new laws to make tobacco products less addictive[4]<br>• Reducing the amount of nicotine in cigarettes to help smokers quit[5] | - |
| | Set product standards for smoked tobacco products to reduce appeal and addictiveness | If framed at sufficient intensity | • A law that bans all additives, including flavourings, in cigarettes/tobacco[2 3 6 7]<br>• Added chemicals that make cigarettes seem less harsh should be banned to make cigarettes more difficult to tolerate[4]<br>• Filters on cigarettes and other combustible tobacco products should be banned to make the products more difficult to tolerate[4] | • A law that bans the use of menthol in cigarettes/tobacco[2]<br>• Prohibiting flavors such as menthol (mint), spicy, sweet, or fruity flavor, in all tobacco products[8]<br>• Banning flavors that make tobacco products more attractive[9] |
| | Move consumers from combustible tobacco products to reduced risk nicotine products (e.g., e-cigarettes) | If framed at sufficient intensity | • All smokers should be encouraged to switch to less harmful clean nicotine products as long-term substitutes for cigarettes[10] | • It should be assessed whether e-cigarettes are safe and effective in assisting smokers to quit[11] |
| User focused | Require consumers to obtain a license or prescription to purchase tobacco | Yes | • People should be required to hold an official licence to buy tobacco products[4] | - |
| | Restrict tobacco supplies and sales by birth year | Yes | • The Government should prevent everyone who is currently under 18 from ever buying tobacco products for the rest of their lives[4]<br>• Ban the sale of cigarettes and tobacco products to everyone born after a certain year from 2030 onward[12] | - |
| Market/ supply focused | Ban commercial sale of combustible tobacco | Yes | • A law that totally bans cigarettes and other smoked tobacco within 10 years[3 13]<br>• Making smoking or cigarette sales illegal[14] | - |

| Policy category | Policy | Considered an endgame policy? | Examples of included policies (as worded in the survey) | Examples of excluded policies (as worded in the survey) |
|---|---|---|---|---|
| | Set a regularly reducing quota on the volume of tobacco products manufactured or imported | Yes | • Set a quota for combustible cigarette retail and reduce it on a yearly basis[15] | - |
| | Reduce commercial viability of tobacco companies | Yes | • Tobacco industry sales should be taxed in order to use the money to address problems caused by tobacco (e.g., health issues, environmental problems, etc.)[11]<br>• Requiring tobacco manufacturers to pay a levy to government for measures to help smokers quit and prevent young people from taking up smoking[16]<br>• Tobacco companies should pay for the health-related costs associated with tobacco use[17] | - |
| | Increase tobacco taxes to an extent where tobacco products are unaffordable | If framed at sufficient intensity | • Increase the tax on tobacco by 20% a year until less than five percent of the population smoke[3 4]<br>• Tax should be used to increase the price of tobacco products 5% above the rate of inflation each year[16] | • Taxes on tobacco products should be increased[18]<br>• Tax on cigarettes should be increased every year[19] |
| | Restrict tobacco retailer density/location/type/licensing to substantially limit availability | If framed at sufficient intensity | • Tobacco product sales should only be allowed in a limited number of specially licenced shops and banned from smaller local shops, newsagents, off-licences and petrol stations[4]<br>• The number of places that can sell tobacco products should be reduced by 95%[4]<br>• Number of places that can sell tobacco products should be greatly reduced, that is by 95%, and sales allowed only in a limited number and type of stores[3]<br>• Only people qualified to give quitting advice should be allowed to sell tobacco products[20]<br>• Shop owners should seriously be thinking about transitioning out of selling cigarettes[21] | • Restrict the number of places where cigarettes/tobacco could be purchased[13]<br>• There should be fewer places where cigarettes and tobacco can be sold[22]<br>• Requiring anyone selling tobacco to have a license which can be removed if they sell to those under-age[12]<br>• Reducing the number of retailers selling cigarettes and tobacco in neighbourhoods with a high density of tobacco retailers[12]<br>• The number of places allowed to sell cigarettes and tobacco should be reduced to make them less easily available[23 24]<br>• Ban the sale of tobacco products in stores that are located near schools[25]<br>• Ban on the sale of products in drugstores[26]<br>• Requiring that tobacco be sold only in stores that sell only tobacco products and nothing else[25] |
| Institutional structure focused* | Transfer management of tobacco supply with a mandate to phase out tobacco sales | Yes | - | - |

| Policy category | Policy | Considered an endgame policy? | Examples of included policies (as worded in the survey) | Examples of excluded policies (as worded in the survey) |
|---|---|---|---|---|
| | Performance-based regulation on tobacco industries | Yes | - | - |

*Publication measuring support for institutional structure focused measures was not identified

### 3. PRISMA diagram

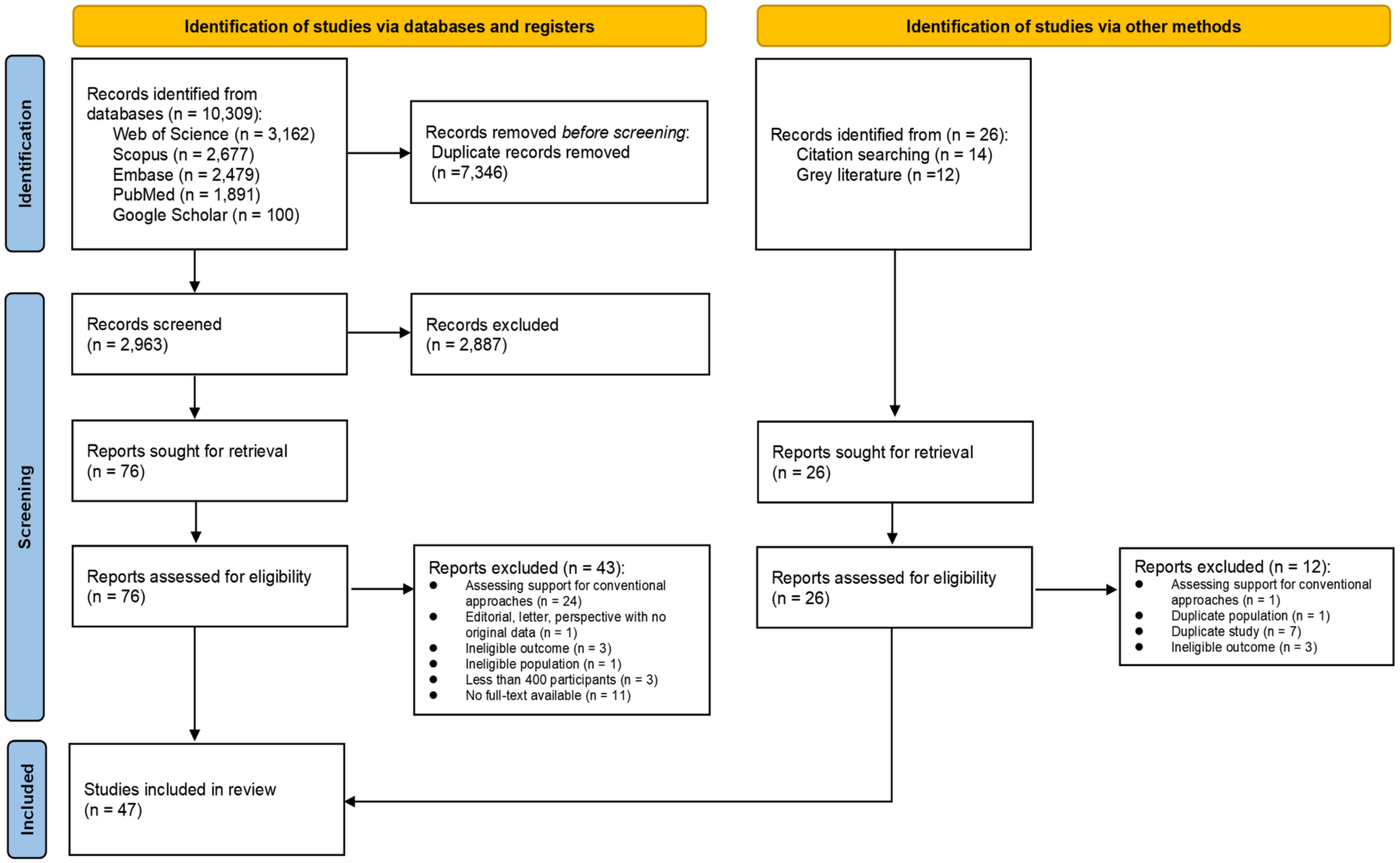

**Figure S3. PRISMA diagram**

### 4. Geographical distribution of included studies in the review

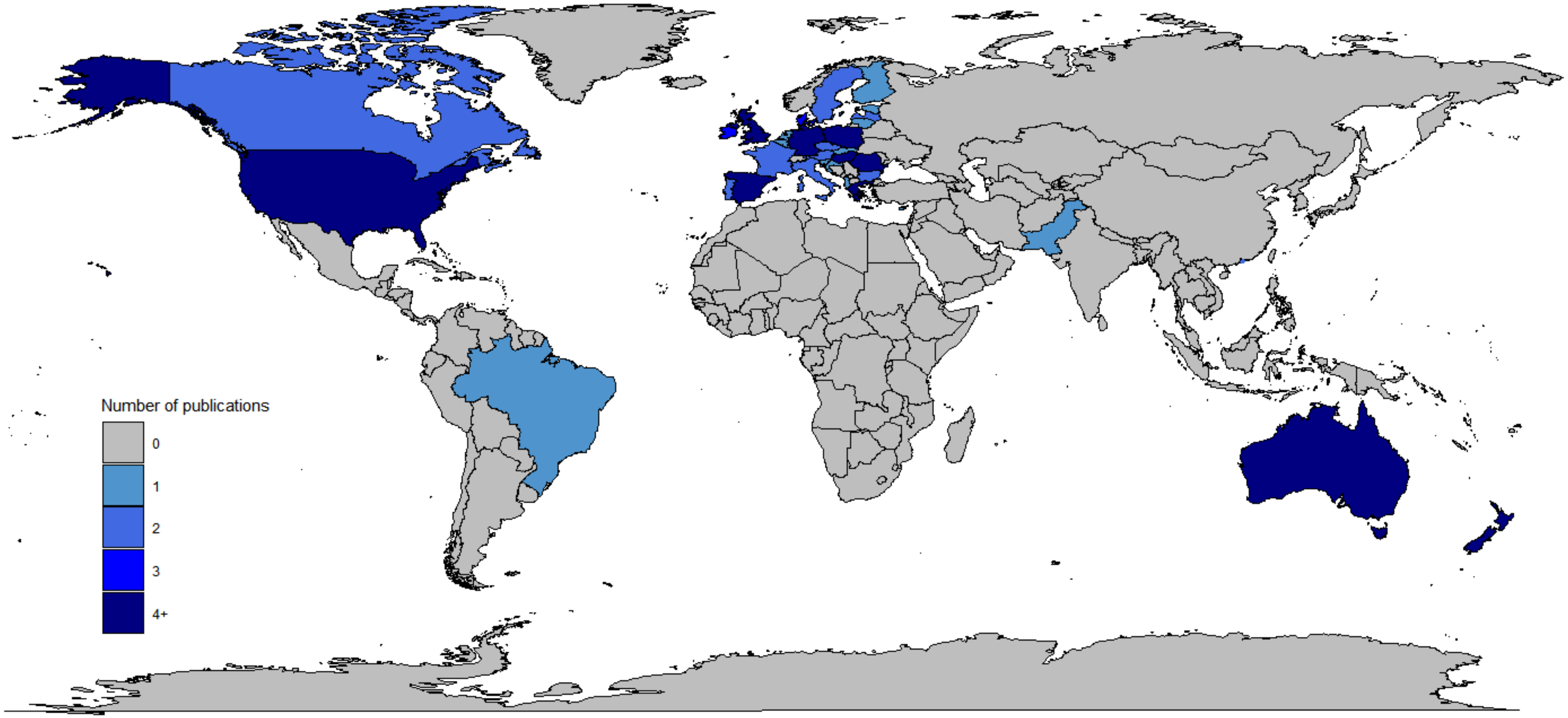

**Figure S4. Countries represented in the review**

### 5. Tobacco endgame policies addressed in the studies

#### Table S5. Policies addressed in each study

| # | Author (year) | Tobacco endgame goal | Product focused | | | User focused | | Market/supply focused | | | | | # of goal/ endgame policies considered |
|---|---|---|---|---|---|---|---|---|---|---|---|---|---|
| | | | Mandate very low Limit nicotine in smoked tobacco products | Set product standards to make products unappealing or intolerable | Move consumers from smoked tobacco products to lower-risk products | Require consumers to obtain a licence or prescription | Restrict tobacco supplies and sales by birth year (tobacco-free generation) | Ban commercial sale of combustible tobacco | Set regularly reducing quota on the volume manufactured or imported ('sinking lid') | Reduce commercial viability of tobacco companies | Increases in tobacco tax to make tobacco products unaffordable | Restrictions retailers to substantially reduce tobacco availability | |
| **Total** | | 7 | 14 | 6 | 1 | 2 | 4 | 24 | 1 | 8 | 3 | 4 | - |
| 1 | Action on Smoking and Health (2017)[27] | | | | | | | | | X | | | 1 |
| 2 | Action on Smoking and Health (2021)[28] | | | | | | | | | X | | | 1 |
| 3 | Action on Smoking and Health (2022)[16] | X | | | | | | | | X | X | | 3 |
| 4 | Ali, Al-Shawaf, Wang, & King (2019)[1] | | X | | | | | | | | | | 1 |
| 5 | Al-Shawaf, Grooms, Mahoney, Lunsford & Kittner (2023)[29] | | | | | | | X | | | | | 1 |
| 6 | Avishai, Ribisl & Sheeran (2023)[30] | | | | | | | X | | | | | 1 |

| # | Author (year) | Tobacco endgame goal | Product focused | | | User focused | | Market/supply focused | | | | | # of goal/ endgame policies considered |
|---|---|---|---|---|---|---|---|---|---|---|---|---|---|
| | | | Mandate very low Limit nicotine in smoked tobacco products | Set product standards to make products unappealing or intolerable | Move consumers from smoked tobacco products to lower-risk products | Require consumers to obtain a licence or prescription | Restrict tobacco supplies and sales by birth year (tobacco-free generation) | Ban commercial sale of combustible tobacco | Set regularly reducing quota on the volume manufactured or imported ('sinking lid') | Reduce commercial viability of tobacco companies | Increases in tobacco tax to make tobacco products unaffordable | Restrictions retailers to substantially reduce tobacco availability | |
| 7 | Boeckmann, Kotz, Shahab, Brown & Kastaun (2018)[11] | | | | | | | X | | X | | | 2 |
| 8 | Bolcic-Jankovic & Biener (2015)[31] | | X | | | | | | | | | | 1 |
| 9 | Brennan, Durkin, Scollo, Swanson & Wakefield (2021)[32] | | | | | | | X | | | | | 1 |
| 10 | Brennan, Ilchenko, Scollo, Durkin & Wakefield (2022)[21] | | | | | | | X | | | | X | 2 |
| 11 | Chung-Hall, Fong, Driezen & Craig (2018)[2] | | X | X | | | | | | | | | 2 |
| 12 | Cosgrave, Blake, Murphy, Sheridan, Doyle & Kavanagh (2023)[4] | X | X | X | | X | X | X | | X | X | X | 9 |

| # | Author (year) | Tobacco endgame goal | Product focused | | | User focused | | Market/supply focused | | | | | # of goal/ endgame policies considered |
|---|---|---|---|---|---|---|---|---|---|---|---|---|---|
| | | | Mandate very low Limit nicotine in smoked tobacco products | Set product standards to make products unappealing or intolerable | Move consumers from smoked tobacco products to lower-risk products | Require consumers to obtain a licence or prescription | Restrict tobacco supplies and sales by birth year (tobacco-free generation) | Ban commercial sale of combustible tobacco | Set regularly reducing quota on the volume manufactured or imported ('sinking lid') | Reduce commercial viability of tobacco companies | Increases in tobacco tax to make tobacco products unaffordable | Restrictions retailers to substantially reduce tobacco availability | |
| 13 | Edwards, Johnson, Stanley, Waa, Ouimet & Fong (2021)[3] | X | X | X | | | | X | | | X | X | 6 |
| 14 | Edwards, Wilson, Peace, Weerasekera, Thomson & Gifford (2013)[33] | | | | | | | X | | | | | 1 |
| 15 | Gallup (2022)[34] | | X | | | | | | | | | | 1 |
| 16 | Gallus, Lugo, Fernandez, Gilmore, Leon, Clancy & La Vecchia (2014)[14] | | | | | | | X | | | | | 1 |
| 17 | Gendall, Hoek & Edwards (2014)[35] | X | | | | | | | | | | | 1 |
| 18 | Gendall, Hoek, Maubach & Edwards (2013)[20] | X | | | | | | X | | | | X | 3 |
| 19 | Hayes, Wakefield & | | | | | | | X | | | | | 1 |

| # | Author (year) | Tobacco endgame goal | Product focused | | | User focused | | Market/supply focused | | | | | # of goal/ endgame policies considered |
|---|---|---|---|---|---|---|---|---|---|---|---|---|---|
| | | | Mandate very low Limit nicotine in smoked tobacco products | Set product standards to make products unappealing or intolerable | Move consumers from smoked tobacco products to lower-risk products | Require consumers to obtain a licence or prescription | Restrict tobacco supplies and sales by birth year (tobacco-free generation) | Ban commercial sale of combustible tobacco | Set regularly reducing quota on the volume manufactured or imported ('sinking lid') | Reduce commercial viability of tobacco companies | Increases in tobacco tax to make tobacco products unaffordable | Restrictions retailers to substantially reduce tobacco availability | |
| | Scollo (2014)[36] | | | | | | | | | | | | |
| 20 | Jaine, Healey, Edwards & Hoek (2015)[22] | X | | | | | | X | | | | | 2 |
| 21 | Kock, Shahab, Moore, Shortt, Pearce & Brown (2022)[12] | | | | | | X | | | | | | 1 |
| 22 | Kulak, Kamper-DeMarco & Kozlowski (2020)[37] | | X | | | | | | | | | | 1 |
| 23 | Kyriakos, Fong, de Abreu Perez, Szklo, Driezen, Quah, Figueiredo & Filippidis (2022)[6] | | | X | | | | | | | | | 1 |
| 24 | Li & Newcombe (2013)[23] | | X | | | | | | | | | | 1 |
| 25 | Li, Newcombe & Walton | | X | | | | | | | | | | 1 |

| # | Author (year) | Tobacco endgame goal | Product focused | | | User focused | | Market/supply focused | | | | | # of goal/ endgame policies considered |
|---|---|---|---|---|---|---|---|---|---|---|---|---|---|
| | | | Mandate very low Limit nicotine in smoked tobacco products | Set product standards to make products unappealing or intolerable | Move consumers from smoked tobacco products to lower-risk products | Require consumers to obtain a licence or prescription | Restrict tobacco supplies and sales by birth year (tobacco-free generation) | Ban commercial sale of combustible tobacco | Set regularly reducing quota on the volume manufactured or imported ('sinking lid') | Reduce commercial viability of tobacco companies | Increases in tobacco tax to make tobacco products unaffordable | Restrictions retailers to substantially reduce tobacco availability | |
| | (2016)[38] | | | | | | | | | | | | |
| 26 | Lykke, Pisinger & Glümer (2016)[18] | | | | | | | X | | | | | 1 |
| 27 | Moodie, Sinclair, Mackintosh, Power & Bauld (2016)[17] | | | | | | | | | X | | | 1 |
| 28 | Morphett, Puljević, Borland, Carter, Hall & Gartner (2021)[10] | | | | X | | | | | | | | 1 |
| 29 | Newcombe & Li (2013)[24] | | | | | | | X | | | | | 1 |
| 30 | Nogueira, Driezen, Fu, Hitchman, Tigova, Castellano, Kyriakos, Zatonski, Mons, Quah, Demjén, Trofor, Przewozniak, Katsaounou, Fong, | | | | | | | X | | X | | | 2 |

| # | Author (year) | Tobacco endgame goal | Product focused | | | User focused | | Market/supply focused | | | | | # of goal/ endgame policies considered |
|---|---|---|---|---|---|---|---|---|---|---|---|---|---|
| | | | Mandate very low Limit nicotine in smoked tobacco products | Set product standards to make products unappealing or intolerable | Move consumers from smoked tobacco products to lower-risk products | Require consumers to obtain a licence or prescription | Restrict tobacco supplies and sales by birth year (tobacco-free generation) | Ban commercial sale of combustible tobacco | Set regularly reducing quota on the volume manufactured or imported ('sinking lid') | Reduce commercial viability of tobacco companies | Increases in tobacco tax to make tobacco products unaffordable | Restrictions retailers to substantially reduce tobacco availability | |
| | Cardavas, Fernández & EUREST-PLUS Consortium (2022)[13] | | | | | | | | | | | | |
| 31 | Palladino, Hone & Filippidis (2018)[39] | | | | | | | X | | | | | 1 |
| 32 | Patel, Cuccia, Zhou, Czaplicki, Pitzer, Hair, Schillo & Vallone (2019)[40] | | X | | | | | | | | | | 1 |
| 33 | Pearson, Abrams, Niaura, Richardson, & Vallone (2013)[5] | | X | | | | | | | | | | 1 |
| 34 | Pepper, Squiers, Bann, Coglaiti & McCormack (2020)[41] | | X | | | | | | | | | | 1 |
| 35 | Robertson, Gendall, Hoek, | X | | | | | | | | | | | 1 |

| # | Author (year) | Tobacco endgame goal | Product focused | | | User focused | | Market/supply focused | | | | | # of goal/ endgame policies considered |
|---|---|---|---|---|---|---|---|---|---|---|---|---|---|
| | | | **Mandate very low Limit nicotine in smoked tobacco products** | **Set product standards to make products unappealing or intolerable** | **Move consumers from smoked tobacco products to lower-risk products** | **Require consumers to obtain a licence or prescription** | **Restrict tobacco supplies and sales by birth year (tobacco-free generation)** | **Ban commercial sale of combustible tobacco** | **Set regularly reducing quota on the volume manufactured or imported (’sinking lid’)** | **Reduce commercial viability of tobacco companies** | **Increases in tobacco tax to make tobacco products unaffordable** | **Restrictions retailers to substantially reduce tobacco availability** | |
| | Cameron, Marsh & McGee (2016)[42] | | | | | | | | | | | | |
| 36 | Schmidt, Kowitt, Myers & Goldstein (2018)[43] | | X | | | | | | | | | | 1 |
| 37 | Siddiqi, Siddiqui, Boeckmann, Islam, Khan, Dobbie, Khan & Kannan (2022)[44] | | | | | | | X | | X | | | 2 |
| 38 | Smith, Nahhas, Borland, Cho, Chung-Hall, Fairman, Fong, McNeill, Popova, Thrasher & Cummings (2021)[7] | | X | X | | | | | | | | | 2 |
| 39 | Sonnenberg, Bostic & Halpern-Felsher (2020)[26] | | | | | | | X | | | | | 1 |

| # | Author (year) | Tobacco endgame goal | Product focused | | | User focused | | Market/supply focused | | | | | # of goal/ endgame policies considered |
|---|---|---|---|---|---|---|---|---|---|---|---|---|---|
| | | | **Mandate very low Limit nicotine in smoked tobacco products** | **Set product standards to make products unappealing or intolerable** | **Move consumers from smoked tobacco products to lower-risk products** | **Require consumers to obtain a licence or prescription** | **Restrict tobacco supplies and sales by birth year (tobacco-free generation)** | **Ban commercial sale of combustible tobacco** | **Set regularly reducing quota on the volume manufactured or imported ('sinking lid')** | **Reduce commercial viability of tobacco companies** | **Increases in tobacco tax to make tobacco products unaffordable** | **Restrictions retailers to substantially reduce tobacco availability** | |
| 40 | Toxværd, Pisinger, Lykke & Lau (2023)[45] | | | | | | | X | | | | | 1 |
| 41 | Trainer, Gall, Smith & Terry (2017)[46] | | | | | | X | | | | | | 1 |
| 42 | Wamamili, Gartner & Lawler (2022)[47] | | | | | | | X | | | | | 1 |
| 43 | Wang, Wang, Lam, Viswanath & Chan (2015)[48] | | | | | | | X | | | | | 1 |
| 44 | White (2015)[19] | | | | | | | X | | | | | 1 |
| 45 | White (2013)[49] | | | | | | | X | | | | | 1 |
| 46 | Wu, Wang, Ho, Cheung, Kwong, Lai & Lam (2019)[15] | | | | | X | X | X | X | | | | 4 |
| 47 | Zatoński, Herbeć, Zatoński, Przewoźniak, Janik-Koncewicz, | | | X | | | | | | | | | 1 |

| # | Author (year) | Tobacco endgame goal | Product focused | | | User focused | | Market/supply focused | | | | | # of goal/ endgame policies considered |
|---|---|---|---|---|---|---|---|---|---|---|---|---|---|
| | | | Mandate very low Limit nicotine in smoked tobacco products | Set product standards to make products unappealing or intolerable | Move consumers from smoked tobacco products to lower-risk products | Require consumers to obtain a licence or prescription | Restrict tobacco supplies and sales by birth year (tobacco-free generation) | Ban commercial sale of combustible tobacco | Set regularly reducing quota on the volume manufactured or imported ('sinking lid') | Reduce commercial viability of tobacco companies | Increases in tobacco tax to make tobacco products unaffordable | Restrictions retailers to substantially reduce tobacco availability | |
| | Mons, Fong, Demjén, Tountas, Trofor, Fernández, McNeil, Willemsen, Hummel, Quah, Kyriakos & Vardavas (2018)[50] | | | | | | | | | | | | |

### 6. Descriptions of the included articles by endgame policy

#### Table S6.1. Study characteristics and percentage support by group: tobacco endgame goal (n = 7)

| #* | Authors (Published year) | Study design | Geographic location; Setting (relevant to policy)** | Name of data source (name of panel, if applicable) | Data collection modality (year) | Study participants (%/n of females, AYA and people who smoke); Overall sample size in analysis | Representative -ness of data | Funding, COI | Group (subgroup/type of question asked) | % Support (95% CI, if available) |
|---|---|---|---|---|---|---|---|---|---|---|
| 3 | Action on Smoking and Health (2022) | Cross-sectional | England, UK; Endgame goal established (≤5% by 2030) | 2022 ASH Smokefree survey | Web (2022) | Adults aged 18+ (1770 AYA aged 18-24, 1415 people who smoke) *n* = 10883 | Representative | Survey funded by cancer Research UK | General population | 74.0% |
| | | | | | | | | | People who smoke | 43.0% |
| | | | | | | | | | AYA | 71.0% |
| 12 | Cosgrave, Blake, Murphy, Sheridan, Doyle & Kavanagh (2023) | Cross-sectional | Ireland; Endgame goal established (≤5% by 2025) | NR | Telephone (2022) | People aged 15+ (50.9% females, 15.9% AYA aged 15-24, 13.6% people who smoke tobacco products) *n* = 1000 | Representative | Survey funded by Health Service Executive Tobacco-Free Ireland Programme | General population | 74.6% (71.9-77.3%) |
| 13 | Edwards, Johnson, Stanley, Waa, Ouimet & Fong (2021) | Cross-sectional analysis of a longitudinal cohort | NZ; Endgame goal established (≤5% by 2025) | ITC NZ | Telephone (W1: 2016-2017, W2: 2018) | Adults aged 18+ who currently smoke and who have quit recently (58.1% and 61.0% females, 8.1% and 8.5% AYA aged 18-24, 78.8% and 71.2% people who smoke for W1 and W2) *n* = 1155 (W1) 1020 (W2) | Representative | One author served as an expert witness in litigation against tobacco industry | People who smoke (goal unprompted) | 42.1% (37.1-47.4%) |
| | | | | | | | | | People who smoke (goal prompted) | 50.6% (45.4-55.7%) |
| 17 | Gendall, Hoek & Edwards (2014) | Cross-sectional | NZ; Endgame goal established (≤5% by 2025) | NR (ResearchNow internet panel) | Web (NR) | People who smoke and who do not smoke (335 people who smoke daily) *n* = 833 | Representative | NR | General population (goal unexplained) | 56.0% |
| | | | | | | | | | General population (goal explained) | 74.0% |
| | | | | | | | | | People who smoke (goal unexplained) | 19.0% |
| | | | | | | | | | People who smoke (goal explained) | 28.0% |

| #* | Authors (Published year) | Study design | Geographic location; Setting (relevant to policy)** | Name of data source (name of panel, if applicable) | Data collection modality (year) | Study participants (%/n of females, AYA and people who smoke); Overall sample size in analysis | Representative-ness of data | Funding, COI | Group (subgroup/type of question asked) | % Support (95% CI, if available) |
|---|---|---|---|---|---|---|---|---|---|---|
| 18 | Gendall, Hoek, Maubach & Edwards (2013) | Cross-sectional | NZ; Endgame goal established (≤5% by 2025) | NR (ResearchNow internet panel) | Web (2012) | People aged 15+ who smoke and who do not smoke (427 females, 151 AYA aged 15-24, 158 people who smoke) *n* = 828 | Representative | None declared | General population | 79.0% |
| | | | | | | | | | People who smoke | 50.0% |
| 20 | Jaine, Healey, Edwards & Hoek (2015) | Repeated cross-sectional | NZ; Endgame goal established in 2011 (≤5% by 2025) | 2009-2012 ASH Year 10 Snapshot Survey | Face-to-face Interview (2009, 2010, 2011, 2012) | Year 10 students aged 14-15 years (47% females, 6% adolescents who smoke daily) *n* = 25762 ('09) 32605 ('10) 26645 ('11) 28447 ('12) | Representative | Survey funded by NZ Ministry of Health | AYA ('11) | 51.0% (49.7-52.3%) |
| | | | | | | | | | AYA ('12) | 59.0% (57.8-60.2%) |
| | | | | | | | | | AYA who smoke daily ('11) | 9.0% |
| | | | | | | | | | AYA who smoke daily ('12) | 10.0% (8-12%) |
| 35 | Robertson, Gendall, Hoek, Cameron, Marsh & McGee (2016) | Cross-sectional | NZ; Endgame goal established (≤5% by 2025) | NR (ResearchNow internet panel) | Web (NR) | Adults aged 18+ who smoke (54.4% females) *n* = 623 | NR | Funded by NZ Asthma Foundation and NZ Lottery Health | People who smoke | 40.0% |

ASH, Action for Smoking and Health; AYA, adolescents and young adults; COI, conflict of interest; ITC, International Tobacco Control; FDA, Food and Drug Administration; NR, Not reported; NZ, New Zealand; UK, United Kingdom; USA, United States of America

*Numbers correspond to the numbers used in Table S3

**At the time of data collection

### Table S6.2. Study characteristics and percentage support by group: limiting nicotine in smoked tobacco products (n = 14)

| #* | Authors (Published year) | Study design | Geographic location; Setting (relevant to policy)** | Name of data source (Panel, if applicable) | Data collection modality (year) | Study participants (%/n of females, AYA and people who smoke); Overall sample size in analysis | Representative-ness of sample | Funding, COI | Group (subgroup) | % Support (95% CI, if available) |
|---|---|---|---|---|---|---|---|---|---|---|
| 4 | Ali, Al-Shawaf, Wang, & King (2019) | Cross-sectional | USA; FDA released potential product standard lowering nicotine in cigarettes | 2018 SummerStyles (Knowledge Panel) | Web (2018) | Adults aged 18+ (50.5% females, 4.2% AYA, 10.9% people who smoke) *n* = 4037 | Representative | None declared | General population | 81.0% (79.6-82.4%) |
| | | | | | | | | | People who smoke | 80.6% (76.5-84.7%) |
| | | | | | | | | | AYA | 79.3% (73.1-85.6%) |
| 8 | Bolcic-Jankovic & Biener (2015) | Cross-sectional | Indianapolis and Dallas/Fort Worth, USA | NR | Mail (2012) | Adults aged 18-65 (494 females, 189 people who smoke) *n* = 934 | Representative | Survey funded by the Ministry for Innovation, Science, and Research of the German Federal State of North Rhine-Westphalia. One author received grants from Cancer Research UK | General population (reduce immediately + gradually) | 79.1% |
| | | | | | | | | | People who smoke (reduce immediately + gradually) | 76.2% (52.9-99.5%) |
| 11 | Chung-Hall, Fong, Driezen & Craig (2018) | Cross-sectional analysis of a longitudinal cohort | Canada; Endgame goal established (5% by 2035) | 2016 ITC Four Country Smoking and Vaping Survey | Telephone or web (2016) | Adults aged 18+ who smoke (52.8% females, 23.6% AYA) *n* = 3215 | Representative (Canadian people who smoke and use e-cigarettes) | Survey funded by US National Cancer Institute and by Foundation Grant | People who smoke | 70.2% |
| | | | | | | | | | AYA who smoke | 71.9% |

| | | | | | | | | | | |
|---|---|---|---|---|---|---|---|---|---|---|
| 12 | Cosgrave, Blake, Murphy, Sheridan, Doyle & Kavanagh (2023) | Cross-sectional | Ireland; Endgame goal established (≤5% by 2025) | NR | Telephone (2022) | People aged 15+ (50.9% females, 15.9% AYA aged 15-24, 13.6% people who smoke tobacco products) *n* = 1000 | Representative | Survey funded by Health Service Executive Tobacco-Free Ireland Programme | General population | 86.1% (84.0-88.2%) |
| | | | | | | | | | People who smoke | 75.5% |
| 13 | Edwards, Johnson, Stanley, Waa, Ouimet & Fong (2021) | Cross-sectional analysis of a longitudinal cohort | NZ; Endgame goal established (≤5% by 2025) | ITC NZ | Telephone (W1: 2016-2017, W2: 2018) | Adults aged 18+ who currently smoke and who have quit recently (58.1% and 61.0% females, 8.1% and 8.5% AYA aged 18-24, 78.8% and 71.2% people who smoke for W1 and W2) *n* = 1155 (W1) 1020 (W2) | Representative | One author served as an expert witness in litigation against tobacco industry | People who smoke | 71.5% (66.5-76.0%) |
| 15 | Gallup (2022) | Cross-sectional | USA and District of Columbia; FDA released potential product standard lowering nicotine in cigarettes | Gallup Poll Social Survey | Telephone (2022) | Adults aged 18+ (516 females, 103 people who smoke) *n* = 1013 | Representative | NR | General population | 74% |
| 22 | Kulak, Kamper-DeMarco & Kozlowski (2020) | Cross-sectional | USA; FDA released potential product standard lowering nicotine in cigarettes | NR (Prime Panel) | Web (2019) | US citizens aged 18+ (61.3% females, 18.7% AYA aged 18-24, 30.7% people who smoke) *n* = 540 | Not representative | None declared | General population (Likert-type response) | 66.1% |
| | | | | | | | | | General population (forced-choice response) | 46.5% |
| | | | | | | | | | People who smoke (Likert-type response) | 51.8% |
| | | | | | | | | | People who smoke (forced-choice response) | 30.1% |
| 24 | Li & Newcombe (2013) | Cross-sectional | NZ; Endgame goal established (≤5% by | 2012 Health and Lifestyles Survey | Face-to-face interview (2012) | People who smoke aged 15+ *n* = 659 | Representative | Survey funded by NZ Ministry of Health | People who smoke (quit-attempters) | 78.1% (69.8-86.4%) |
| | | | | | | | | | People who smoke (non-attempters) | 56.3% (46.2-66.4%) |

| | | | | | | | | | | |
|---|---|---|---|---|---|---|---|---|---|---|
| | | | 2025) | | | | | | | |
| 25 | Li, Newcombe & Walton (2016) | Cross-sectional | NZ; Endgame goal established (≤5% by 2025) | 2014 Health and Lifestyles Survey | Face-to-face interview (2014) | Adults age 15+ (52.1% females, 17.2% AYA aged 15-24, 16.2% people who smoke) *n* = 2594 | Representative | Survey funded by NZ Ministry of Health | General population | 80.7% |
| | | | | | | | | | AYA | 47.4% |
| | | | | | | | | | People who smoke | 62.9% |
| 32 | Patel, Cuccia, Zhou, Czaplicki, Pitzer, Hair, Schillo & Vallone (2019) | Cross-sectional | USA; FDA released potential product standard lowering nicotine in cigarettes | NR (AmeriSpeak panel) | Telephone or web (2018) | Adults aged 18-54 who smoke (40% females, 16% AYA aged 18-24) *n* = 854 | Representative | Funded by Truth Initiative | General population | 72.0% |
| 33 | Pearson, Abrams, Niaura, Richardson, & Vallone (2013) | Cross-sectional | USA; FDA issued notice on potential regulation to explore low nicotine standards in cigarettes | NR (Knowledge Panel) | Web (2010) | Adults, mean age 46.4 (51.6% females, 22.2% people who smoke) *n* = 2649 | Representative | One author supported by a National Research Service Award from the National Institute on Drug Abuse | General population | 46.7% (43.6-49.7%) |
| | | | | | | | | | People who smoke | 45.5% (41.8-49.3%) |
| 34 | Pepper, Squiers, Bann, Coglaiti & McCormack (2020) | Cross-sectional | USA; FDA released potential product standard lowering nicotine in cigarettes | NR (ReseachNow ) | Web (2017) | Adults aged 18+ (56.3% females, 46.9% people who smoke) *n* = 1285 | Not representative | Funded by institutional funds provided by Research Triangle Institute International | General population | 77.8% |
| | | | | | | | | | People who smoke | 46.3% |

| | | | | | | | | | | |
|---|---|---|---|---|---|---|---|---|---|---|
| 36 | Schmidt, Kowitt, Myers & Goldstein (2018) | Cross-sectional | USA; FDA issued notice on potential regulation to explore low nicotine standards in cigarettes | Center for Regulatory Research on Tobacco Communication Survey | Telephone (2014-2015) | Adults aged 18+ (51.1% females, 17.6% people who smoke) *n* = 4337 | Representative | Funded by National Cancer Institute and FDA CTP | General population | 71.0% |
| 38 | Smith, Nahhas, Borland, Cho, Chung-Hall, Fairman, Fong, McNeill, Popova, Thrasher & Cummings (2021) | Cross-sectional | Canada, USA, UK, Australia; Endgame goal established in Canada, UK, Australia | 2016 ITC Four Country Smoking and Vaping Survey | Web (2016) | Adults age 18+ who smoke daily (44.5% females, 11.6% AYA aged 18-24) *n* = 2220 (Canada) 1850 (USA) 2880 (England) 1215 (Australia) | Representative | Funded by National Cancer Institute and FDA CTP | People who smoke (Canada) | 68.0% |
| | | | | | | | | | People who smoke (USA) | 53.8% |
| | | | | | | | | | People who smoke (England) | 56.9% |
| | | | | | | | | | People who smoke (Australia) | 56.7% |

AYA: adolescents and young adults; COI, conflict of interest; CTP, Center for Tobacco Products; ITC, International Tobacco Control; FDA, Food and Drug Administration; NR, Not reported; NZ, New Zealand; UK, United Kingdom; USA, United States of America
*Numbers correspond to the numbers used in Table S3
**At the time of data collection

### Table S6.3. Study characteristics and percentage support by group: standards to make smoked tobacco products unappealing or intolerable (n = 6)

| #* | Authors (Published year) | Study design | Geographic location; Setting (relevant to policy)** | Name of data source (Panel, if applicable) | Data collection modality (year) | Study participants (%/n of females, AYA and people who smoke); Overall sample size in analysis | Representative-ness of sample | Funding, COI | Group (subgroup) | % Support (95% CI, if available) |
|---|---|---|---|---|---|---|---|---|---|---|
| **Banning all additives, including flavourings (n = 6)** | | | | | | | | | | |
| 11 | Chung-Hall, Fong, Driezen & Craig (2018) | Cross-sectional analysis of a longitudinal cohort | Canada; Endgame goal established (5% by 2035) | 2016 ITC Four Country Smoking and Vaping Survey | Telephone or web (2016) | Adults aged 18+ who smoke (52.8% females, 23.6% AYA aged 18-24) *n* = 3215 | Representative (Canadian people who smoke and use e-cigarettes) | Survey funded by US National Cancer Institute and by Foundation Grant | People who smoke | 42.5% |
| | | | | | | | | | AYA who smoke | 36.7% |
| 12 | Cosgrave, Blake, Murphy, Sheridan, Doyle & Kavanagh (2023) | Cross-sectional | Ireland; Endgame goal established (≤5% by 2025) | NR | Telephone (2022) | People aged 15+ (50.9% females, 15.9% AYA aged 15-24, 13.6% people who smoke tobacco products) *n* = 1000 | Representative | Survey funded by Health Service Executive Tobacco-Free Ireland Programme | General population | 69.2% (66.3-72.1%) |
| | | | | | | | | | People who smoke | 58.2% |
| 13 | Edwards, Johnson, Stanley, Waa, Ouimet & Fong (2021) | Cross-sectional analysis of a longitudinal cohort | NZ; Endgame goal established (≤5% by 2025) | ITC NZ | Telephone (W1: 2016-2017, W2: 2018) | Adults aged 18+ who currently smoke and who have quit recently (58.1% and 61.0% females, 8.1% and 8.5% AYA aged 18-24, 78.8% and 71.2% people who smoke for W1 and W2) *n* = 1155 (W1) 1020 (W2) | Representative | One author served as an expert witness in litigation against tobacco industry | People who smoke | 49.1% (44.0-54.3%) |
| 23 | Kyriakos, Fong, de Abreu Perez, Szklo, Driezen, Quah, Figueiredo & Filippidis (2022) | Cross-sectional analysis of a longitudinal cohort | Rio de Janeiro, São Paulo, and Porto Alegre city of Brazil; Additive ban approved as a national policy but yet to be implemented | ITC Brazil | Telephone (2016-2017) | Adults aged 18+ who smoke (52.1% females, 4.4% AYA aged 18-24) *n* = 1216 | Representative | Authors funded by Imperial College London, Canadian Institute of Health Research, Ontario | People who smoke | 61.7% (57.5-65.8%) |
| | | | | | | | | | AYA who smoke | 52.8% |

| | | | | | | | | | | |
|---|---|---|---|---|---|---|---|---|---|---|
| | | | | | | | | Institute for Cancer Research, and Canadian Cancer Society | | |
| 38 | Smith, Nahhas, Borland, Cho, Chung-Hall, Fairman, Fong, McNeill, Popova, Thrasher & Cummings (2021) | Cross-sectional analysis of a longitudinal cohort | Canada, USA, UK, Australia; Endgame goal established in Canada, UK, Australia | 2016 ITC Four Country Smoking and Vaping Survey | Web (2016) | Adults age 18+ who smoke daily<br>(44.5% females, 11.6% AYA aged 18-24)<br>*n* = 2220 (Canada)<br>1850 (USA)<br>2880 (England)<br>1215 (Australia) | Representative | Funded by National Cancer Institute and FDA CTP | People who smoke (Canada) | 40.3% |
| | | | | | | | | | People who smoke (USA) | 34.5% |
| | | | | | | | | | People who smoke (England) | 29.7% |
| | | | | | | | | | People who smoke (Australia) | 34.3% |
| 47 | Zatoński, Herbeć, Zatoński, Przewoźniak, Janik-Koncewicz, Mons, Fong, Demjén, Tountas, Trofor, Fernández, McNeil, Willemsen, Hummel, Quah, Kyriakos & Vardavas (2018) | Cross-sectional | Germany, Greece, Hungary, Poland, Romania, Spain, NL and England | 2016 ITC 6E Survey, 2016 ITC Netherlands Survey and 2016 ITC Four Country Smoking and Vaping Survey (England only) | Face-to-face interview (6E) or web (NL, England) (2016) | Adults aged 18+ who smoke (6E, England) and people aged 15+ who smoke (NL)<br>(44.6% females, 12.4% AYA aged 18-24)<br>*n* = 10760 | Representative | Funded by European Union's Horizon 2020 | People who smoke | 46.3% (44.8-47.8%) |
| **Banning filters (n = 1)** | | | | | | | | | | |
| 12 | Cosgrave, Blake, Murphy, Sheridan, Doyle & Kavanagh (2023) | Cross-sectional | Ireland; Endgame goal established (≤5% by 2025) | NR | Telephone (2022) | People aged 15+<br>(50.9% females, 15.9% AYA aged 15-24, 13.6% people who smoke tobacco products)<br>*n* = 1000 | Representative | Survey funded by Health Service Executive Tobacco-Free Ireland Programme | General population | 51.3% (48.2-54.4%) |
| | | | | | | | | | People who smoke | 35.8% |

AYA, adolescents and young adults; ITC, International Tobacco Control; FDA, Food and Drug Administration; NL, Netherlands; NR, Not reported; NZ, New Zealand; UK, United Kingdom; USA, United States of America; 6E: 6 European Country

*Numbers correspond to the numbers used in Table S3

**At the time of data collection

### Table S6.4. Study characteristics and percentage support by group: transition from smoked tobacco products to lower-risk products (n = 1)

| #* | Authors (Published year) | Study design | Geographic location; Setting (relevant to policy)** | Name of data source (Panel, if applicable) | Data collection modality (year) | Study participants (%/n of females, AYA and people who smoke); Overall sample size in analysis | Representative ness of sample | Funding, COI | Group (subgroup) | % Support (95% CI, if available) |
|---|---|---|---|---|---|---|---|---|---|---|
| 28 | Morphett, Puljević, Borland, Carter, Hall & Gartner (2021) | Cross-sectional | Australia | NR | Web (2015) | Adults aged 18+ who smoke daily (46% females) *n* = 1538 | Representative | Funded by Australia Research Council | People who smoke | 66.2% |

COI, conflict of interests; NR, Not reported
*Numbers correspond to the numbers used in Table S3
**At the time of data collection

### Table S6.5. Study characteristics and percentage support by group: require consumers to obtain a license or prescription (n = 2)

| #* | Authors (Published year) | Study design | Geographic location; Setting (relevant to policy)* | Name of data source (Panel, if applicable) | Data collection modality (year) | Study participants (%/n of females, AYA and people who smoke); Overall sample size in analysis | Representative ness of sample | Funding, COI | Group (subgroup) | % Support (95% CI, if available) |
|---|---|---|---|---|---|---|---|---|---|---|
| 12 | Cosgrave, Blake, Murphy, Sheridan, Doyle & Kavanagh (2023) | Cross-sectional | Ireland; Endgame goal established (≤5% by 2025) | NR | Telephone (2022) | People aged 15+ (50.9% females, 15.9% AYA aged 15-24, 13.6% people who smoke tobacco products) *n* = 1000 | Representative | Survey funded by Health Service Executive Tobacco-Free Ireland Programme | General population | 40.3% (37.3-43.4%) |
| | | | | | | | | | People who smoke | 30.0% |
| 46 | Wu, Wang, Ho, Cheung, Kwong, Lai & Lam (2019) | Cross-sectional | Hong Kong; | Tobacco Control Policy-related Survey 2015 | Telephone (2015) | Adults aged 15+ (52.2% females, 11.5% people who smoke) *n* = 2004 | Representative | Funded by Hong Kong Council on Smoking and Health | General population | 54.1% |

COI, conflict of interests; NR, Not reported
*Numbers correspond to the numbers used in Table S3
**At the time of data collection

### Table S6.6. Study characteristics and percentage support by group: restrict tobacco supplies and sales by birth year (tobacco-free generation) (n = 4)

| #* | Authors (Published year) | Study design | Geographic location; Setting (relevant to policy)* | Name of data source (Panel, if applicable) | Data collection modality (year) | Study participants (%/n of females, AYA and people who smoke); Overall sample size in analysis | Representative ness of sample | Funding, COI | Group (subgroup) | % Support (95% CI, if available) |
|---|---|---|---|---|---|---|---|---|---|---|
| 12 | Cosgrave, Blake, Murphy, Sheridan, Doyle & Kavanagh (2023) | Cross-sectional | Ireland; Endgame goal established (≤5% by 2025) | NR | Telephone (2022) | People aged 15+ (50.9% females, 15.9% AYA aged 15-24, 13.6% people who smoke tobacco products) *n* = 1000 | Representative | Survey funded by Health Service Executive Tobacco-Free Ireland Programme | General population | 56.0% (52.9-59.1%) |
| | | | | | | | | | People who smoke | 39.1% |
| 21 | Kock, Shahab, Moore, Shortt, Pearce & Brown (2022) | Cross-sectional | Great Britain (Scotland, Wales and England), UK; Government outlined an endgame goal | Smoking Toolkit Study | Telephone (2021) | Adults aged 18+ (50% females, 11% AYA aged 18-24, 15% people who smoke) *n* = 2197 | Representative | Authors supported by UK Prevention Research Partnership | General population | 34.5% |
| | | | | | | | | | People who smoke | 21.0% |
| 41 | Trainer, Gall, Smith & Terry (2017) | Cross-sectional | Tasmania, Australia | Tasmanian Smoking and Health Survey (TSHS) and Australian Secondary Students' Alcohol and Drug Survey (ASSAD) | TSHS: telephone (2014) ASSAD: paper (2014) | TSHS: Tasmanians aged 18+ (292 females, 18% people who smoke) *n* = 581 ASSAD: Tasmanian secondary school students aged 12-17 (865 females, 8% people who smoke) *n* = 1743 | Representative | Survey funded by Primary Health Tasmania and Cancer Council Tasmania. Author supported by National Heart Foundation | General population | 73% (69-77%) |
| | | | | | | | | | People who smoke | 71% (63-79%) |
| | | | | | | | | | AYA | 68% (66-70%) |

| #* | Authors (Published year) | Study design | Geographic location; Setting (relevant to policy)* | Name of data source (Panel, if applicable) | Data collection modality (year) | Study participants (%/n of females, AYA and people who smoke); Overall sample size in analysis | Representativeness of sample | Funding, COI | Group (subgroup) | % Support (95% CI, if available) |
|---|---|---|---|---|---|---|---|---|---|---|
| | | | | | | | | | AYA who smoke | 46% (38-54%) |
| 46 | Wu, Wang, Ho, Cheung, Kwong, Lai & Lam (2019) | Cross-sectional | Hong Kong; | Tobacco Control Policy-related Survey 2015 | Telephone (2015) | Adults aged 15+ (52.2% females, 11.5% people who smoke) *n* = 2004 | Representative | Funded by Hong Kong Council on Smoking and Health | General population | 51.8% |

AYA, adolescents and young adults; COI, conflict of interests; NR, Not reported; UK, United Kingdom
*Numbers correspond to the numbers used in Table S3
**At the time of data collection

### Table S6.7. Study characteristics and percentage support by group: ban commercial sales of combustible tobacco (n = 24)

| #* | Authors (Published year) | Study design | Geographic location; Setting (relevant to policy)* | Name of data source (Panel, if applicable) | Data collection modality (year) | Study participants (%/n of females, AYA and people who smoke); Overall sample size in analysis | Representativeness of sample | Funding, COI | Group (subgroup) | % Support (95% CI, if available) |
|---|---|---|---|---|---|---|---|---|---|---|
| 5 | Al-Shawaf, Grooms, Mahoney, Lunsford & Kittner (2023) | Cross-sectional | USA; | 2021 SpringStyles | Web (2021) | Adults aged 18+ (51.6% females, 11.0% people who smoke) *n* = 6455 | Representative | Authors affiliated with the US Office on Smoking and Health at the Centers for Disease Control and Prevention | General population | 57.3% (55.9-58.8%) |
| | | | | | | | | | People who smoke | 25.2 (21.1-29.3%) |
| 6 | Avishai, Ribisl & Sheeran (2023) | Cross-sectional | USA; | NR | Web (NR) | People who smoke, previously smoked and never smoked (57% females, 35.1% people who smoke) *n* = 479 | Not representative | NR | General population | 52.4% |
| | | | | | | | | | People who smoke | 36.3% |
| 7 | Boeckmann, Kotz, Shahab, Brown & Kastaun (2018) | Cross-sectional | Germany; | DEutsche Befragung zum RAuchverhalten (DEBRA) | Face-to-face interviews (2016) | People aged 14+ (51.9% females, 28.4% people who smoke) *n* = 2062 | Representative | One author received funding from Cancer Research UK | General population | 22.9% (21.1-24.8%) |
| | | | | | | | | | People who smoke | 9.9% (7.4-12.5%) |
| 9 | Brennan, Durkin, Scollo, Swanson & Wakefield (2021) | Cross-sectional | Victoria, Australia; | Victorian Smoking and Health Survey | Telephone (2019) | Adults aged 18+ (31.7% people who smoke) *n* = 2774 | Representative | Authors received funding from VicHealth, the Victorian Department of Health and Human Services, Cancer | General population (Likert) | 52.8% |
| | | | | | | | | | General population (Timeframe) | 79.5% |
| | | | | | | | | | People who smoke (Likert) | 31.7% |

| #* | Authors (Published year) | Study design | Geographic location; Setting (relevant to policy)* | Name of data source (Panel, if applicable) | Data collection modality (year) | Study participants (%/n of females, AYA and people who smoke); Overall sample size in analysis | Representativeness of sample | Funding, COI | Group (subgroup) | % Support (95% CI, if available) |
|---|---|---|---|---|---|---|---|---|---|---|
| | | | | | | | | Council Victoria | People who smoke (Timeframe) | 63.1% |
| 10 | Brennan, Ilchenko, Scollo, Durkin & Wakefield (2022) | Cross-sectional | Australia | NR (Life in Australia) | Web or telephone (2019) | Adults aged 18+ (950 females, 282 people who smoke) *n* = 1874 | Representative | Funded by Cancer Council Victoria | General population | 61.6% |
| | | | | | | | | | People who smoke | 39.2% |
| 12 | Cosgrave, Blake, Murphy, Sheridan, Doyle & Kavanagh (2023) | Cross-sectional | Ireland; Endgame goal established (≤5% by 2025) | NR | Telephone (2022) | People aged 15+ (50.9% females, 15.9% AYA aged 15-24, 13.6% people who smoke tobacco products) *n* = 1000 | Representative | Survey funded by Health Service Executive Tobacco-Free Ireland Programme | General population | 82.8% (80.5-85.1%) |
| | | | | | | | | | People who smoke | 66.4% |
| 13 | Edwards, Johnson, Stanley, Waa, Ouimet & Fong (2021) | Cross-sectional analysis of a longitudinal cohort | NZ; Endgame goal established (≤5% by 2025) | ITC NZ | Telephone (W1: 2016-2017, W2: 2018) | Adults aged 18+ who currently smoke and who have quit recently (58.1% and 61.0% females, 8.1% and 8.5% AYA aged 18-24, 78.8% and 71.2% people who smoke for W1 and W2) *n* = 1155 (W1) 1020 (W2) | Representative | One author served as an expert witness in litigation against tobacco industry | People who smoke | 44.9% (40.5-59.4%) |
| 14 | Edwards, Wilson, Peace, Weerasekera, | Cross-sectional analysis of a | NZ | ITC NZ | Telephone (2007-2009) | Adults aged 18+ who currently smoke (61.6% females) | Representative | Survey funded by NZ Health Research | People who smoke (W2) | 46.0% (41.6-50.4%) |

| #* | Authors (Published year) | Study design | Geographic location; Setting (relevant to policy)* | Name of data source (Panel, if applicable) | Data collection modality (year) | Study participants (%/n of females, AYA and people who smoke); Overall sample size in analysis | Representativeness of sample | Funding, COI | Group (subgroup) | % Support (95% CI, if available) |
|---|---|---|---|---|---|---|---|---|---|---|
| | Thomson & Gifford (2013) | longitudinal cohort | | | | *n* = 1376 (W1)<br>923 (W2) | | Council | | |
| 16 | Gallus, Lugo, Fernandez, Gilmore, Leon, Clancy & La Vecchia (2014) | Cross-sectional | 18 European countries (Albania, Austria, Bulgaria, the Czech Republic, Croatia, England, Finland, France, Greece, Hungary, Ireland, Italy, Latvia, Poland, Portugal, Romania, Spain and Sweden); | Pricing policies and Control of Tobacco in Europe (PPACTE) | Face-to-face interviews (2010) | Adults aged 15+ (8765 females, 2651 AYA aged 15-24, 5019 people who smoke)<br>*n* = 16947 | Representative | Funded by the European Commission Seventh Framework Programme Grant. Some authors are supported by National Cancer Institute | General population | 34.9% |
| | | | | | | | | | People who smoke | 25.6% |
| | | | | | | | | | AYA | 37.8% |
| 18 | Gendall, Hoek, Maubach & Edwards (2013) | Cross-sectional | NZ;<br>Endgame goal established (≤5% by 2025) | NR (ResearchNow internet panel) | Web (2012) | People aged 15+ who smoke and who do not smoke (427 females, 151 AYA aged 15-24, 158 people who smoke)<br>*n* = 828 | Representative | None declared | General population | 50.0% |
| | | | | | | | | | People who smoke | 18.0% |
| 19 | Hayes, Wakefield & Scollo (2014) | Cross-sectional | Victoria, Australia | Victorian Smoking and Health Survey | Telephone (2009, 2010) | Adults aged 18+<br>*n* = 4502 ('09)<br>4499 ('10) | Representative | Funded by Quit Victoria | General population ('09) | 71.6% |
| | | | | | | | | | General population ('10, with time frame) | 70.9% |

| #* | Authors (Published year) | Study design | Geographic location; Setting (relevant to policy)* | Name of data source (Panel, if applicable) | Data collection modality (year) | Study participants (%/n of females, AYA and people who smoke); Overall sample size in analysis | Representativeness of sample | Funding, COI | Group (subgroup) | % Support (95% CI, if available) |
|---|---|---|---|---|---|---|---|---|---|---|
| | | | | | | | | | People who smoke ('09) | 57.0% |
| | | | | | | | | | People who smoke ('10, with time frame) | 42.2% |
| 20 | Jaine, Healey, Edwards & Hoek (2015) | Repeated cross-sectional | NZ;<br>Endgame goal established in 2011 (≤5% by 2025) | 2009-2012 ASH Year 10 Snapshot Survey | Face-to-face Interview (2009, 2010, 2011, 2012) | Year 10 students aged 14-15 years (49% females, 5.5% adolescents who smoke daily)<br>*n* = 25762 ('09)<br>32605 ('10)<br>26645 ('11)<br>28447 ('12) | Representative | Survey funded by NZ Ministry of Health | AYA ('10) | 48% (47.0-49.0%) |
| | | | | | | | | | AYA ('11) | 52% |
| | | | | | | | | | AYA ('12) | 57% (55.8-58.2%) |
| | | | | | | | | | AYA who smoke ('10) | 11% |
| | | | | | | | | | AYA who smoke ('11) | 13% |
| | | | | | | | | | AYA who smoke ('12) | 13% (10.6-15.4%) |
| 26 | Lykke, Pisinger & Glümer (2016) | Cross-sectional | København, Denmark; | How Are You? Survey 2013 | Mail (2013) | Adults aged 16+ (51.6% females, 14.3% AYA aged 16-24, 15.5% people who smoke daily)<br>*n* = 41356 | Representative | Funded by Capital Region of Denmark | General population | 30.6% |
| | | | | | | | | | People who smoke | 18.2% (16.9-19.5%) |
| | | | | | | | | | AYA | 34.0% (32.3-35.7%) |
| 29 | Newcombe & Li (2013) | Cross-sectional | NZ; | 2012 Health and Lifestyles Survey | Face-to-face interview (2012) | Adults aged 15+<br>*n* = 2672 | Representative | Survey funded by NZ Ministry of Health | General population | 54% (50-57%) |
| | | | | | | | | | People who smoke | 34.0% |
| 30 | Nogueira, Driezen, Fu, Hitchman, Tigova, Castellano, Kyriakos, Zatonski, Mons, Quah, | Cross-sectional analyses of a longitudinal sample | Germany, Greece, Hungary, Poland, Romania, Spain | ITC 6E Survey | Face-to-face interview (2016, 2018) | Adults aged 18+ who smoke (in W1)<br>*n* = 6011 (W1)<br>6027 (W2) | Representative | Funded by European Union's Horizon 2020 research and innovation programme | People who smoke (W2) | 37.8% (35.3-40.4%) |

| #* | Authors (Published year) | Study design | Geographic location; Setting (relevant to policy)* | Name of data source (Panel, if applicable) | Data collection modality (year) | Study participants (%/n of females, AYA and people who smoke); Overall sample size in analysis | Representativeness of sample | Funding, COI | Group (subgroup) | % Support (95% CI, if available) |
|---|---|---|---|---|---|---|---|---|---|---|
| | Demjen, Trofor, Przewozniak, Katsaounou, Fong, Cardavas, Fernandez & EUREST-PLUS Consortium (2022) | | | | | | | | AYA who smoke (W2) | 36.7% |
| 31 | Palladino, Hone, Filippidis (2018) | Repeated cross-sectional | 27 EU member states (Austria, Belgium, Bulgaria, Croatia, Republic of Cyprus, Czech Republic, Denmark, Estonia, Finland, France, Germany, Greece, Hungary, Ireland, Italy, Latvia, Lithuania, Luxembourg, Malta, Netherlands, | Flash Eurobarometer Survey | Telephone (2008, 2011, 2014) | Adolescents and young adults aged 15-24<br>*n* = 12312 ('08)<br>12313 ('11)<br>12628 ('14) | Representative | Funded by National Institute for Health Research | AYA ('08)*<br>*Study provided estimates for each 27 EU member states. But only the overall is provided here | 17.9%<br>(16.8-19.0%) |
| | | | | | | | | | AYA ('11)*<br>*Study provided estimates for each 27 EU member states. But only the overall is provided here | 16.5%<br>(15.4-17.5%) |

| #* | Authors (Published year) | Study design | Geographic location; Setting (relevant to policy)* | Name of data source (Panel, if applicable) | Data collection modality (year) | Study participants (%/n of females, AYA and people who smoke); Overall sample size in analysis | Representativeness of sample | Funding, COI | Group (subgroup) | % Support (95% CI, if available) |
|---|---|---|---|---|---|---|---|---|---|---|
| | | | Poland, Portugal, Romania, Slovakia, Slovenia, Spain, Sweden) | | | | | | AYA ('14)*<br>*Study provided estimates for each 27 EU member states. But only the overall is provided here | 16.0%<br>(15.0-17.1%) |
| 37 | Siddiqi, Siddiqui, Boeckmann, Islam, Khan, Dobbie, Khan & Kannan (2022) | Cross-sectional | Pakistan | Studying Tobacco Users of Pakistan (STOP) | Face-to-face interview (2019-2020) | People who smoke aged 15+<br>(1.3% females)<br>*n* = 6014 | Representative | Funded by European Union Horizon 2020 Grant | People who smoke | 93.9% |
| 39 | Sonnenberg, Bostic & Halpern-Felsher (2020) | Cross-sectional | California, USA | Tobacco Perceptions Study | Web (2018) | AYA aged 16-23<br>(284 females)<br>*n* = 450 | Not representative | Funded by National Cancer Institute and FDA CTP | AYA<br>(ban sale immediately) | 57.0% |
| | | | | | | | | | AYA<br>(ban sale gradually) | 73.4% |
| 40 | Toxværd, Pisinger, Lykke & Lau (2023) | Repeated cross-sectional | Denmark; Endgame goal established (first smoke-free generation by 2030), but lacked clear definition | Denmark Capital Region Health Survey | Web or paper (2013, 2017) | Adults aged 16+<br>(51.6% females, 14.3% AYA aged 16-24, 15.5% people who smoke daily)<br>*n* = 55185 ('17) | Representative | Funded by Capital Region of Copenhagen | General population | 50.3%<br>(49.2-51.5%) |
| | | | | | | | | | AYA | 51.0%<br>(49.5-52.6%) |
| | | | | | | | | | People who smoke | 27.2%<br>(26.0-28.5%) |

| #* | Authors (Published year) | Study design | Geographic location; Setting (relevant to policy)* | Name of data source (Panel, if applicable) | Data collection modality (year) | Study participants (%/n of females, AYA and people who smoke); Overall sample size in analysis | Representativeness of sample | Funding, COI | Group (subgroup) | % Support (95% CI, if available) |
|---|---|---|---|---|---|---|---|---|---|---|
| 42 | Wamamili, Gartner & Lawler (2022) | Cross-sectional | NZ and Queensland, Australia | NR | Web or paper (NZ: 2018 Australia: 2017) | University students aged 18+ (60.4% and 57.7% females, 68.5% and 82.6% AYA aged 18-24, 8.7% and 10.5% people who smoke daily for UQ and NZ respectively) *n* = 5172 (UQ) 1932 (NZ) | Not representative | None declared | General population (UQ) | 51.6% |
| | | | | | | | | | General population (NZ) | 53.3% |
| 43 | Wang, Wang, Lam, Viswanath & Chan (2015) | Cross-sectional | Hong Kong; | Hong Kong Family and Health Information Trends Survey | Telephone (2012) | Adults aged 18+ (52.1% females, 13.2% AYA aged 18-24, 9.5% people who smoke daily) *n* = 1516 | Representative | Funded by Hong Kong Jockey Club Charities Trust | General population | 71.2% (65.6-76.8%) |
| | | | | | | | | | People who smoke | 50.5% (32.4-68.6%) |
| 44 | White (2015) | Cross-sectional | NZ; Endgame goal established (≤5% by 2025) | 2014 Youth Insights Survey | Paper (2014) | Year 10 students aged 14-15 (8% AYA who smoke daily) *n* = 2919 | Representative | NR | AYA | 56.0% (54.0-58.0%) |
| | | | | | | | | | AYA who smoke | 12.0% |
| 45 | White (2013) | Cross-sectional | NZ; Endgame goal established (≤5% by 2025) | 2012 Youth Insights Survey | Paper (2012) | Year 10 students aged 14-15 (7% AYA who smoke daily) *n* = 3143 | Representative | NR | AYA | 57.0% (55.0-59.0%) |
| | | | | | | | | | AYA who smoke | 11.0% |
| 46 | Wu, Wang, Ho, Cheung, Kwong, Lai & Lam (2019) | Cross-sectional | Hong Kong; | Tobacco Control Policy-related Survey 2015 | Telephone (2015) | Adults aged 15+ (52.2% females, 11.5% people who smoke) *n* = 2004 | Representative | Funded by Hong Kong Council on Smoking and Health | General population | 70.5% |

| #* | Authors (Published year) | Study design | Geographic location; Setting (relevant to policy)* | Name of data source (Panel, if applicable) | Data collection modality (year) | Study participants (%/n of females, AYA and people who smoke); Overall sample size in analysis | Representativeness of sample | Funding, COI | Group (subgroup) | % Support (95% CI, if available) |
|---|---|---|---|---|---|---|---|---|---|---|

AYA, adolescents and young adults; COI, conflict of interests; CTP, Center for Tobacco Products; ITC, International Tobacco Control; FDA, Food and Drug Administration; NL, Netherlands; NR, Not reported; NZ, New Zealand; UK, United Kingdom; UQ, University of Queensland; USA, United States of America; W, Wave; 6E: 6 European Country

*Numbers correspond to the numbers used in Table S3

**At the time of data collection

### Table S6.8. Study characteristics and percentage support by group: regularly reduce the volume manufactured or imported (sinking lid) (n = 1)

| #* | Authors (Published year) | Study design | Geographic location; Setting (relevant to policy)** | Name of data source (Panel, if applicable) | Data collection modality (year) | Study participants (%/n of females, AYA and people who smoke); Overall sample size in analysis | Representativeness of sample | Funding, COI | Group (subgroup) | % Support (95% CI, if available) |
|---|---|---|---|---|---|---|---|---|---|---|
| 46 | Wu, Wang, Ho, Cheung, Kwong, Lai & Lam (2019) | Cross-sectional | Hong Kong; | Tobacco Control Policy-related Survey 2015 | Telephone (2015) | Adults aged 15+ (52.2% females, 11.5% people who smoke) *n* = 2004 | Representative | Funded by Hong Kong Council on Smoking and Health | General population | 80.0% |

COI, conflict of interests
*Numbers correspond to the numbers used in Table S3
**At the time of data collection

### Table S6.9. Study characteristics and percentage support by group: reduce the commercial viability of tobacco companies (n = 8)

| #* | Authors (Published year) | Study design | Geographic location; Setting (relevant to policy)** | Name of data source (Panel, if applicable) | Data collection modality (year) | Study participants (%/n of females, AYA and people who smoke); Overall sample size in analysis | Representativeness of sample | Funding, COI | Group (subgroup) | % Support (95% CI, if available) |
|---|---|---|---|---|---|---|---|---|---|---|
| 1 | Action on Smoking and Health (2017) | Cross-sectional | England, UK; | 2017 ASH Smokefree survey | Web (2017) | Adults aged 18+ (1351 people who smoke) $n$ = 10488 | Representative | Survey funded by cancer Research UK | General population | 71.0% |
| | | | | | | | | | People who smoke | 43.0% |
| 2 | Action on Smoking and Health (2021) | Cross-sectional | England, UK; Endgame goal established (≤5% by 2030) | 2021 ASH Smokefree survey | Web (2021) | Adults aged 18+ (1228 people who smoke) $n$ = 10211 | Representative | Survey funded by cancer Research UK | General population | 77% |
| 3 | Action on Smoking and Health (2022) | Cross-sectional | England, UK; Endgame goal established (≤5% by 2030) | 2022 ASH Smokefree survey | Web (2022) | Adults aged 18+ (1770 AYA aged 18-24, 1415 people who smoke) $n$ = 10883 | Representative | Survey funded by cancer Research UK | General population | 76.0% |
| | | | | | | | | | People who smoke | 50.0% |
| | | | | | | | | | AYA | 71.0% |
| 7 | Boeckmann, Kotz, Shahab, Brown & Kastaun (2018) | Cross-sectional | Germany; | DEutsche Befragung zum RAuchverhalten (DEBRA) | Face-to-face interviews (2016) | People aged 14+ (51.9% females, 28.4% people who smoke) $n$ = 2062 | Representative | One author received funding from Cancer Research UK | General population | 57.3% |
| 12 | Cosgrave, Blake, Murphy, Sheridan, Doyle & | Cross-sectional | Ireland; Endgame goal established (≤5% by 2025) | NR | Telephone (2022) | People aged 15+ (50.9% females, 15.9% AYA aged 15-24, 13.6% people who smoke tobacco products) | Representative | Survey funded by Health Service Executive | General population | 78.4% (75.9-81.0%) |

| #* | Authors (Published year) | Study design | Geographic location; Setting (relevant to policy)** | Name of data source (Panel, if applicable) | Data collection modality (year) | Study participants (%/n of females, AYA and people who smoke); Overall sample size in analysis | Representativeness of sample | Funding, COI | Group (subgroup) | % Support (95% CI, if available) |
|---|---|---|---|---|---|---|---|---|---|---|
| | Kavanagh (2023) | | | | | *n* = 1000 | | Tobacco-Free Ireland Programme | People who smoke | 62.4% |
| 27 | Moodie, Sinclair, Mackintosh, Power & Bauld (2016) | Cross-sectional | UK | NR (YouGov) | Web (2014) | Adults aged 16+ (50.9% females, 14.9% AYA aged 16-24, 19.2% people who smoke tobacco products) *n* = 2253 | Representative | Funded by Cancer Research UK | General population | 46.0% |
| 30 | Nogueira, Driezen, Fu, Hitchman, Tigova, Castellano, Kyriakos, Zatonski, Mons, Quah, Demjen, Trofor, Przewozniak, Katsaounou, Fong, Cardavas, Fernandez & EUREST-PLUS Consortium (2022) | Cross-sectional analyses of a longitudinal sample | Germany, Greece, Hungary, Poland, Romania, Spain | ITC 6E Survey | Face-to-face interview (2016, 2018) | Adults aged 18+ who smoke (in W1) *n* = 6011 (W1) 6027 (W2) | Representative | Funded by European Union Horizon 2020 Grant | General population (W2) | 48.7% (45.9-51.5%) |
| | | | | | | | | | AYA | 54.1% |

| #* | Authors (Published year) | Study design | Geographic location; Setting (relevant to policy)** | Name of data source (Panel, if applicable) | Data collection modality (year) | Study participants (%/n of females, AYA and people who smoke); Overall sample size in analysis | Representativeness of sample | Funding, COI | Group (subgroup) | % Support (95% CI, if available) |
|---|---|---|---|---|---|---|---|---|---|---|
| 37 | Siddiqi, Siddiqui, Boeckmann, Islam, Khan, Dobbie, Khan & Kannan (2022) | Cross-sectional | Pakistan | Studying Tobacco Users of Pakistan (STOP) | Face-to-face interview (2019-2020) | People who smoke aged 15+ (1.5% females) *n* = 6014 | Representative | Funded by European Union Horizon 2020 Grant | People who smoke | 85.9% |

ASH, Action on Smoking and Health; AYA, adolescents and young adults; COI, conflict of interests; ITC, International Tobacco Control; NR, not reported; UK, United Kingdom; W, Wave; 6E: 6 European Country

*Numbers correspond to the numbers used in Table S3

**At the time of data collection

### Table S6.10. Study characteristics and percentage support by group: increase taxes to make tobacco products unaffordable (n = 3)

| #* | Authors (Published year) | Study design | Geographic location; Setting (relevant to policy)** | Name of data source (Panel, if applicable) | Data collection modality (year) | Study participants (%/n of females, AYA and people who smoke); Overall sample size in analysis | Representativeness of sample | Funding, COI | Group (subgroup) | % Support (95% CI, if available) |
|---|---|---|---|---|---|---|---|---|---|---|
| 3 | Action on Smoking and Health (2022) | Cross-sectional | England, UK; Endgame goal established (≤5% by 2030) | 2022 ASH Smokefree survey | Web (2022) | Adults aged 18+ (1770 AYA aged 18-24, 1415 people who smoke) $n$ = 10883 | Representative | Survey funded by cancer Research UK | General population | 63.0% |
| | | | | | | | | | People who smoke | 21.0% |
| | | | | | | | | | AYA | 58.0% |
| 12 | Cosgrave, Blake, Murphy, Sheridan, Doyle & Kavanagh (2023) | Cross-sectional | Ireland; Endgame goal established (≤5% by 2025) | NR | Telephone (2022) | People aged 15+ (50.9% females, 15.9% AYA aged 15-24, 13.6% people who smoke tobacco products) $n$ = 1000 | Representative | Survey funded by Health Service Executive Tobacco-Free Ireland Programme | General population | 59.6% (56.6-62.6%) |
| | | | | | | | | | People who smoke | 26.6% |
| 13 | Edwards, Johnson, Stanley, Waa, Ouimet & Fong (2021) | Cross-sectional analysis of a longitudinal cohort | NZ; Endgame goal established (≤5% by 2025) | ITC NZ | Telephone (W1: 2016-2017, W2: 2018) | Adults aged 18+ who currently smoke and who have quit recently (58.1% and 61.0% females, 8.1% and 8.5% AYA aged 18-24, 78.8% and 71.2% people who smoke for W1 and W2) $n$ = 1155 (W1) 1020 (W2) | Representative | One author served as an expert witness in litigation against tobacco industry | People who smoke | 22.2% (18.3-26.7%) |

ASH, Action on Smoking and Health; AYA, adolescents and young adults; COI, conflict of interests; ITC, International Tobacco Control; NR, not reported; NZ, New Zealand; UK, United Kingdom; W, Wave
*Numbers correspond to the numbers used in Table S3
**At the time of data collection

### Table S6.11. Study characteristics and percentage support by group: restrict retailers to substantially limit availability (n = 4)

| #* | Authors (Published year) | Study design | Geographic location; Setting (relevant to policy)** | Name of data source (Panel, if applicable) | Data collection modality (year) | Study participants (%/n of females, AYA and people who smoke); Overall sample size in analysis | Representativeness of sample | Funding, COI | Group (subgroup) | % Support (95% CI, if available) |
|---|---|---|---|---|---|---|---|---|---|---|
| **Substantially reduce availability or transition out (n = 3)** | | | | | | | | | | |
| 10 | Brennan, Ilchenko, Scollo, Durkin & Wakefield (2022) | Cross-sectional | Australia | NR (Life in Australia) | Web (88%) or telephone (12%) | Adults aged 18+ (950 females, 282 people who smoke) *n* = 1874 | Representative | Funded by Cancer Council Victoria | General population | 64.6% |
| | | | | | | | | | People who smoke | 38.5% |
| 12 | Cosgrave, Blake, Murphy, Sheridan, Doyle & Kavanagh (2023) | Cross-sectional | Ireland; Endgame goal established (≤5% by 2025) | NR | Telephone (2022) | People aged 15+ (50.9% females, 15.9% AYA aged 15-24, 13.6% people who smoke tobacco products) *n* = 1000 | Representative | Survey funded by Health Service Executive Tobacco-Free Ireland Programme | General population | 58.9% (55.9-62.0%) |
| | | | | | | | | | People who smoke | 33.0% |
| 13 | Edwards, Johnson, Stanley, Waa, Ouimet & Fong (2021) | Cross-sectional analysis of a longitudinal cohort | NZ; Endgame goal established (≤5% by 2025) | ITC NZ | Telephone (W1: 2016-2017, W2: 2018) | Adults aged 18+ who currently smoke and who have quit recently (58.1% and 61.0% females, 8.1% and 8.5% AYA aged 18-24, 78.8% and 71.2% people who smoke for W1 and W2) *n* = 1155 (W1) 1020 (W2) | Representative | One author served as an expert witness in litigation against tobacco industry | People who smoke | 40.3% (36.0-44.8%) |
| **Limiting sales to a specific type of store (n = 1)** | | | | | | | | | | |

| #* | Authors (Published year) | Study design | Geographic location; Setting (relevant to policy)** | Name of data source (Panel, if applicable) | Data collection modality (year) | Study participants (%/n of females, AYA and people who smoke); Overall sample size in analysis | Representativeness of sample | Funding, COI | Group (subgroup) | % Support (95% CI, if available) |
|---|---|---|---|---|---|---|---|---|---|---|
| 18 | Gendall, Hoek, Maubach & Edwards (2013) | Cross-sectional | NZ; Endgame goal established (≤5% by 2025) | NR (ResearchNow internet panel) | Web (2012) | People aged 15+ who smoke and who do not smoke (427 females, 151 AYA aged 15-24, 158 people who smoke) *n* = 828 | Representative | None declared | General population | 22% |
| | | | | | | | | | People who smoke | 15% |

COI, conflict of interests; ITC, International Tobacco Control; NR, not reported; NZ, New Zealand; W, Wave;
*Numbers correspond to the numbers used in Table S3
**At the time of data collection

### 7. Quality assessment results

#### Table S7. Quality assessment results of the individual studies

| # | Author (Year) | Q1 | Q2 | Q3 | Q4 | Q5 | Q6 | Q7 | Q8 | Q9 | Total |
|---|---|---|---|---|---|---|---|---|---|---|---|
| 1 | Action on Smoking and Health (2017) | Y | Y | Y | Y | U | Y | Y | N | Y | 7 |
| 2 | Action on Smoking and Health (2021) | Y | Y | Y | Y | U | Y | Y | N | U | 7 |
| 3 | Action on Smoking and Health (2022) | Y | Y | Y | Y | U | Y | Y | N | U | 7 |
| 4 | Ali, Al-Shawaf, Wang, & King (2019) | Y | Y | Y | Y | Y | Y | Y | Y | Y | 9 |
| 5 | Al-Shawaf, Grooms, Mahoney, Lunsford & Kittner (2023) | Y | Y | Y | Y | Y | Y | Y | Y | Y | 9 |
| 6 | Avishai, Ribisl & Sheeran (2023) | Y | N | Y | Y | NA | Y | Y | N | NA | 5 |
| 7 | Boeckmann, Kotz, Shahab, Brown & Kastaun (2018) | Y | Y | Y | Y | Y | Y | Y | N | Y | 9 |
| 8 | Bolcic-Jankovic & Biener (2015) | Y | Y | Y | Y | Y | Y | Y | Y | Y | 9 |
| 9 | Brennan, Durkin, Scollo, Swanson & Wakefield (2021) | Y | Y | Y | Y | Y | Y | Y | N | Y | 8 |
| 10 | Brennan, Ilchenko, Scollo, Durkin & Wakefield (2022) | Y | Y | Y | Y | Y | Y | Y | N | Y | 8 |
| 11 | Chung-Hall, Fong, Driezen & Craig (2018) | Y | Y | Y | Y | Y | Y | Y | Y | Y | 9 |
| 12 | Cosgrave, Blake, Murphy, Sheridan, Doyle & Kavanagh (2023) | Y | Y | Y | Y | Y | Y | Y | Y | Y | 9 |
| 13 | Edwards, Johnson, Stanley, Waa, Ouimet & Fong (2021) | Y | Y | Y | Y | Y | Y | Y | Y | Y | 9 |
| 14 | Edwards, Wilson, Peace, Weerasekera, Thomson & Gifford (2013) | Y | Y | Y | Y | Y | Y | Y | N | Y | 8 |
| 15 | Gallup (2022) | Y | Y | Y | Y | U | Y | Y | N | Y | 7 |
| 16 | Gallus, Lugo, Fernandez, Gilmore, Leon, Clancy & La Vecchia (2014) | Y | Y | Y | Y | Y | Y | Y | N | Y | 8 |
| 17 | Gendall, Hoek & Edwards (2014) | Y | Y | Y | Y | U | Y | Y | N | Y | 7 |
| 18 | Gendall, Hoek, Maubach & Edwards (2013) | Y | Y | Y | Y | U | Y | Y | N | Y | 7 |
| 19 | Hayes, Wakefield & Scollo (2014) | Y | Y | Y | Y | Y | Y | Y | N | Y | 8 |
| 20 | Jaine, Healey, Edwards & Hoek (2015) | Y | Y | Y | Y | Y | Y | Y | Y | Y | 9 |
| 21 | Kock, Shahab, Moore, Shortt, Pearce & Brown (2022) | Y | Y | Y | Y | U | Y | Y | N | Y | 7 |
| 22 | Kulak, Kamper-DeMarco & Kozlowski (2020) | Y | Y | N | Y | Y | Y | Y | Y | N | 7 |
| 23 | Kyriakos, Fong, de Abreu Perez, Szklo, Driezen, Quah, Figueiredo & Filippidis (2022) | Y | Y | Y | Y | Y | Y | Y | Y | Y | 9 |
| 24 | Li & Newcombe (2013) | Y | Y | Y | Y | Y | Y | Y | Y | Y | 9 |

| # | Author (Year) | Q1 | Q2 | Q3 | Q4 | Q5 | Q6 | Q7 | Q8 | Q9 | Total |
|---|---|---|---|---|---|---|---|---|---|---|---|
| 25 | Li, Newcombe & Walton (2016) | Y | Y | Y | Y | Y | Y | Y | N | Y | 8 |
| 26 | Lykke, Pisinger & Glümer (2016) | Y | Y | Y | Y | Y | Y | Y | Y | Y | 9 |
| 27 | Moodie, Sinclair, Mackintosh, Power & Bauld (2016) | Y | Y | Y | Y | Y | Y | Y | N | Y | 8 |
| 28 | Morphett, Puljević, Borland, Carter, Hall & Gartner (2021) | Y | Y | Y | Y | Y | Y | Y | N | Y | 8 |
| 29 | Newcombe & Li (2013) | Y | Y | Y | Y | Y | Y | Y | Y | Y | 9 |
| 30 | Nogueira, Driezen, Fu, Hitchman, Tigova, Castellano, Kyriakos, Zatonski, Mons, Quah, Demjen, Trofor, Przewozniak, Katsaounou, Fong, Cardavas, Fernandez & EUREST-PLUS Consortium (2022) | Y | Y | Y | Y | Y | Y | Y | Y | Y | 9 |
| 31 | Palladino, Hone, Filippidis (2018) | Y | Y | Y | Y | N | Y | Y | Y | Y | 8 |
| 32 | Patel, Cuccia, Zhou, Czaplicki, Pitzer, Hair, Schillo & Vallone (2019) | Y | Y | Y | Y | U | Y | Y | N | Y | 7 |
| 33 | Pearson, Abrams, Niaura, Richardson, & Vallone (2013) | Y | Y | Y | Y | Y | Y | Y | Y | Y | 9 |
| 34 | Pepper, Squiers, Bann, Coglaiti & McCormack (2020) | Y | Y | N | Y | NA | Y | Y | N | NA | 5 |
| 35 | Robertson, Gendall, Hoek, Cameron, Marsh & McGee (2016) | Y | N | Y | Y | NA | Y | Y | N | NA | 5 |
| 36 | Schmidt, Kowitt, Myers & Goldstein (2018) | Y | Y | Y | Y | Y | Y | Y | N | Y | 8 |
| 37 | Siddiqi, Siddiqui, Boeckmann, Islam, Khan, Dobbie, Khan & Kannan (2022) | Y | Y | Y | Y | Y | Y | Y | N | Y | 8 |
| 38 | Smith, Nahhas, Borland, Cho, Chung-Hall, Fairman, Fong, McNeill, Popova, Thrasher & Cummings (2021) | Y | Y | Y | Y | Y | Y | Y | N | Y | 8 |
| 39 | Sonnenberg, Bostic & Halpern-Felsher (2020) | Y | N | N | Y | Y | Y | Y | N | N | 5 |
| 40 | Toxværd, Pisinger, Lykke & Lau (2023) | Y | Y | Y | Y | Y | Y | Y | Y | Y | 9 |
| 41 | Trainer, Gall, Smith & Terry (2017) | Y | Y | Y | Y | Y | Y | Y | Y | Y | 9 |
| 42 | Wamamili, Gartner & Lawler (2022) | Y | N | Y | Y | NA | Y | Y | N | NA | 5 |
| 43 | Wang, Wang, Lam, Viswanath & Chan (2015) | Y | Y | Y | Y | Y | Y | Y | Y | Y | 9 |
| 44 | White (2013) | Y | Y | Y | Y | Y | Y | Y | Y | Y | 9 |
| 45 | White (2015) | Y | Y | Y | Y | Y | Y | Y | Y | Y | 9 |
| 46 | Wu, Wang, Ho, Cheung, Kwong, Lai & Lam (2019) | Y | Y | Y | Y | Y | U | Y | Y | Y | 8 |
| 47 | Zatoński, Herbeć, Zatoński, Przewoźniak, Janik-Koncewicz, Mons, Fong, Demjén, Tountas, Trofor, Fernández, McNeil, Willemsen, Hummel, Quah, Kyriakos & Vardavas (2018) | Y | Y | Y | Y | Y | Y | N | Y | Y | 8 |

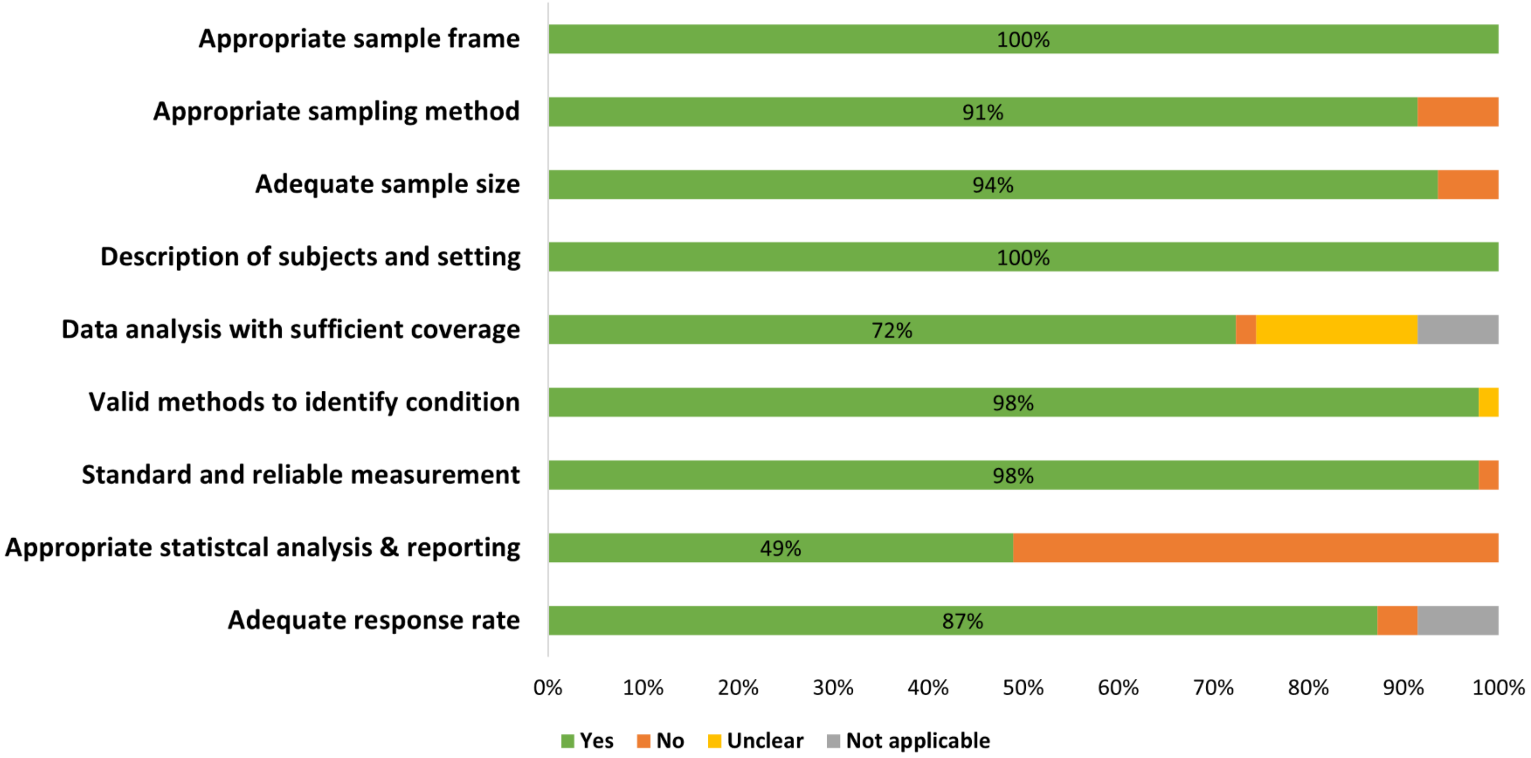

**Figure S7. Quality assessment results for the included studies**

### 8. Number of studies, estimates, and descriptive statistics for each policy/population group

**Table S8.** Number of studies, estimates, and descriptive statistics for each policy/population group

| Policy | | Group | Number of studies | Number of estimates | Minimum | Q1 | Median | Mean | Q3 | Maximum |
|---|---|---|---|---|---|---|---|---|---|---|
| Tobacco endgame goal | | General population | 4 | 5 | 56.0% | 74.0% | 74.0% | 71.5% | 74.6% | 79.0% |
| | | People who smoke | 5 | 7 | 19.0% | 34.0% | 42.1% | 39.0% | 46.5% | 50.6% |
| | | AYA | 2 | 3 | 51.0% | 55.0% | 59.0% | 60.3% | 65.0% | 71.0% |
| | | AYA who smoke | 1 | 2 | 9.0% | 9.2% | 9.5% | 9.5% | 9.8% | 10.0% |
| Limit nicotine | | General population | 9 | 10 | 46.5% | 67.3% | 75.9% | 70.9% | 80.3% | 86.1% |
| | | People who smoke | 12 | 17 | 30.1% | 53.8% | 62.9% | 61.9% | 72.0% | 80.6% |
| | | AYA | 2 | 2 | 47.4% | 55.4% | 63.3% | 63.3% | 71.3% | 79.3% |
| | | AYA who smoke | 1 | 1 | 71.9% | 71.9% | 71.9% | 71.9% | 71.9% | 71.9% |
| Set product standards | Ban all additives | General population | 1 | 1 | 69.2% | 69.2% | 69.2% | 69.2% | 69.2% | 69.2% |
| | | People who smoke | 6 | 9 | 29.7% | 34.5% | 42.5% | 44.1% | 49.1% | 61.7% |
| | | AYA | 0 | 0 | - | - | - | - | - | - |
| | | AYA who smoke | 2 | 2 | 36.7% | 40.7% | 44.8% | 44.8% | 48.8% | 52.8% |
| | Ban filters | General population | 1 | 1 | 51.3% | 51.3% | 51.3% | 51.3% | 51.3% | 51.3% |
| | | People who smoke | 1 | 1 | 35.8% | 35.8% | 35.8% | 35.8% | 35.8% | 35.8% |
| | | AYA | 0 | 0 | - | - | - | - | - | - |
| | | AYA who smoke | 0 | 0 | - | - | - | - | - | - |
| Move to reduced risk products | | General population | 0 | 0 | - | - | - | - | - | - |
| | | People who smoke | 1 | 1 | 66.2% | 66.2% | 66.2% | 66.2% | 66.2% | 66.2% |
| | | AYA | 0 | 0 | - | - | - | - | - | - |
| | | AYA who smoke | 0 | 0 | - | - | - | - | - | - |
| Require licence or prescription to purchase | | General population | 2 | 2 | 40.3% | 43.8% | 47.2% | 47.2% | 50.7% | 54.1% |
| | | People who smoke | 1 | 1 | 30.0% | 30.0% | 30.0% | 30.0% | 30.0% | 30.0% |
| | | AYA | 0 | 0 | - | - | - | - | - | - |
| | | AYA who smoke | 0 | 0 | - | - | - | - | - | - |
| Restrict sales by birth year (TFG) | | General population | 4 | 4 | 34.5% | 47.5% | 53.9% | 53.8% | 60.3% | 73.0% |

| Policy | | Group | Number of studies | Number of estimates | Minimum | Q1 | Median | Mean | Q3 | Maximum |
|---|---|---|---|---|---|---|---|---|---|---|
| | | People who smoke | 3 | 3 | 21.0% | 30.1% | 39.1% | 43.7% | 55.0% | 71.0% |
| | | AYA | 1 | 1 | 68.0% | 68.0% | 68.0% | 68.0% | 68.0% | 68.0% |
| | | AYA who smoke | 1 | 1 | 46.0% | 46.0% | 46.0% | 46.0% | 46.0% | 46.0% |
| Ban cigarette sales | | General population | 15 | 18 | 22.9% | 50.6% | 53.7% | 56.6% | 70.8% | 82.8% |
| | | People who smoke | 17 | 19 | 9.9% | 26.4% | 37.8% | 41.2% | 53.5% | 93.9% |
| | | AYA | 8 | 91 | 7.1% | 12.9% | 16.0% | 20.2% | 21.5% | 73.4% |
| | | AYA who smoke | 4 | 6 | 11.0% | 11.2% | 12.5% | 16.1% | 13.0% | 36.7% |
| Set quota (sinking lid) | | General population | 1 | 1 | 80.0% | 80.0% | 80.0% | 80.0% | 80.0% | 80.0% |
| | | People who smoke | 0 | 0 | - | - | - | - | - | - |
| | | AYA | 0 | 0 | - | - | - | - | - | - |
| | | AYA who smoke | 0 | 0 | - | - | - | - | - | - |
| Reduce company viability | | General population | 6 | 6 | 46.0% | 60.7% | 73.5% | 67.6% | 76.8% | 78.4% |
| | | People who smoke | 5 | 5 | 43.0% | 48.7% | 50.0% | 58.0% | 62.4% | 85.9% |
| | | AYA | 1 | 1 | 71.0% | 71.0% | 71.0% | 71.0% | 71.0% | 71.0% |
| | | AYA who smoke | 1 | 1 | 54.1% | 54.1% | 54.1% | 54.1% | 54.1% | 54.1% |
| Increase tax | | General population | 2 | 2 | 59.6% | 60.4% | 61.3% | 61.3% | 62.1% | 63.0% |
| | | People who smoke | 3 | 3 | 21.0% | 21.6% | 22.2% | 23.3% | 24.4% | 26.6% |
| | | AYA | 1 | 1 | 58.0% | 58.0% | 58.0% | 58.0% | 58.0% | 58.0% |
| | | AYA who smoke | 0 | 0 | - | - | - | - | - | - |
| Restrict retailer | Density | General population | 2 | 2 | 58.9% | 60.3% | 61.8% | 61.8% | 63.2% | 64.6% |
| | | People who smoke | 3 | 3 | 33.0% | 35.8% | 38.5% | 37.3% | 39.4% | 40.3% |
| | | AYA | 0 | 0 | - | - | - | - | - | - |
| | | AYA who smoke | 0 | 0 | - | - | - | - | - | - |
| | Type | General population | 1 | 1 | 22.0% | 22.0% | 22.0% | 22.0% | 22.0% | 22.0% |
| | | People who smoke | 1 | 1 | 15.0% | 15.0% | 15.0% | 15.0% | 15.0% | 15.0% |
| | | AYA | 0 | 0 | - | - | - | - | - | - |
| | | AYA who smoke | 0 | 0 | - | - | - | - | - | - |

### 9. Sensitivity analyses

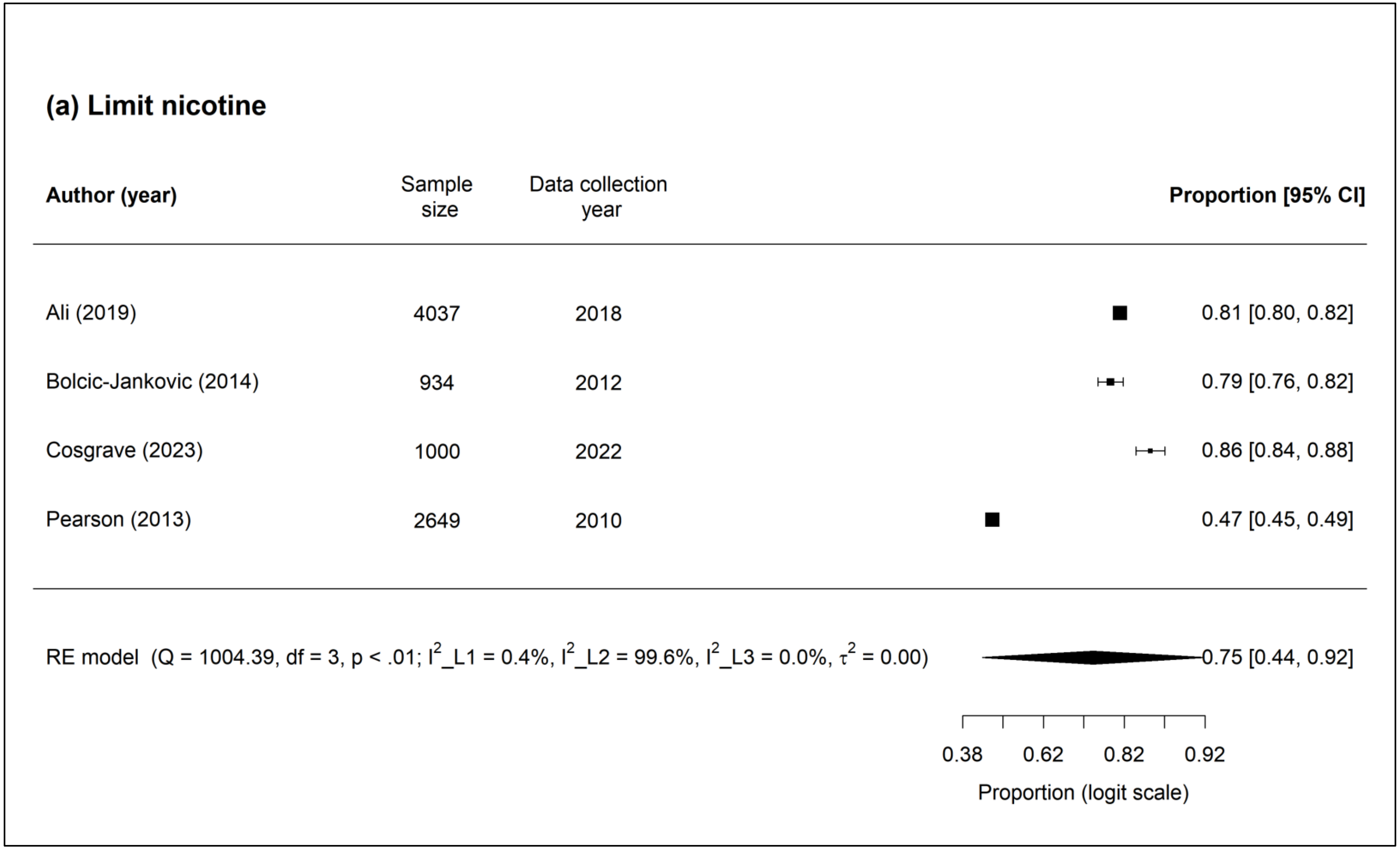

**Figure S9.1 Forest plot showing sensitivity analyses results for limiting nicotine in smoked tobacco products**

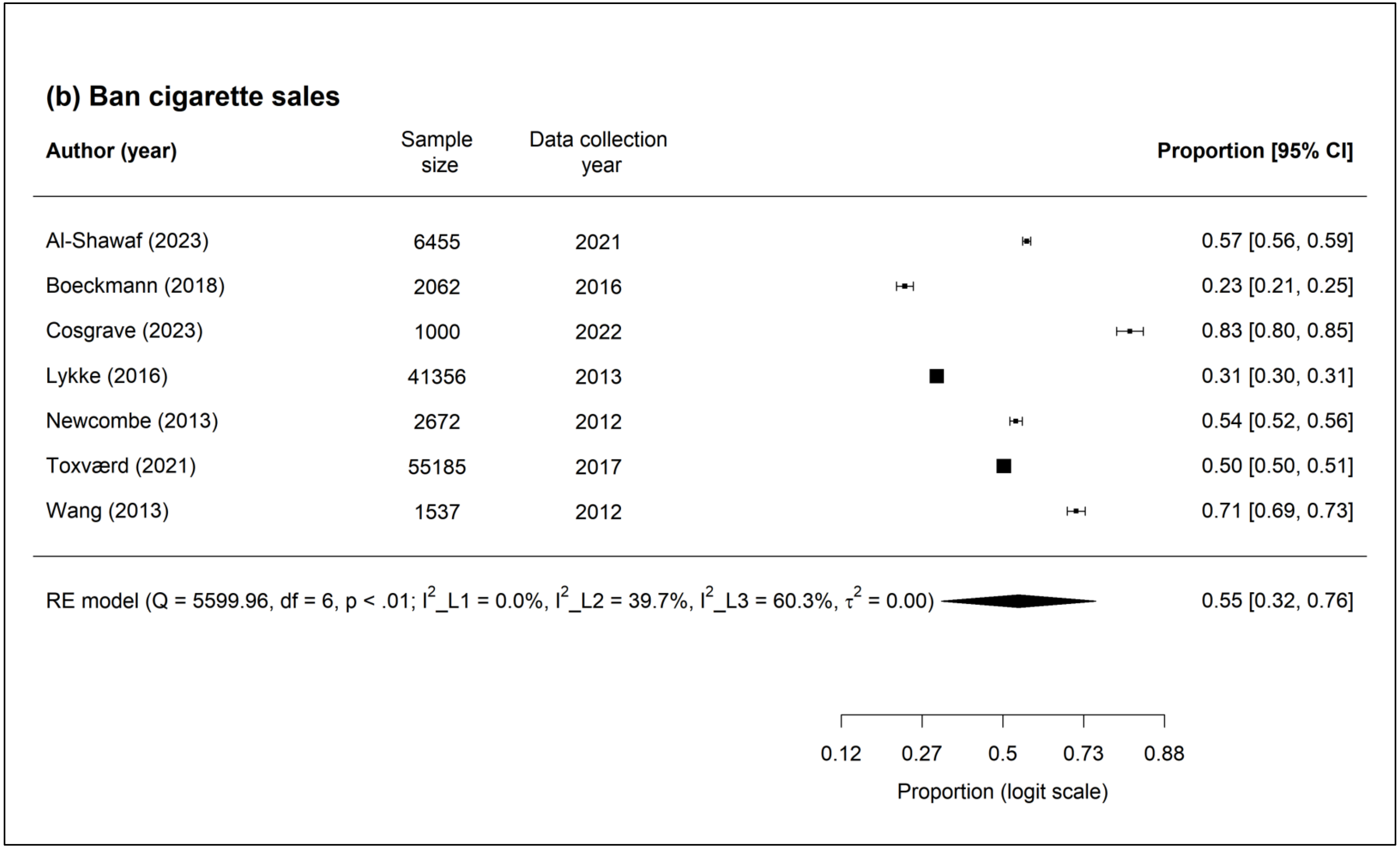

**Figure S9.2 Forest plot showing sensitivity analyses results banning cigarette sales**

### 10. Funnel plots

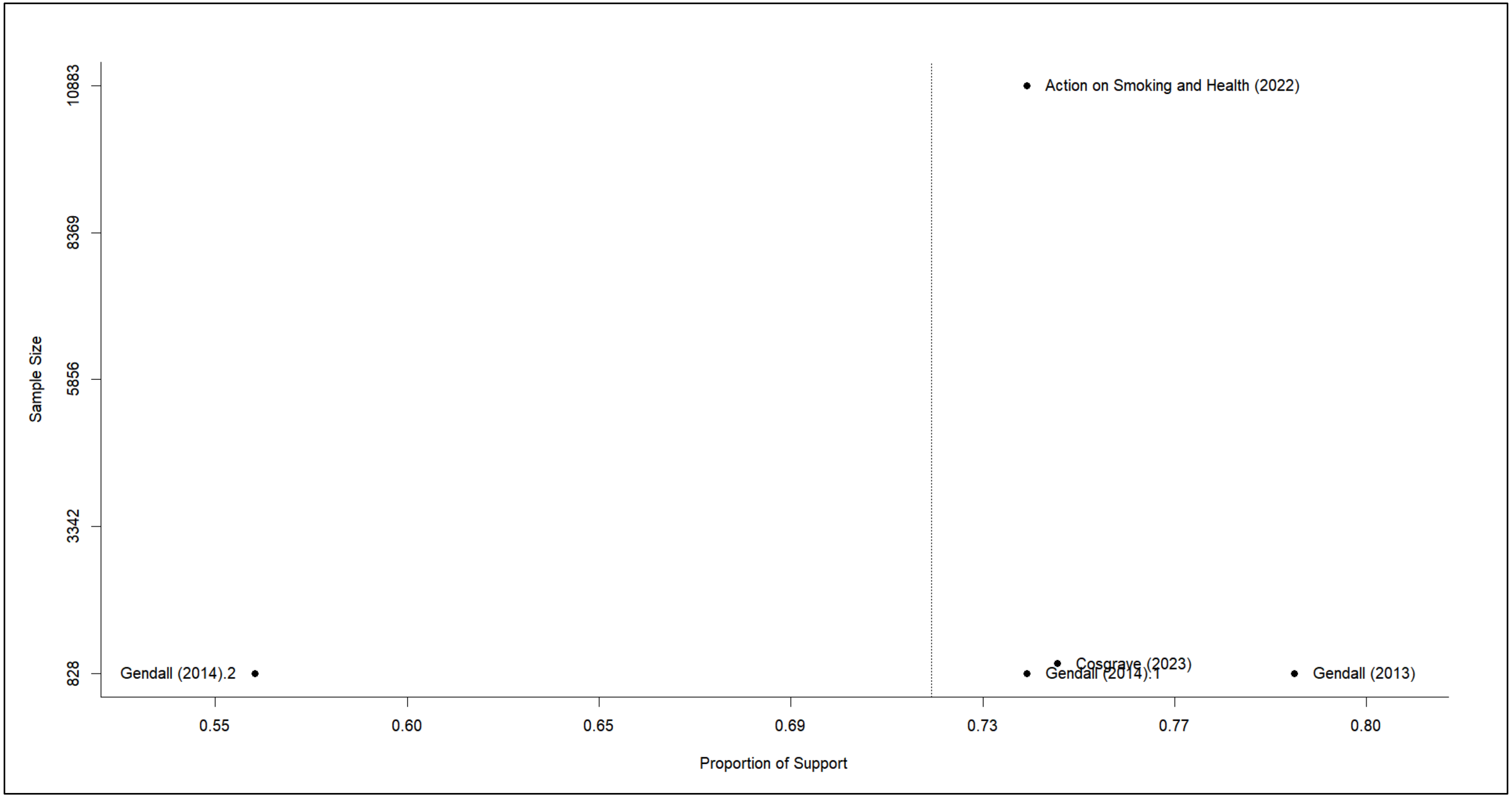

**Figure S10.1 Forest plot of sample size against proportion of support (tobacco endgame goal)**

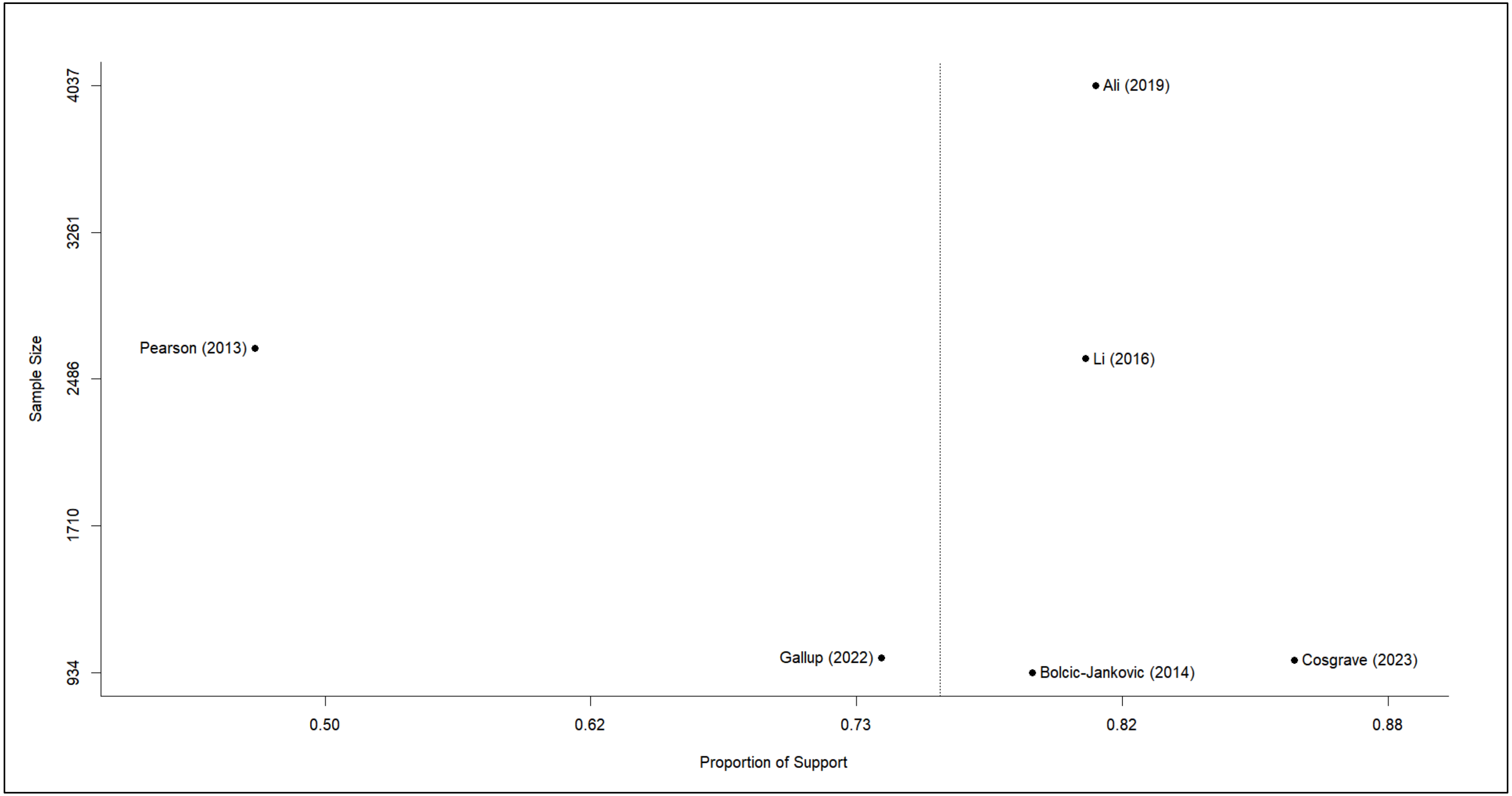

**Figure S10.2 Forest plot of sample size against proportion of support (limiting nicotine in smoked tobacco products)**

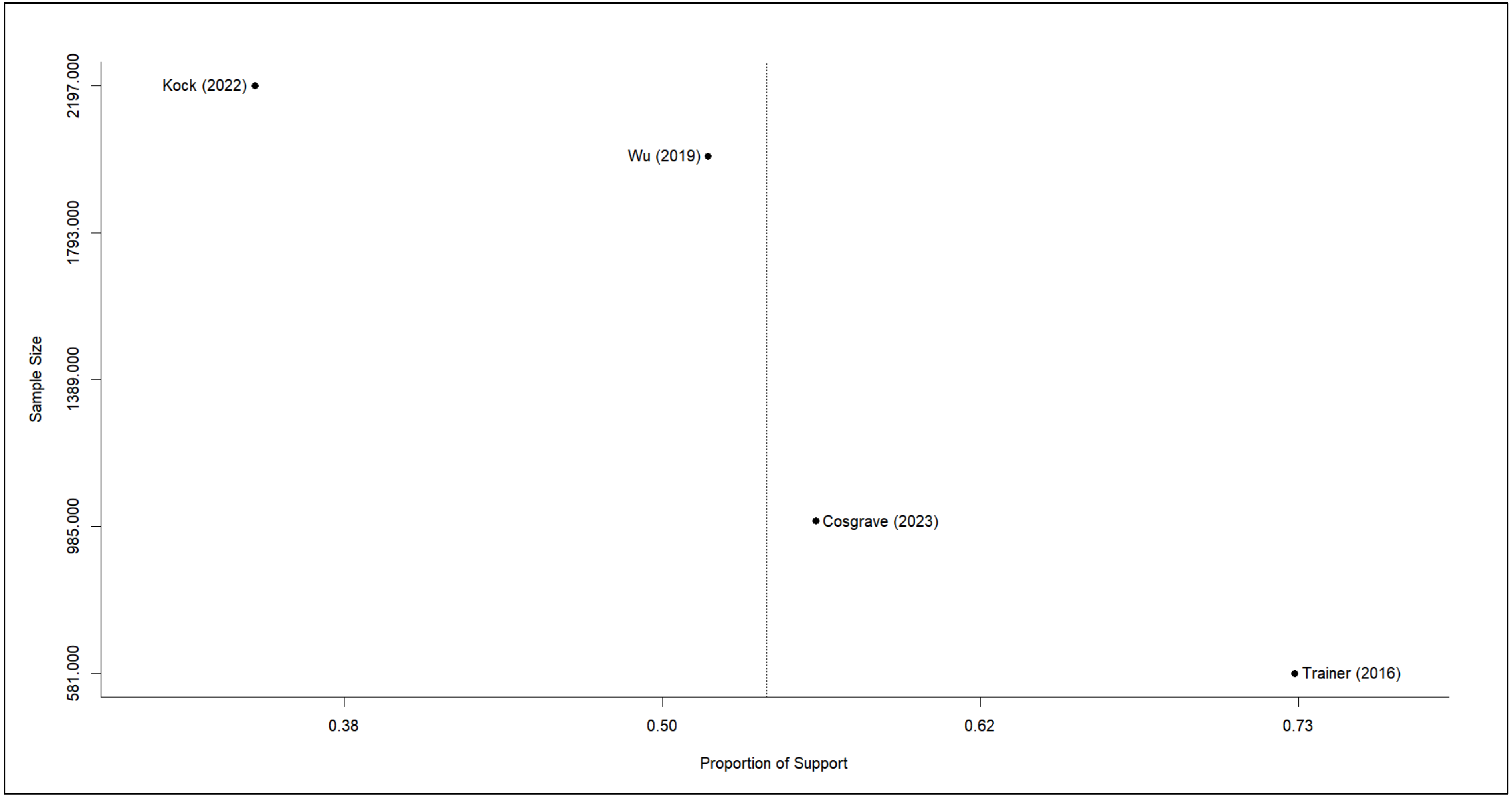

**Figure S10.3 Forest plot of sample size against proportion of support (restricting cigarettes supplies and sales by birth year, i.e., TFG)**

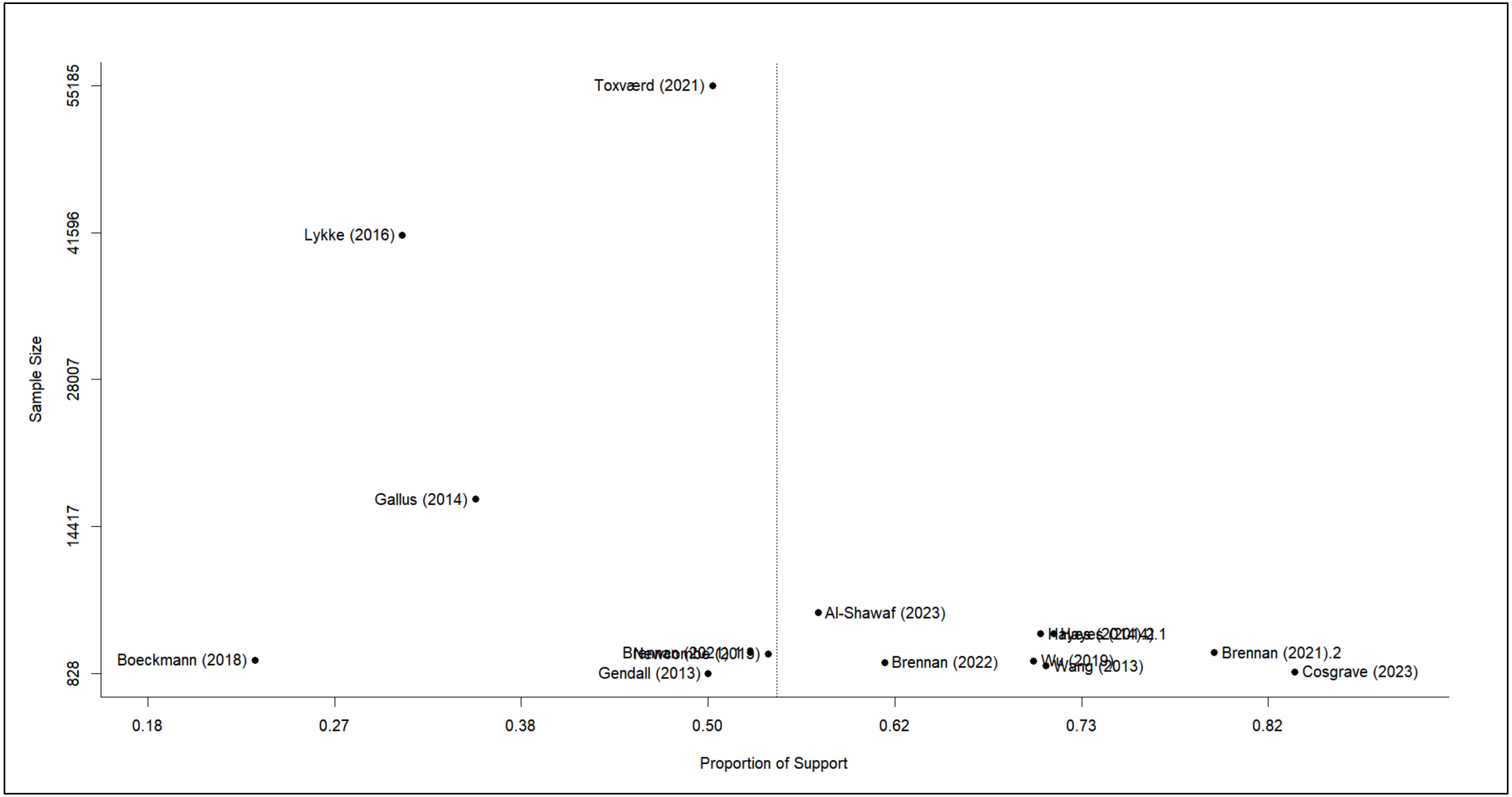

**Figure S10.4 Forest plot of sample size against proportion of support (banning cigarette sales)**

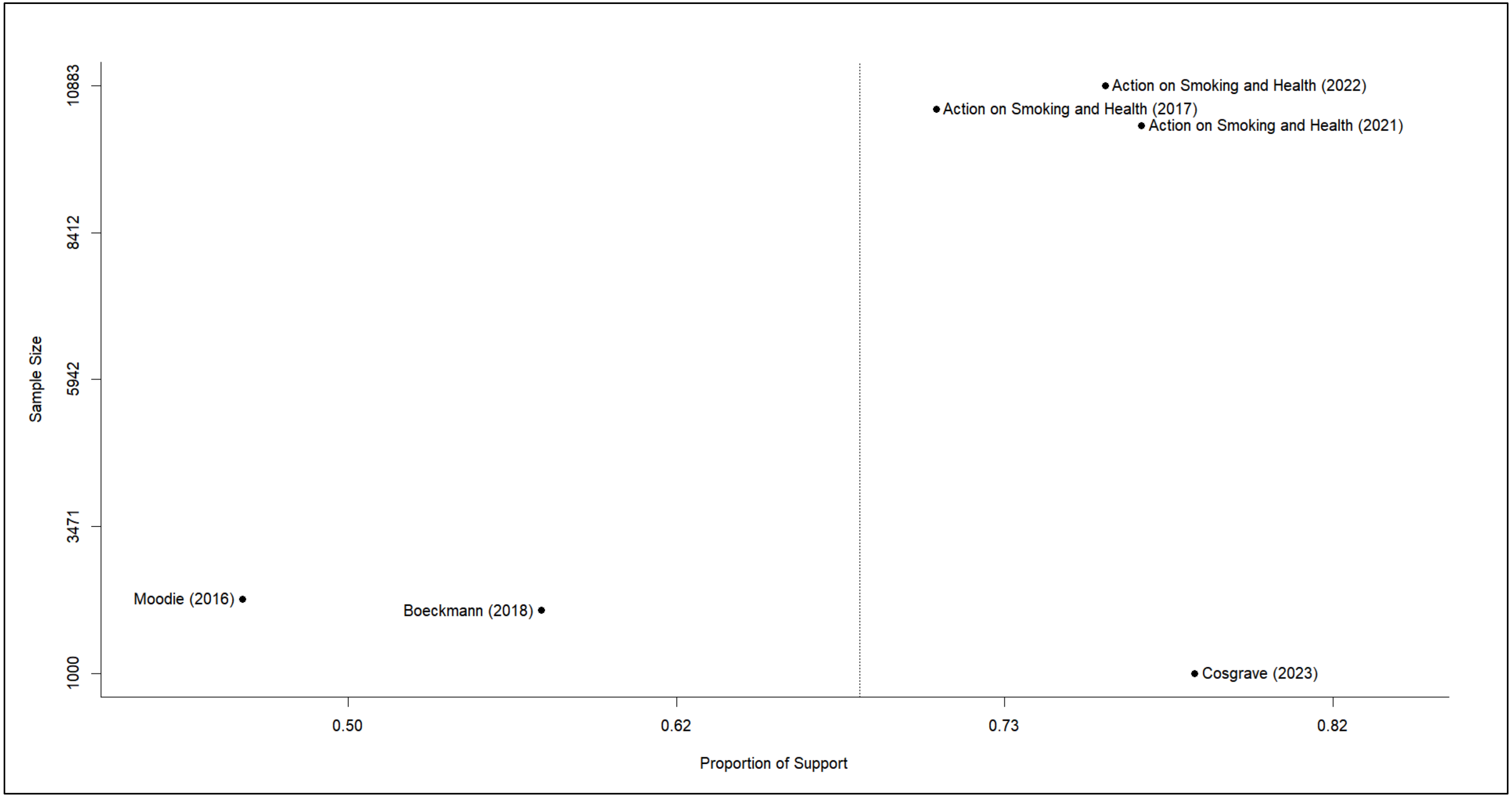

**Figure S10.5 Forest plot of sample size against proportion of support (reducing commercial viability of tobacco companies)**

### 11. Survey questions and response options

#### Table S11.1. Survey questions and response options used in each study/policy: Likert-type

| #* | Authors (Year) | Question asked** | Response options | % opinion | | | | | |
|---|---|---|---|---|---|---|---|---|---|
| | | | | Strongly agree | Agree | Neither agree nor disagree | Disagree | Strongly disagree | Don't know |
| **Tobacco endgame goal (n = 7)** | | | | | | | | | |
| 3 | Action on Smoking and Health (2022) | How strongly, if at all, do you support or oppose the following? The Government has set a target to end smoking by 2030. This will mean fewer than 5% of people smoking. | • Strongly support<br>• Tend to support<br>• Neither support nor oppose<br>• Tend to oppose<br>• Strongly oppose<br>• Don't know | 74% | | 19% (neither support nor oppose + don't know) | 7% | | 19% (neither support nor oppose + don't know) |
| 12 | Cosgrave, Blake, Murphy, Sheridan, Doyle & Kavanagh (2023) | The 'Tobacco-Free Ireland' goal aims to reduce the proportion of Irish adults who smoke to less than 5% by 2025. Would you say you… | • Strongly support this goal<br>• Support this goal<br>• Are neutral about this goal<br>• Oppose this goal<br>• Strongly oppose this goal<br>• Don't know | 74.6% | | Not reported | | | |
| 13 | Edwards, Johnson, Stanley, Waa, Ouimet & Fong (2021) | Do you support or oppose the Smokefree 2025 goal? | • Strongly support<br>• Support<br>• Oppose<br>• Strongly oppose<br>• Refused<br>• Don't know | 48.4% | | - | Not reported | Not reported | Excluded from the analyses |
| | | I'll now describe what the government's Smokefree 2025 goal states: the goal aims to reduce the number of people smoking and tobacco availability to minimal levels, thereby making New Zealand essentially a smokefree nation by 2025. 'Minimal numbers of people smoking' is often interpreted as less than 5% of people in all population groups will smoke. Do you support or oppose this Smokefree 2025 policy goal? | | 56.0% | | - | Not reported | Not reported | Excluded from the analyses |
| 17 | Gendall, Hoek & Edwards (2014) | Do you support or oppose the Smokefree 2025 goal, or do you have no preference either way? | • Support<br>• Oppose<br>• No preference<br>• Never heard of goal | 19% | | 24% | 48% | | 10% (never heard of goal) |
| | | To reduce smoking prevalence and tobacco availability to minimal levels, thereby making New Zealand essentially a smokefree nation by 2025. (This is often assumed to mean reducing | • Support<br>• Oppose<br>• No preference | 28% | | 24% | 48% | | - |

| #* | Authors (Year) | Question asked** | Response options | % opinion | | | | | |
|---|---|---|---|---|---|---|---|---|---|
| | | | | Strongly agree | Agree | Neither agree nor disagree | Disagree | Strongly disagree | Don't know |
| | | smoking to less than 5% of people in all population groups.) Do you support or oppose this Smokefree 2025 policy goal, or do you have no preference either way? | | | | | | | |
| 18 | Gendall, Hoek, Maubach & Edwards (2013) | I support the goal of reducing smoking from around 20% of the population to 5% or less by 2025 | • Strongly agree<br>• Agree<br>• Neither agree nor disagree<br>• Disagree<br>• Strongly disagree<br>• Don't know | 79% | | Not reported | 6% | | Excluded from the analyses |
| 20 | Jaine, Healey, Edwards & Hoek (2013) | For each of the statements listed below, please indicate whether you agree or disagree with them. I want to live in a country where no one smokes | • Agree<br>• Disagree<br>• Don't know | ('11) 51%<br>('12) 59% | | - | ('11) 22%<br>('12) 16% | | ('11) 28%<br>('12) 25% |
| 35 | Robertson, Gendall, Hoek, Cameron, Marsh & McGee (2016) | The government has a goal of making New Zealand smokefree by 2025. The goal is: To reduce smoking rates and tobacco sales to minimal levels, so that less than 5% of people smoke (in all population groups). Do you support or oppose this goal or do you have no preference either way? | • Support the 2025 goal<br>• Oppose the 2025 goal<br>• Have no preference either way | 40.0% | | Not reported | Not reported | | - |
| **Limiting nicotine in smoked tobacco products (n = 13)** | | | | | | | | | |
| 4 | Ali, Al-Shawaf, Wang, & King (2019) | Do you favor or oppose requiring cigarette makers to lower the nicotine levels in cigarettes so that they are less addictive? | • Strongly favor<br>• Somewhat favor<br>• Somewhat oppose<br>• Strongly oppose | 52.4% | 28.6% | - | 10.3% | 8.7% | - |
| 11 | Chung-Hall, Fong, Driezen & Craig (2018) | If you could get nicotine in products other than tobacco, would you support or oppose a law that reduced the amount of nicotine in cigarettes and tobacco, to make them less addictive? | • Strongly support<br>• Support<br>• Strongly oppose<br>• Oppose<br>• Refused<br>• Don't know | 70.2% | | - | Not reported | Not reported | Excluded from the analyses |
| 12 | Cosgrave, Blake, Murphy, Sheridan, Doyle & | Moving on, I am going to ask you about a number of potential measures that might help achieve the Tobacco-Free goal. To what extent do you agree or disagree with the following? The amount of nicotine in tobacco products should be reduced | • Strongly Agree<br>• Somewhat agree<br>• Neither agree nor disagree<br>• Somewhat disagree<br>• Strongly disagree | 86.1% | | 13.9% | | | |

| #* | Authors (Year) | Question asked** | Response options | % opinion | | | | | |
|---|---|---|---|---|---|---|---|---|---|
| | | | | Strongly agree | Agree | Neither agree nor disagree | Disagree | Strongly disagree | Don't know |
| | Kavanagh (2023) | through new laws to make tobacco products less addictive | • Don't know | | | | | | |
| 13 | Edwards, Johnson, Stanley, Waa, Ouimet & Fong (2021) | If you could get nicotine in products other than tobacco, would you support or oppose a law that reduces the amount of nicotine in cigarettes and tobacco, to make them less addictive? | • Strongly support<br>• Support<br>• Oppose<br>• Strongly oppose<br>• Refused<br>• Don't know | 72.9% | | - | Not reported | Not reported | Excluded from the analyses |
| 15 | Gallup (2022)[34] | Would you favor or oppose each of the following proposals? Requiring tobacco companies to lower nicotine levels in cigarettes to make them less addictive | • Favor<br>• Oppose<br>• No opinion/Don't know | 74% | | 0% | 26% | | 0% |
| 22 | Kulak, Kamper-DeMarco & Kozlowski (2020) | According to US law, the government can require the amount of nicotine in the tobacco in cigarettes be reduced. Would you support or oppose a law that reduced the amount of nicotine in all cigarettes, so every cigarette sold is very close to being free of nicotine, to make cigarettes less addictive? | • Strongly support<br>• Somewhat support<br>• Neutral<br>• Somewhat oppose<br>• Strongly oppose<br>• Don't know | (CS) 31.7%<br>(FS) 62.2%<br>(NS) 66.6% | (CS) 21.1%<br>(FS) 14.4%<br>(NS) 13.9% | (CS) 16.8%<br>(FS) 13.3%<br>(NS) 10.0% | (CS) 13.7%<br>(FS) 3.3%<br>(NS) 3.2% | (CS) 16.8%<br>(FS) 6.7%<br>(NS) 6.4% | Excluded from the analyses |
| 24 | Li & Newcombe (2013) | Whether or not you smoke, please tell me how much you agree or disagree with the following statements: The nicotine content of cigarettes should be reduced to very low levels so that they are less addictive | • Strongly agree<br>• Agree<br>• Neither agree nor disagree<br>• Disagree<br>• Strongly disagree<br>• Don't know<br>• Refused | (Quit attempters) 78.1%<br>(Non-attempters) 56.3% | | Not reported | Not reported | Not reported | Not reported |
| 25 | Li, Newcombe & Walton (2016) | The next few questions ask for your opinion about smoking. We are interested in these whether you smoke or not. The nicotine content of cigarettes should be reduced to very low levels so that they are less addictive… | • Strongly agree<br>• Agree<br>• Neither agree nor disagree<br>• Disagree<br>• Strongly disagree | 80.7% | | Not reported | Not reported | Not reported | Not reported |
| 32 | Patel, Cuccia, Zhou, Czaplicki, Pitzer, Hair, Schillo & Vallone (2019) | To what extent would you support a government policy that reduced the level of nicotine in cigarettes? | • Strongly support<br>• Somewhat support<br>• Somewhat oppose<br>• Strongly oppose | 72% | | - | 28% | | - |

| #* | Authors (Year) | Question asked** | Response options | % opinion | | | | | |
|---|---|---|---|---|---|---|---|---|---|
| | | | | Strongly agree | Agree | Neither agree nor disagree | Disagree | Strongly disagree | Don't know |
| 33 | Pearson, Abrams, Niaura, Richardson, & Vallone (2013) | Please tell us if you strongly agree, agree, neither agree nor disagree, disagree, or strongly disagree with the following statement (or do not know): The government should reduce the amount of nicotine in cigarettes to help smokers quit. | • Strongly agree<br>• Agree<br>• Neither agree nor disagree<br>• Disagree<br>• Strongly disagree | 46.7% | | 26.8% | 16.5% | | 10.0% |
| 34 | Pepper, Squiers, Bann, Coglaiti & McCormack (2020) | The Food and Drug Administration (FDA) recently announced that it wants to reduce the amount of nicotine in cigarettes to make them less addictive. Please indicate your agreement with the potential product standard. | • Scale from 1 = "strongly disagree" to 7 = "strongly agree" | 77.8% | Excluded from the analyses | Excluded from the analyses | Excluded from the analyses | 22.2% | Excluded from the analyses |
| 36 | Schmidt, Kowitt, Myers & Goldstein (2018) | Do you think the FDA should require companies to reduce the nicotine level in cigarettes? | • Yes<br>• No<br>• Don't know (not explicitly offered) | 71.0% | | - | 24.6% | | 4.4% |
| 38 | Smith, Nahhas, Borland, Cho, Chung-Hall, Fairman, Fong, McNeill, Popova, Thrasher & Cummings (2021) | If you could get nicotine in products other than tobacco, would you support or oppose a law that reduced the amount of nicotine in cigarettes and tobacco, to make them less addictive? | • Strongly support<br>• Support<br>• Oppose<br>• Strongly oppose<br>• Don't know<br>• Refused | (CA) 26.4%<br>(US) 17.9%<br>(UK) 18.9%<br>(AU) 18.6% | (CA) 41.6%<br>(US) 35.8%<br>(UK) 38.0%<br>(AU) 38.1% | - | (CA) 10.3%<br>(US) 16.6%<br>(UK) 12.9%<br>(AU) 17.0% | (CA) 6.5%<br>(US) 8.4%<br>(UK) 7.1%<br>(AU) 7.5% | (CA) 15.2%<br>(US) 21.3%<br>(UK) 23.1%<br>(AU) 18.8% |
| **Product standards to make smoked tobacco products unappealing or intolerable (banning all additives) (n = 6)** | | | | | | | | | |
| 11 | Chung-Hall, Fong, Driezen & Craig (2018) | Would you support or oppose a law that bans all additives, including flavourings, in cigarettes/tobacco? | • Strongly support<br>• Support<br>• Strongly oppose<br>• Oppose<br>• Refused<br>• Don't know | 42.5% | | - | Not reported | Not reported | Not reported |
| 12 | Cosgrave, Blake, Murphy, Sheridan, | Moving on, I am going to ask you about a number of potential measures that might help achieve the Tobacco-Free goal. To what extent do you agree or disagree with the following? Additive chemicals | • Strongly Agree<br>• Somewhat agree<br>• Neither agree nor disagree<br>• Somewhat disagree | 69.2% | | 30.8% | | | |

| #* | Authors (Year) | Question asked** | Response options | % opinion | | | | | |
|---|---|---|---|---|---|---|---|---|---|
| | | | | Strongly agree | Agree | Neither agree nor disagree | Disagree | Strongly disagree | Don't know |
| | Doyle & Kavanagh (2023) | that make cigarettes seem less harsh should be banned to make cigarettes more difficult to tolerate | • Strongly disagree<br>• Don't know | | | | | | |
| 13 | Edwards, Johnson, Stanley, Waa, Ouimet & Fong (2021) | Would you support or oppose a law that bans all additives, including flavourings, in cigarettes and tobacco? | • Strongly support<br>• Support<br>• Oppose<br>• Strongly oppose<br>• Refused<br>• Don't know | 53.2% | | - | Not reported | Not reported | Excluded from the analyses |
| 23 | Kyriakos, Fong, de Abreu Perez, Szklo, Driezen, Quah, Figueiredo & Filippidis (2022) | Would you support or oppose a law that banned all additives, including flavours, from cigarettes and tobacco? | • Strongly support<br>• Support<br>• Against<br>• Strongly against<br>• Don't know | 61.7% | | - | 29.3% | | 9.0% |
| 38 | Smith, Nahhas, Borland, Cho, Chung-Hall, Fairman, Fong, McNeill, Popova, Thrasher & Cummings (2021) | Would you support or oppose a law that bans all additives, including flavorings, in cigarettes/tobacco? | • Strongly support<br>• Support<br>• Oppose<br>• Strongly oppose<br>• Don't know | (CA) 15.0%<br>(US) 12.2%<br>(UK) 10.0%<br>(AU) 14.3% | (CA) 25.3%<br>(US) 22.3%<br>(UK) 19.7%<br>(AU) 20.0% | - | (CA) 27.1%<br>(US) 26.6%<br>(UK) 26.4%<br>(AU) 25.2% | (CA) 13.9%<br>(US) 18.5%<br>(UK) 16.6%<br>(AU) 16.6% | (CA) 18.7%<br>(US) 20.4%<br>(UK) 27.3%<br>(AU) 23.9% |
| 47 | Zatoński, Herbeć, Zatoński, Przewoźniak, Janik-Koncewicz, Mons, Fong, Demjén, Tountas, | Would you support or oppose a law that bans all additives, including flavorings, in cigarettes/tobacco? | • Strongly support<br>• Support<br>• Oppose<br>• Strongly oppose<br>• Refused<br>• Don't know | 46.3% | | - | NR | | |

| #* | Authors (Year) | Question asked** | Response options | % opinion | | | | | |
|---|---|---|---|---|---|---|---|---|---|
| | | | | Strongly agree | Agree | Neither agree nor disagree | Disagree | Strongly disagree | Don't know |
| | Trofor, Fernández, McNeil, Willemsen, Hummel, Quah, Kyriakos & Vardavas (2018) | | | | | | | | |
| **Product standards to make smoked tobacco products unappealing or intolerable (banning filters) (n = 1)** | | | | | | | | | |
| 12 | Cosgrave, Blake, Murphy, Sheridan, Doyle & Kavanagh (2023) | Moving on, I am going to ask you about a number of potential measures that might help achieve the Tobacco-Free goal. To what extent do you agree or disagree with the following? Filters on cigarettes and other combustible tobacco products should be banned to make the products more difficult to tolerate | • Strongly Agree<br>• Somewhat agree<br>• Neither agree nor disagree<br>• Somewhat disagree<br>• Strongly disagree<br>• Don't know | 51.3% | | 48.7% | | | |
| **Move consumers from smoked tobacco products to lower-risk product (n = 1)** | | | | | | | | | |
| 28 | Morphett, Puljević, Borland, Carter, Hall & Gartner (2021) | Please indicate how much you agree/disagree with the following statements. By "clean nicotine product", we mean nicotine gum, nicotine lozenges, e-cigarettes or other nicotine vaporisers. All smokers should be encouraged to switch to less harmful clean nicotine products as long-term substitutes for cigarettes | • Strongly agree<br>• Agree<br>• Disagree<br>• Strongly disagree<br>• Don't know | 20.4% | 45.8% | - | 10.7% | 5.5% | 17.6% |
| **Require consumers to obtain a license or prescription (n = 2)** | | | | | | | | | |
| 12 | Cosgrave, Blake, Murphy, Sheridan, Doyle & Kavanagh (2023) | Moving on, I am going to ask you about a number of potential measures that might help achieve the Tobacco-Free goal. To what extent do you agree or disagree with the following? People should be required to hold an official licence to buy tobacco products | • Strongly Agree<br>• Somewhat agree<br>• Neither agree nor disagree<br>• Somewhat disagree<br>• Strongly disagree<br>• Don't know | 40.3% | | 59.7% | | | |
| 46 | Wu, Wang, Ho, Cheung, Kwong, Lai & Lam (2019) | License to purchase combustible cigarettes *(?)* | • Yes<br>• No<br>• Don't know<br>• Refused | 54.1% | | - | Not reported | | Excluded from the analyses |
| **Restrict tobacco supplies and sales by birth year (tobacco-free generation) (n = 4)** | | | | | | | | | |

| #[*] | Authors (Year) | Question asked[**] | Response options | % opinion | | | | | |
|---|---|---|---|---|---|---|---|---|---|
| | | | | Strongly agree | Agree | Neither agree nor disagree | Disagree | Strongly disagree | Don't know |
| 12 | Cosgrave, Blake, Murphy, Sheridan, Doyle & Kavanagh (2023) | Moving on, I am going to ask you about a number of potential measures that might help achieve the Tobacco-Free goal. To what extent do you agree or disagree with the following? The government should prevent everyone who is currently under 18 from ever buying tobacco products for the rest of their lives | • Strongly Agree<br>• Somewhat agree<br>• Neither agree nor disagree<br>• Somewhat disagree<br>• Strongly disagree<br>• Don't know | 56.0% | | 44.0% | | | |
| 21 | Kock, Shahab, Moore, Shortt, Pearce & Brown (2022) | Please say to what extent you support or oppose each suggestion, or whether you have no opinion. Ban the sale of cigarettes and tobacco products to everyone born after a certain year from 2030 onward. | • Strongly support<br>• Tend to support<br>• No opinion either way<br>• Tend to oppose<br>• Strongly oppose<br>• Unsure or don't know | 34.5% | | 24.2% (No opinion either way + Unsure or don't know) | 41.3% | | 24.2% (No opinion either way + Unsure or don't know) |
| 41 | Trainer, Gall, Smith & Terry (2017) | Would you support a proposal that bans the sale of tobacco to Tasmanians born after the year 2000? | • Yes<br>• No<br>• Don't know | 73% | | - | 24% | | 3% |
| | | Would you support a proposal that stops the sale of tobacco to those Tasmanians born after the year 2000? | • Yes—support<br>• No—do not support<br>• Don't know/not sure | 68% | | - | 10% | | 22% |
| 46 | Wu, Wang, Ho, Cheung, Kwong, Lai & Lam (2019) | Ban people who are born after the year 2010 from smoking CCs *(?)* | • Yes<br>• No<br>• Don't know<br>• Refused | 51.8% | | - | Not reported | | Not reported |
| **Ban commercial sale of combustible tobacco (n = 20)** | | | | | | | | | |
| 5 | Al-Shawaf, Grooms, Mahoney, Lunsford & Kittner | To what extent would you support a policy to prohibit the sale of all tobacco products? | • Strongly support<br>• Somewhat support<br>• Somewhat oppose<br>• Strongly oppose | 30.0% | 27.3% | - | 21.8% | 20.9% | - |
| 6 | Avishai, Ribisl & Sheeran (2023) | 1) What percentage of people would need to die from smoking cigarettes in order to ban cigarettes (that is, to make it illegal to buy/sell cigarettes)<br>2) How many years would smoking cigarettes need to take off a person's life in order to ban cigarettes (that is, to make it illegal to buy/sell cigarettes) | • Specific percentage/number of years<br>• Never ban | 52.4% (If participant indicated both percentage and years) | | - | Not reported | | - |

| #* | Authors (Year) | Question asked** | Response options | % opinion | | | | | |
|---|---|---|---|---|---|---|---|---|---|
| | | | | Strongly agree | Agree | Neither agree nor disagree | Disagree | Strongly disagree | Don't know |
| 7 | Boeckmann, Kotz, Shahab, Brown & Kastaun (2018) | Do you support the following statements? The sale of cigarettes and tobacco in Germany should be banned completely within the next 10 years | • Strongly support<br>• Tend to support<br>• No opinion either way<br>• Tend to oppose<br>• Strongly oppose<br>• No answer | 22.9% | | 21.8% | 49.0% | | Not reported |
| 9 | Brennan, Durkin, Scollo, Swanson & Wakefield (2021) | Some people believe that there may come a time when it will no longer be legal to sell cigarettes in retail outlets in Australia. Do you think this would be… | • A good thing<br>• A bad thing<br>• Neither a good nor a bad thing<br>• Don't know/can't say | 52.8% | | 25.1% | 19.2% | | 2.8% |
| 10 | Brennan, Ilchenko, Scollo, Durkin & Wakefield (2022) | Some people believe that there may come a time when it will be no longer be legal to sell cigarettes in shops in Australia. Do you think it would be a good thing if there came a time when it was no longer legal to sell cigarettes in shops in Australia? | • A good thing<br>• A bad thing<br>• Neither a good nor a bad thing<br>• Don't know/refused | 61.6% | | 21.3% | 16.7% | | Not reported |
| 13 | Edwards, Johnson, Stanley, Waa, Ouimet & Fong (2021) | If the government provides assistance such as clinics to help smokers quit, would you support or oppose a law that… Totally bans cigarettes and other smoked tobacco within 10 years? | • Strongly support<br>• Support<br>• Oppose<br>• Strongly oppose<br>• Refused<br>• Don't know | 48.1% | | - | Not reported | Not reported | Excluded from the analyses |
| 14 | Edwards, Wilson, Peace, Weerasekera, Thomson & Gifford (2013) | If effective nicotine substitutes that are not smoked became available, the government should then set a date to ban cigarette sales in ten years time. | • Strongly agree<br>• Agree<br>• Neither agree nor disagree<br>• Disagree<br>• Strongly disagree<br>• Refused<br>• Don't know | 46.0% | | Not reported | 46.6% | | Not reported |
| 16 | Gallus, Lugo, Fernandez, Gilmore, Leon, Clancy & La Vecchia (2014) | The government or the national political decision makers could adopt<br>several strategies to control and limit tobacco use; how useful do you consider making smoking or cigarette sales illegal? | • Very useful<br>• Quite useful<br>• Rather useless<br>• Completely useless<br>• He/she does not know or He/she does not answer | 16.4% | 18.5% | - | 24.3% | 40.8% | Excluded from the analyses |

| #[*] | Authors (Year) | Question asked[**] | Response options | % opinion | | | | | |
|---|---|---|---|---|---|---|---|---|---|
| | | | | Strongly agree | Agree | Neither agree nor disagree | Disagree | Strongly disagree | Don't know |
| 18 | Gendall, Hoek, Maubach & Edwards (2013) | Cigarettes and tobacco should not be sold in New Zealand in ten years' time. | • Strongly agree<br>• Agree<br>• Neither agree nor disagree<br>• Disagree<br>• Strongly disagree<br>• Don't know | 50% | | Not reported | 25% | | Excluded from the analyses |
| 19 | Hayes, Wakefield & Scollo (2014) | Some people think that there will come a day when cigarettes will no longer be available for sale from retail outlets in Australia. Do you think this would be… | • A good thing<br>• A bad thing<br>• Neither a good or a bad thing<br>• Don't know/can't say | 71.6% | | 17.4% | 8.9% | | 2.1% |
| 20 | Jaine, Healey, Edwards & Hoek (2015) | For each of the statements listed below, please indicate whether you agree or disagree with them. Cigarettes and tobacco should not be sold in New Zealand in 10 years' time | • Agree<br>• Disagree<br>• Don't know | ('10) 48%<br>('11) 52%<br>('12) 57% | | - | ('10) 23%<br>('11) 20%<br>('12) 16% | | ('10) 30%<br>('11) 28%<br>('12) 27% |
| 29 | Newcombe & Li (2013) | Whether or not you smoke, please tell me how much you agree or disagree with the following statements. Cigarettes and tobacco should not be sold in New Zealand in 10 years' time. | • Strongly agree<br>• Agree<br>• Neither agree nor disagree<br>• Disagree<br>• Strongly disagree<br>• Don't know<br>• Refused | 18% | 36% | 13% | Not reported | Not reported | Not reported |
| 30 | Nogueira, Driezen, Fu, Hitchman, Tigova, Castellano, Kyriakos, Zatonski, Mons, Quah, Demjen, Trofor, Przewozniak, Katsaounou, Fong, Cardavas, Fernandez & EUREST-PLUS | Would you support or oppose a total ban on cigarettes and other smoked tobacco within 10 years, if the government provided assistance such as cessation clinics to help smokers quit? | • Strongly support<br>• Support<br>• Oppose<br>• Strongly oppose<br>• Refused<br>• Don't know | 37.8% | | - | Not reported | Not reported | Not reported |

| #[*] | Authors (Year) | Question asked[**] | Response options | % opinion | | | | | |
|---|---|---|---|---|---|---|---|---|---|
| | | | | Strongly agree | Agree | Neither agree nor disagree | Disagree | Strongly disagree | Don't know |
| | Consortium (2022) | | | | | | | | |
| 31 | Palladino, Hone, Filippidis (2018) | The sale of drugs such as cannabis, cocaine, ecstasy, and heroin is officially banned in all EU Member States. The sale of legal substances such as alcohol and tobacco is not prohibited but is regulated in all EU countries, which means for example that there is a minimum age limit for buying, limits in the concentration of active components, or licensed sales through specialised shops and pharmacies. Do you think the following substances should continue to be banned or should be banned, or should they be regulated? | • Should continue to be banned or should be banned<br>• Should be regulated<br>• Should be available without restrictions | ('08) 17.9%<br>('11) 16.5%<br>('14) 16.0% | | - | Not reported | | - |
| 37 | Siddiqi, Siddiqui, Boeckmann, Islam, Khan, Dobbie, Khan & Kannan (2022) | Do you support the following statements? The sale of cigarettes and tobacco in Pakistan should be banned completely within the next 10 years | • Strongly support<br>• Tend to support<br>• No opinion either way<br>• Tend to oppose<br>• Strongly oppose | 93.9% | | 3.4% | 2.7% | | - |
| 39 | Sonnenberg, Bostic & Halpern-Felsher (2020) | How much do you agree with the following statement for each of the following products. There should be a gradual ban on the sale of cigarettes | • Strongly disagree<br>• Disagree<br>• Agree<br>• Strongly agree | 37.6% | 35.8% | - | 16.0% | 7.1% | 3.6% (missing data) |
| | | How much do you agree with the following statement for each of the following products. There should be an immediate ban on the sale of cigarettes | | 29.3% | 27.7% | - | 26.8% | 11.1% | 4.9% (missing data) |
| 42 | Wamamili, Gartner & Lawler (2022) | How much do you agree or disagree with the following statements?: Cigarettes should not be sold (in Australia or NZ) in 10 years' time (i.e., ending tobacco sales). | • Strongly agree<br>• Agree<br>• Neutral<br>• Strongly disagree<br>• Disagree | (NZ) 53.3%<br>(AU) 51.6% | | (NZ) 28.1%<br>(AU) 24.3% | (NZ) 18.6%<br>(AU) 24.1% | | - |
| 44 | White (2015) | For each of the statements below, please indicate whether you agree or disagree with them. Cigarettes and tobacco should not be sold in NZ | • Agree<br>• Disagree<br>• Don't know | 56% | | - | 22% | | 22% |
| 45 | White (2013) | For each of the statements below, please indicate whether you agree or disagree with them. Cigarettes and tobacco should not be sold in NZ | • Agree<br>• Disagree<br>• Don't know | 57% | | - | 21% | | 22% |

| #[*] | Authors (Year) | Question asked[**] | Response options | % opinion | | | | | |
|---|---|---|---|---|---|---|---|---|---|
| | | | | Strongly agree | Agree | Neither agree nor disagree | Disagree | Strongly disagree | Don't know |
| 46 | Wu, Wang, Ho, Cheung, Kwong, Lai & Lam (2019) | Ban combustible cigarette sales in 10 years if there is a product providing nicotine not made from tobacco *(?)* | • Yes<br>• No<br>• Don't know<br>• Refused | 51.8% | | - | Not reported | | Not reported |
| **Set a regularly reducing quota on the volume manufactured or imported (sinking lid) (n = 1)** | | | | | | | | | |
| 46 | Wu, Wang, Ho, Cheung, Kwong, Lai & Lam (2019) | Set a quota for combustible cigarette retail and reduce it on a yearly basis | • Yes<br>• No<br>• Don't know<br>• Refused | 80.0% | | - | Not reported | | Not reported |
| **Reduce commercial viability of tobacco companies (n = 8)** | | | | | | | | | |
| 1 | Action on Smoking and Health (2017) | Below is a suggestion that has been made to reduce smoking. How strongly, if at all, would you support or oppose the following measure...? Requiring tobacco manufacturers to pay a levy to Government for measures to help smokers quit and prevent young people from taking up smoking | • Strongly support<br>• Tend to support<br>• Neither support nor oppose<br>• Tend to oppose<br>• Strongly oppose<br>• Don't know | 71.0% | | Not reported | 9.0% | | Not reported |
| 2 | Action on Smoking and Health (2021) | Below is a suggestion that has been made to reduce smoking. How strongly, if at all, would you support or oppose the following measure...? Requiring tobacco manufacturers to pay a levy to Government for measures to help smokers quit and prevent young people from taking up smoking | • Strongly support<br>• Tend to support<br>• Neither support nor oppose<br>• Tend to oppose<br>• Strongly oppose<br>• Don't know | 77.0% | | Not reported | 6.0% | | Not reported |
| 3 | Action on Smoking and Health (2022) | Below is a suggestion that has been made to reduce smoking. How strongly, if at all, would you support or oppose the following measure...? Requiring tobacco manufacturers to pay a levy to Government for measures to help smokers quit and prevent young people from taking up smoking | • Strongly support<br>• Tend to support<br>• Neither support nor oppose<br>• Tend to oppose<br>• Strongly oppose<br>• Don't know | 76.0% | | 18.0% (neither support nor oppose + don't know) | 6.0% | | 18.0% (neither support nor oppose + don't know) |
| 7 | Boeckmann, Kotz, Shahab, Brown & Kastaun (2018) | Do you support the following statements? Tobacco industry sales should be taxed in order to use the money to address problems caused by tobacco (e.g., health issues, environmental problems, etc.) | • Strongly support<br>• Tend to support<br>• No opinion either way<br>• Tend to oppose<br>• Strongly oppose<br>• No answer | 57.3% | | Not reported | Not reported | | Not reported |
| 12 | Cosgrave, Blake, Murphy, | Now I would like to discuss measures which target the tobacco industry. To what extent do you agree or disagree with the following? Tobacco | • Strongly Agree<br>• Somewhat agree | 78.4% | | 21.6% | | | |

| #* | Authors (Year) | Question asked** | Response options | % opinion | | | | | |
|---|---|---|---|---|---|---|---|---|---|
| | | | | Strongly agree | Agree | Neither agree nor disagree | Disagree | Strongly disagree | Don't know |
| | Sheridan, Doyle & Kavanagh (2023) | companies should be required to pay the state for the health costs due to the harm caused by tobacco products | • Neither agree nor disagree<br>• Somewhat disagree<br>• Strongly disagree<br>• Don't know | | | | | | |
| 27 | Moodie, Sinclair, Mackintosh, Power & Bauld (2016) | How much do you agree or disagree with the following statements? Tobacco companies should pay for the health related costs associated with tobacco use | • Strongly Agree<br>• Somewhat agree<br>• Neither agree nor disagree<br>• Somewhat disagree<br>• Strongly disagree<br>• Don't know | 46% | | Not reported | 27% | | Not reported |
| 30 | Nogueira, Driezen, Fu, Hitchman, Tigova, Castellano, Kyriakos, Zatonski, Mons, Quah, Demjen, Trofor, Przewozniak, Katsaounou, Fong, Cardavas, Fernandez & EUREST-PLUS Consortium (2022) | How much do you agree with the following statement: tobacco companies should take responsibility for the harm caused by smoking | • Strongly Agree<br>• Agree<br>• Neither agree nor disagree<br>• Disagree<br>• Strongly disagree<br>• Refused<br>• Don't know | 48.7% | | Not reported | Not reported | Not reported | Not reported |
| 37 | Siddiqi, Siddiqui, Boeckmann, Islam, Khan, Dobbie, Khan & Kannan (2022) | Do you support the following statements? Tobacco industry sales should be taxed in order to use the money to address problems caused by tobacco (eg, health issues) | • Strongly support<br>• Tend to support<br>• No opinion either way<br>• Tend to oppose<br>• Strongly oppose | 85.9% | | 6.1% | 8.0% | | - |
| **Increase taxes to make tobacco products unaffordable (n = 3)** | | | | | | | | | |

| #* | Authors (Year) | Question asked** | Response options | % opinion | | | | | |
|---|---|---|---|---|---|---|---|---|---|
| | | | | Strongly agree | Agree | Neither agree nor disagree | Disagree | Strongly disagree | Don't know |
| 3 | Action on Smoking and Health (2022) | Below is a suggestion that has been made to reduce smoking. How strongly, if at all, would you support or oppose the following measure...? Tax should be used to increase the price of tobacco products 5% above the rate of inflation each year | • Strongly support<br>• Tend to support<br>• Neither support nor oppose<br>• Tend to oppose<br>• Strongly oppose<br>• Don't know | 63% | | 21% (neither support nor oppose + don't know) | 16% | | 21% (neither support nor oppose + don't know) |
| 12 | Cosgrave, Blake, Murphy, Sheridan, Doyle & Kavanagh (2023) | Moving on, I am going to ask you about a number of potential measures that might help achieve the Tobacco-Free goal. To what extent do you agree or disagree with the following? The Government should increase the tax on tobacco products by 20% a year until less than 5% of people smoke | • Strongly Agree<br>• Somewhat agree<br>• Neither agree nor disagree<br>• Somewhat disagree<br>• Strongly disagree<br>• Don't know | 59.6% | | 40.4% | | | |
| 13 | Edwards, Johnson, Stanley, Waa, Ouimet & Fong (2021) | Would you support or oppose each of the following initiatives? The government should INCREASE the tax on tobacco by 20% a year until less than five percent of the population smoke. | • Strongly support<br>• Support<br>• Oppose<br>• Strongly oppose<br>• Refused<br>• Don't know | 26.7% | | - | Not reported | Not reported | Excluded from the analyses |
| **Restrict retailers to substantially limit availability (n = 4)** | | | | | | | | | |
| 10 | Brennan, Ilchenko, Scollo, Durkin & Wakefield (2022) | Shop owners should seriously be thinking about transitioning out of selling cigarettes *(?)* | • Strongly support<br>• Somewhat support<br>• Somewhat oppose<br>• Strongly oppose | 64.6% | | Not reported | Not reported | | Not reported |
| 12 | Cosgrave, Blake, Murphy, Sheridan, Doyle & Kavanagh (2023) | Moving on, I am going to ask you about a number of potential measures that might help achieve the Tobacco-Free goal. The number of places that can sell tobacco products should be reduced by 95% | • Strongly Agree<br>• Somewhat agree<br>• Neither agree nor disagree<br>• Somewhat disagree<br>• Strongly disagree<br>• Don't know | 58.9% | | 41.1% | | | |
| 13 | Edwards, Johnson, Stanley, Waa, Ouimet & Fong (2021) | I am now going to read you some statements about tobacco companies and tobacco sales. Please tell me what you think of each statement. The number of places that can sell tobacco products should be greatly reduced, that is by 95%, and sales allowed | • Strongly agree<br>• Agree<br>• Neither agree nor disagree<br>• Disagree<br>• Strongly disagree | 42.8% | | Not reported | Not reported | Not reported | Excluded from the analyses |

| #* | Authors (Year) | Question asked** | Response options | % opinion | | | | | |
|---|---|---|---|---|---|---|---|---|---|
| | | | | Strongly agree | Agree | Neither agree nor disagree | Disagree | Strongly disagree | Don't know |
| | | only in a limited number and type of stores. | • Refused<br>• Don't know | | | | | | |
| 18 | Gendall, Hoek, Maubach & Edwards (2013) | Only people qualified to give quitting advice should be allowed to sell tobacco products | • Strongly agree<br>• Agree<br>• Neither agree nor disagree<br>• Strongly disagree<br>• Disagree<br>• Don't know | 22% | | Not reported | 46% | | Excluded from the analyses |

*Numbers correspond to numbers in Table S3
**We attempted to contact the authors of studies that did not provide the full survey question and responses in the published article. In cases where we did not receive a response after three attempts, we added a notation of '(?)' in the 'question is asked' column

### Table S11.2. Survey questions and response options used in each study/policy: forced-type

| #* | Authors (Year) | Question asked | Response options | % opinion |
|---|---|---|---|---|
| **Limiting nicotine in smoked tobacco products (n = 2)** | | | | |
| 8 | Bolcic-Jankovic & Biener (2015) | Nicotine is the substance in cigarettes that makes people get addicted to smoking. The FDA has the authority to reduce the amount of nicotine in cigarettes to a very low level. What do you think the FDA should do? | • Require that cigarette manufacturers reduce the level of nicotine to the lowest level possible | 46.6% |
| | | | • Set a limit for the amount of nicotine permitted in cigarettes, gradually lower that level over the next 10 years | 32.5% |
| | | | • Do nothing about nicotine in cigarettes | 21.0% |
| 22 | Kulak, Kamper-DeMarco & Kozlowski (2020) | Which one of the following best describes a policy that you would support? | • I support that the levels of nicotine in cigarette vary from zero or very low levels of nicotine in some brands to high nicotine levels in others, so that smokers can chose the nicotine levels they like. No nicotine levels would be banned | 41.0% |
| | | | • I support that the levels of nicotine in cigarette vary from zero or very low levels of nicotine in some brands to moderate nicotine levels in others. Smokers would have some choice over the nicotine levels they like, but high nicotine cigarettes would not be sold | 28.6% |
| | | | • I support that only very low levels of nicotine be allowed in all cigarettes sold. Nicotine would not be banned, but these low levels would not cause any addiction or nicotine effects to occur and would help increase the chances of quitting smoking | 20.5% |
| | | | • I support the banning of nicotine in all cigarettes. Only nicotine free cigarettes could be sold. This would stop all nicotine effects, stop the chances of addiction, and help increase the chances of quitting smoking | 9.9% |
| **Ban commercial sale of combustible tobacco (n = 1)** | | | | |
| 12 | Cosgrave, Blake, Murphy, Sheridan, Doyle & Kavanagh (2023) | Which of the following, if any, comes closest to your own view on the sale of tobacco products? | • Tobacco product sales should be phased out<br>• Tobacco product sales should be phased out but only if the government provides assistance to help smokers to quit<br>• Tobacco product sales should be phased out but only if existing smokers can continue to buy tobacco products using a licence<br>• Tobacco product sales should be phased out but only if the government provides assistance to help smokers to quit AND existing smokers can continue to buy tobacco products using a licence | 82.8% |
| | | | • Tobacco product sales should not be phased out<br>• None of these/other option<br>• Don’t know | 17.2% |

*Numbers correspond to numbers in Table S3

### Table S10.3. Survey questions and response options used in each study/policy: timeframe

| #* | Authors (Year) | Question asked | Response options | % opinion | | | | | | | | | |
|---|---|---|---|---|---|---|---|---|---|---|---|---|---|
| | | | | Immediately | ≤1 year | ≤ 3 years | ≤5 years | ≤10 years | ≤15 years | ≤20 years | > 20 years | Never | Don't know |
| **Ban commercial sale of combustible tobacco (n = 7)** | | | | | | | | | | | | | |
| 9 | Brennan, Durkin, Scollo, Swanson & Wakefield (2021) | What timeframe do you think is fair in relation to the proposed phasing out of the sale of cigarettes from retail outlets? Would you say… | • Within the next 5 years<br>• Within the next 10 years<br>• Within the next 20 years<br>• Yes, but not within the next 20 years<br>• No, not ever<br>• Don't know/can't say | - | - | - | 38.2% | 26.0% | - | 11.9% | 3.4% | 16.6% | 3.9% |
| 10 | Brennan, Ilchenko, Scollo, Durkin & Wakefield (2022) | What timeframe would you think is appropriate in relation to phasing out the sale of cigarettes from shops? | • Within the next 5 years<br>• Within the next 10 years<br>• Within the next 15 years<br>• Within the next 20 years<br>• Not for at least 20 years<br>• Don't know/refused | - | - | - | 38.5% | Not reported | | | | - | Not reported |
| 19 | Hayes, Wakefield & Scollo (2014) | Some people believe that there may come a time when it will no longer be legal to sell cigarettes in retail outlets in Australia. What timeframe do you think is fair in relation to the proposed phasing out of the sale of cigarettes from retail outlets? Would you say… | • Within the next 5 years<br>• Within the next 10 years<br>• Within the next 20 years<br>• Yes, but not within the next 20 years<br>• No, not ever<br>• Don't know/can't say | - | - | - | 31.4% | 21.4% | - | 12.2% | 5.9% | 25.6% | 3.6% |
| 26 | Lykke, Pisinger & Glümer (2016)[18] | Do you think it's a good idea to set a date for when smoking in Denmark should be banned (with the possibility of tobacco | • No<br>• Yes, within 10 years<br>• Yes, within 20 years<br>• Yes, within 30 years or more | - | - | - | - | 23.3% | - | 3.9% | 3.4% | 69.4% | - |

| | | | | | | | | | | | | | |
|---|---|---|---|---|---|---|---|---|---|---|---|---|---|
| | | on prescription for already addicted smokers? | | | | | | | | | | | |
| 40 | Toxværd, Pisinger, Lykke & Lau (2023)[45] | Do you think it's a good idea to set a date for when smoking in Denmark should be banned (with the possibility of tobacco on prescription for already addicted smokers? | • No<br>• Yes, within 10 years<br>• Yes, within 20 years<br>• Yes, within 30 years or more | - | - | - | 36.1% | 8.2% | - | 3.6% | 2.4% | 49.7% | - |
| 43 | Wang, Wang, Lam, Viswanath & Chan (2015)[49] | Do you support a total ban on tobacco sales in Hong Kong and when should the ban be implemented? | • Immediately<br>• Within 1 year<br>• Within 3 years<br>• Within 5 years<br>• Within 10 years<br>• Within 20 years<br>• After 20 years<br>• Not sure when<br>• Did not support a total ban | 64.8% | | | | | 6.4% | | | 28.8% | Excluded |
| 46 | Wu, Wang, Ho, Cheung, Kwong, Lai & Lam (2019) | Do you support completely banning the sale of CCs? | • Yes, Immediately<br>• Yes, in 1 year<br>• Yes, in 3 years<br>• Yes, in 5 years<br>• Yes, in 10 years<br>• Yes, after 10 years<br>• Yes, but deadline not yet determined<br>• No, people have a right to smoke<br>• No, it will have a negative effect on the economy<br>• No, ban is useless<br>• No, it will increase CC smuggling<br>• No, I am a smoker<br>• No, other reasons<br>• Don't know<br>• Refused | 63.8% | | | | | Not reported | Not reported | Not reported | Not reported | Not reported |

*Numbers correspond to numbers in Table S3

16. Action on Smoking and Health. Public support in England for Government action on tobacco: Results extracted from the ASH Smokefree GB Survey 2022

London: ASH, 2022.

17. Moodie C, Sinclair L, Mackintosh AM, et al. How Tobacco Companies are Perceived Within the United Kingdom: An Online Panel. *Nicotine & Tobacco Research* 2016;18(8):1766-72. doi: 10.1093/ntr/ntw024

18. Lykke M, Pisinger C, Glümer C. Ready for a goodbye to tobacco? — Assessment of support for endgame strategies on smoking among adults in a Danish regional health survey. *Preventive Medicine* 2016;83:5-10. doi: https://doi.org/10.1016/j.ypmed.2015.11.016

19. White J. Young peoples' opinions on tobacco control measures [In Fact]. Wellington: Health Promotion Agency Research and Evaluation Unit, 2015.

20. Gendall P, Hoek J, Maubach N, et al. Public support for more action on smoking. *N Z Med J* 2013;126(1375):85-94. [published Online First: 2013/07/05]

21. Brennan E, Ilchenko E, Scollo M, et al. Public support for policies to phase out the retail sale of cigarettes in Australia: results from a nationally representative survey. *Tobacco Control* 2022:tobaccocontrol-2021-057122. doi: 10.1136/tobaccocontrol-2021-057122

22. Jaine R, Healey B, Edwards R, et al. How adolescents view the tobacco endgame and tobacco control measures: trends and associations in support among 14–15 year olds. *Tobacco Control* 2015;24(5):449-54. doi: 10.1136/tobaccocontrol-2013-051440

23. Li J, Newcombe R. Attitudes towards smoking and tobacco control strategies – a comparison of recent quit-attempters versus non-attempters. [In Fact]. Wellington: Health Promotion Agency Research and Evaluation Unit, 2013.

24. Newcombe R, Li J. Public Opinion on access to tobacco [In Fact]. Wellington: Health Promotion Agency Research and Evaluation Unit, 2013.

25. Farley SM, Coady MH, Mandel-Ricci J, et al. Public opinions on tax and retail-based tobacco control strategies. *Tobacco Control* 2015;24(e1):e10-e13. doi: 10.1136/tobaccocontrol-2013-051272

26. Sonnenberg J, Bostic C, Halpern-Felsher B. Support for Aggressive Tobacco Control Interventions Among California Adolescents and Young Adults. *Journal of Adolescent Health* 2020;66(4):506-09. doi: https://doi.org/10.1016/j.jadohealth.2019.11.302

27. Action on Smoking and Health. Smokefree: The First Ten Years. London: Action on Smoking and Health, 2017.

28. Action on Smoking and Health. The Smokefree Great Britain Survey: Public Opinion in England. London: Action on Smoking and Health, 2021.

34. Gallup. GALLUP POLL SOCIAL SERIES: CONSUMPTION HABITS, 2022.

35. Gendall P, Hoek J, Edwards R. What does the 2025 Smokefree Goal mean to the New Zealand public? *The New Zealand medical journal* 2014;127

48. White J. Young people's opinion on the sale of tobacco in New Zealand. [In Fact]. Wellington: Health Promotion Agency Research and Evaluation Unit, 2013.

49. Wang MP, Wang X, Lam TH, et al. The tobacco endgame in Hong Kong: public support for a total ban on tobacco sales. *Tobacco Control* 2015;24(2):162-67. doi: 10.1136/tobaccocontrol-2013-051092

50. Zatoński M, Herbeć A, Zatoński W, et al. Characterising smokers of menthol and flavoured cigarettes, their attitudes towards tobacco regulation, and the anticipated impact of the Tobacco Products Directive on their smoking and quitting behaviours: The EUREST-PLUS ITC Europe Surveys. *Tobacco Induced Diseases* 2018;16(2) doi: 10.18332/tid/96294
